# Benchmarking General-Purpose and Medical AI Large Language Models for Clinical Assessment and Management in Parkinson’s Disease

**DOI:** 10.64898/2026.05.13.26353021

**Authors:** Shechter Yosef, Klevor Raymond, Kouchache Trycia, Bouhadoun Sarah, Ronald B Postuma

## Abstract

**Background:** The clinical applicability of large language models (LLMs) in Parkinson’s disease (PD) management remains insufficiently characterized, particularly in generative responses to clinical vignette scenarios.

**Objective:** To evaluate the quality of clinical assessments and management plans generated by a general-purpose LLM (Gemini 1.5 Pro) and a medically specialized LLM (OpenEvidence), and to compare their performance.

**Methods:** Models generated free-text responses to 45 open clinical queries, focused on assessment of the situation, and recommended management plan. Two movement disorders fellows rated outputs using 5-point Likert scales, dichotomized into clinically appropriate (≥4) versus inappropriate (≤3). Discrepancies were adjudicated by a senior movement disorders specialist. Paired comparisons used McNemar’s test; qualitative analysis examined severe errors.

**Results:** Gemini 1.5 Pro and OpenEvidence showed high rates of clinically appropriate assessments (80.0% vs. 86.7%) but lower performance in management plans (48.9% vs. 57.8%). Cases in which both assessment and plan were clinically appropriate occurred in 46.7% and 55.6% of cases, respectively. None of these differences reached statistical significance. Severe errors were uncommon in assessments (6.7% vs. 8.9%) but more frequent in plans (26.7% in both), predominantly reflecting treatment strategy errors.

**Conclusions:** In generative clinical reasoning tasks involving Parkinson’s disease management vignettes, LLMs demonstrated reasonable performance in assessment, but consistent limitations in plan generation. The medically specialized LLM demonstrated several qualitative advantages but no statistically significant performance benefit over the general-purpose model. Therefore, these tools should be used with appropriate caution in Parkinson’s disease management, particularly regarding treatment recommendations.

## Introduction

The use of artificial intelligence (AI), and particularly large language models (LLMs), in medicine has expanded rapidly in recent years, driven by advances in deep learning, computational power, and large-scale data availability. These technologies are increasingly integrated across clinical practice, medical education, and research, where they are used to enhance efficiency, support decision-making, and streamline information processing (Thirunavukarasu et al. 2023; F. Liu et al. 2025; Clusmann et al. 2023).

One of the most prominent emerging applications of AI is its use as a clinical support tool for diagnostic reasoning and treatment planning during routine workflows (Bedi et al. 2025). A common approach to modeling and benchmarking this use case is to evaluate model performance on medical examination–style questions, which simulate scenarios in which clinicians pose clinical queries and assess the generated responses. Performance on these tasks has improved substantially over time, with accuracy increasing from approximately 60% in earlier models to around 90% in more advanced systems (Gilson et al. 2023; Saab et al. 2024; Fonseca et al. 2024; Natour et al. 2026). However, these benchmarks rely predominantly on multiple-choice formats and may not adequately capture real-world “open question” clinical reasoning (F. Liu et al. 2025).

Consistent with this limitation, recent reviews have described a gap between medical knowledge and practical clinical application, whereby model performance declines when applied to more complex or realistic scenarios, such as open-ended questions or cases with incomplete information. (Gong et al. 2025; Johri et al. 2025).

Comparative studies evaluating general-purpose versus domain-specific medical LLMs have yielded inconsistent findings. While earlier work suggested an advantage for domain-specific models due to specialized training (Singhal et al. 2025; Low et al. 2025; F. Liu et al. 2025; X. Liu et al. 2025), more recent evidence indicates that general-purpose models may outperform them in certain tasks, highlighting the importance of scale and reasoning capabilities over narrow domain specialization (Ray et al. 2025).

Evidence regarding LLM performance in neurology—and particularly in Parkinson’s disease and movement disorders—remains limited (Gorenshtein et al. 2026; Wunsch III and Hier 2026). Although some studies have evaluated performance in the diagnostic domain of Parkinson’s disease, there is a relative paucity of data addressing disease management and treatment planning.

Two key gaps remain: (1) limited evaluation of LLM performance on Parkinson’s disease– specific clinical questions, and (2) insufficient direct comparison between general-purpose and domain-specific medical models. Therefore, the aim of this study was to evaluate the performance of Gemini, a general-purpose LLM, and OpenEvidence, a domain-specific medical LLM, on Parkinson’s disease–related management clinical scenarios, and to directly compare the performance of these two model classes.

## Methods

### Clinical Vignette Development

A total of 50 common clinical topics frequently encountered during follow-up consultations for patients with Parkinson’s Disease (PD) were identified by the authors. Based on these topics, 50 original clinical vignettes were developed to simulate real-world neurological scenarios (see Supp. Table 1). These vignettes were synthetic and did not correspond to any specific individual, ensuring that no Protected Health Information (PHI) was utilized.

### Large Language Model (LLM) Selection and Technical Implementation

Two distinct LLMs were selected for benchmarking: a general-purpose model and a medically specialized model:

1. Gemini 1.5 Pro (Google LLC) The general-purpose model utilized was Gemini 1.5 Pro (Google LLC, Mountain View, CA). Data acquisition was conducted on November 17, 2025, via the native AI() function within Google Sheets. The testing environment was facilitated through Google Workspace Labs, an experimental preview program. Access was maintained through a personal Google account under the Google Generative AI Additional Terms of Service.
2. OpenEvidence The specialized medical model, OpenEvidence (OpenEvidence, Inc., Santa Monica, CA), was selected for its optimization toward evidence-based medical literature and real-time citation integration (OpenEvidence, n.d.). To facilitate large-scale querying, a custom Python script was developed to interface with the model (code available upon reasonable academic request). Due to the substantial length of OpenEvidence outputs, only the Assessment and Plan sections were manually extracted for rating, while supplementary explanations and citations were omitted. Both the full and shortened versions are provided in the Supplementary Materials. Access was authenticated through a verified healthcare provider account using a Medical Identification Number for Canada (MINC) to simulate standard clinical professional access.

### Prompting Strategy and Data Extraction

To ensure the generation of a specific Assessment and Plan (A&P), model-specific prompting was employed. Because LLMs respond differently to instructional structures, the prompts were tailored to each model to yield a consistent A&P format.

The prompt utilized for Gemini was:

> “*You are a neurologist specializing in movement disorders, with a focus on Parkinson’s disease. Read the clinical case and provide a concise, medically accurate response. Begin your assessment with a bold capital letter A, followed by a one-sentence summary stating the most likely diagnosis or clinical impression. Then, begin your plan with a bold capital letter P, and present a treatment plan in no more than two sentences. The plan must reflect a single, evidence-based therapeutic strategy. If you recommend medication, include specific medication details written in prescription format (drug name, dosage, route, and frequency). Do not offer multiple options—commit to the most appropriate treatment based on the case. Your response must be clinically precise, definitive, and concise. If you are unsure about the assessment or the optimal treatment plan, clearly write “I am not sure,” and specify the two options you are considering*”. Followed by the case vignette.

The prompt utilized for OpenEvidence was:

> “*Write an assessment and plan for the following case. Decide and commit to a single treatment recommendation. also be specific in dosages if the recommendation involves a medication*”. Followed by the case vignette.

All queries were performed using a zero-shot approach, meaning each clinical vignette was processed in a new, independent session without prior context to prevent inter-case learning or bias. Responses were extracted and saved into a structured database for subsequent rating (see Figure 1, Supp. Table 4, Supp. Table 5).

**Figure 1.**
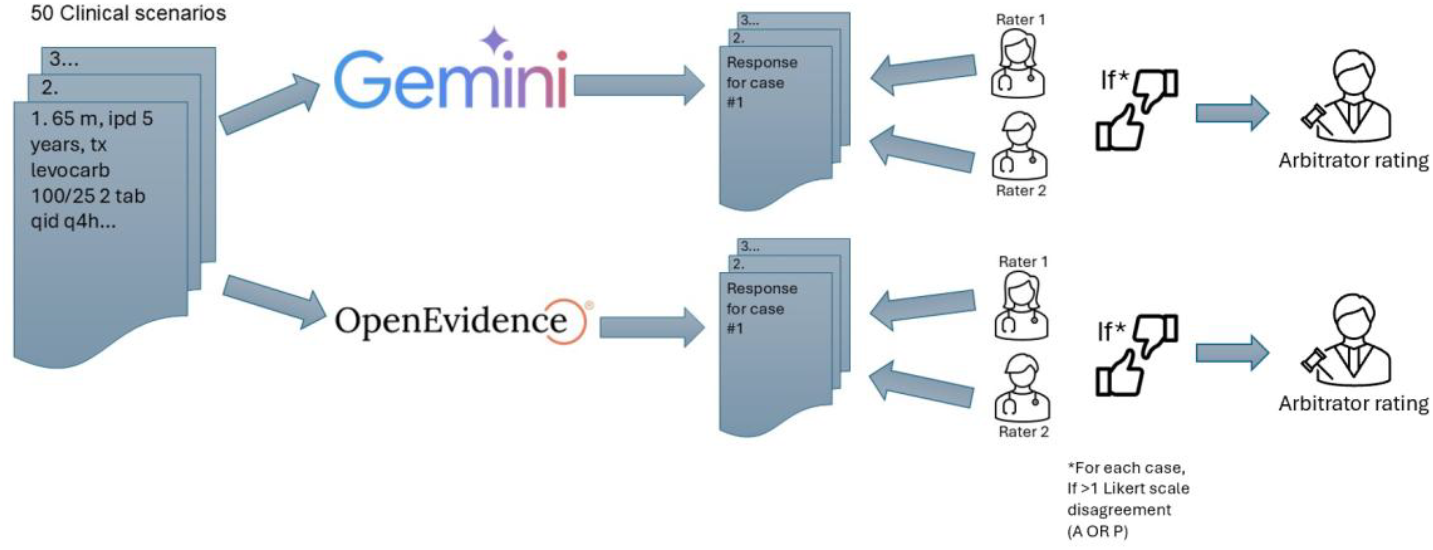
Schematic representation of the study workflow and research methodology.

### Evaluation Framework and Scoring

Outputs were independently evaluated by two neurologists (YS and RK) currently in advanced stages of fellowship training in Parkinson’s disease and movement disorders at the Montreal Neurological Institute, McGill University, Montreal, Canada. The evaluation comprised three distinct metrics:

Clinical performance was evaluated using two 5-point Likert scales (See Supp. Table 2 and 3 for full Likert scale definitions):

- Assessment (1: Completely Incorrect; 2: Significantly Incorrect; 3: Too General; 4: Good; 5: Completely Accurate),
- Plan (1: Unsafe/Dangerous; 2: Inappropriate; 3: Acceptable but Suboptimal; 4: Good/Appropriate; 5: Optimal/Precise).

The Assessment scale measured the accuracy of the clinical impression and identification of primary issues, while the Plan scale quantified the safety, appropriateness, and evidence-based quality of therapeutic recommendations.

In instances in which Likert scores assigned by the two primary raters differed by more than one point, a third senior Movement Disorders expert neurologist served as an adjudicator to establish a consensus rating (RP). In addition, for the dichotomized binary analysis (clinically appropriate vs. inappropriate), any 1-point disagreement crossing the appropriateness threshold (i.e., ratings of 3 vs. 4) was predefined to undergo additional adjudication by the senior neurologist. For cases in which arbitration was not required, the lower of the two ratings was retained. This approach was selected to align with the study’s objective of evaluating clinical appropriateness, favoring a more conservative and stringent assessment framework.

### Statistical Analysis

All statistical analyses were performed using IBM SPSS Statistics for Windows, Version 31.0 (IBM Corp., Armonk, NY, USA). Descriptive statistics were used to summarize performance scores. Paired comparisons between models were conducted using the McNemar test, which is appropriate for paired binary outcomes because both models were evaluated on the same clinical cases. A conventional chi-square test would not be appropriate because it assumes independence of observations, and Cohen’s kappa assesses agreement rather than directional differences in performance. The McNemar test specifically evaluates whether discordant classifications occur asymmetrically between two related methods. A p-value of <.05 was considered statistically significant.

### Ethical Considerations and Disclosures

As this study utilized exclusively synthetic clinical vignettes developed by the authors and did not involve human subjects, medical records, or identifiable patient data, it did not meet the criteria for human subject research. The McGill University Health Centre Research Ethics Board reviewed the study protocol and formally determined that it was exempt from institutional review board (IRB) approval requirements.

The authors declare no financial or professional affiliation with OpenEvidence, Inc. or Google LLC. This research was conducted independently; the platform developers had no role in study design, data collection, analysis, or manuscript preparation.

## Results

### Case Eligibility

Prior to analysis, all 50 cases were screened for data sufficiency. Each clinical vignette was independently evaluated by the two raters to determine whether sufficient clinical information was available to allow meaningful evaluation of the model-generated responses. Cases were excluded only when both raters judged the vignette to contain insufficient information. This approach was selected to avoid unnecessary exclusion of potentially evaluable cases, under the rationale that if at least one clinician considered the vignette sufficiently detailed, the scenario could reasonably be considered answerable by the LLM. Following this process, 45 cases remained for final analysis

### Adjudicated model performance

To facilitate clinically meaningful interpretation, Likert-scale ratings were dichotomized into a binary outcome reflecting clinical usability. Ratings of 4 or higher were classified as “clinically appropriate,” representing responses deemed suitable for clinical use, whereas ratings of 3 or lower were considered not clinically appropriate.

For assessment, across the 45 included cases, both models demonstrated high rates of clinically appropriate assessments, with OpenEvidence achieving 86.7% performance and Gemini 1.5 Pro receiving 80.0%.

In contrast, performance in the ‘plan’ domain was substantially lower for both models, with clinically appropriate plans observed in 57.8% of cases for OpenEvidence and 48.9% for Gemini 1.5 Pro. Overall, these findings indicate a consistent pattern in which both models perform better in diagnostic assessment than in treatment planning, with a numerically higher performance of OpenEvidence across both domains (Table 1).

A stricter composite outcome required both the assessment and plan to be rated as clinically appropriate (Likert ≥4), reflecting potential clinical usability. Using this definition, performance declined for both models, with OpenEvidence achieving appropriate composite outcomes in 55.6% of cases (25/45) compared to 46.7% (21/45) for Gemini 1.5 Pro, underscoring the greater difficulty of generating appropriate management plans (Table 1).

Paired comparisons using McNemar’s test demonstrated no statistically significant differences between Gemini 1.5 Pro and OpenEvidence across all evaluated outcomes, including assessment (p = 0.453), plan (p = 0.523), and the composite outcome (p = 0.523) (Table 1, Figure 2).

**Table 1:** Proportion of Clinically Appropriate Ratings for each model (Assessment and Plan)

| Outcome (Clinically Appropriate Scales) | Gemini 1.5 Pro (%; n/N) | OpenEvidence (%; n/N) | p-value (McNemar) |
| --- | --- | --- | --- |
| Assessment | 80.0%; 36/45 | 86.7%; 39/45 | 0.453 |
| Plan | 48.9%; 22/45 | 57.8%; 26/45 | 0.523 |
| Composite (Assessment + Plan) | 46.7%; 21/45 | 55.6%; 25/45 | 0.523 |

**Figure 2.**
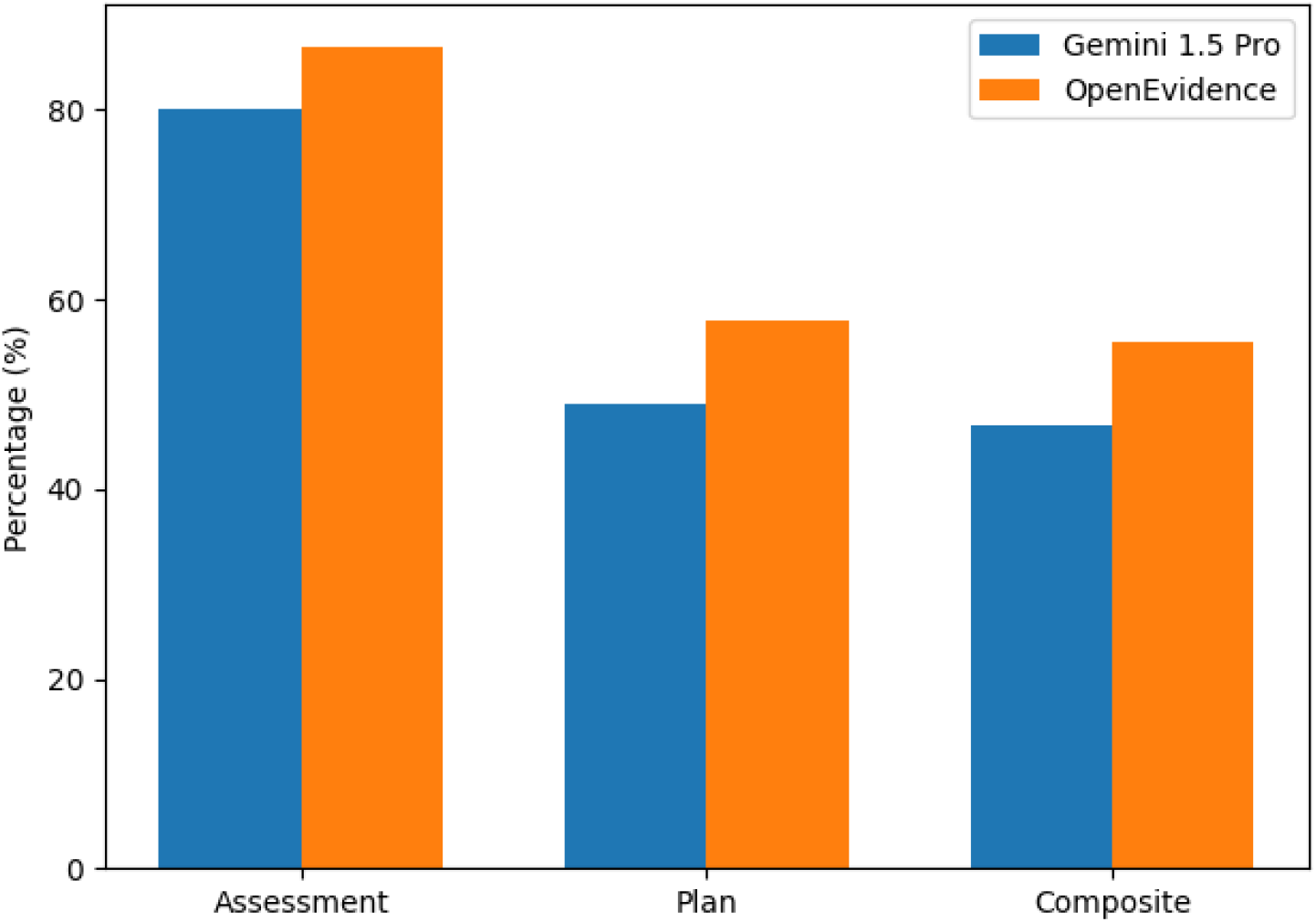
Clinically Appropriate ratings by model

### Error severity

Error severity analysis demonstrated distinct patterns between assessment and plan domains. Examples of errors are listed in the supplementary section (see Supp. Figure 1). In the assessment domain, both models exhibited relatively low rates of severe errors (Likert scores 1, “Completely Incorrect” or 2, Partially “Significantly Incorrect”), observed in 6.7% (3/45) of cases for Gemini 1.5 Pro and 8.9% (4/45) for OpenEvidence. Moderate errors (score 3, “Too General / Acceptable but Not Focused”) occurred in 11.1% (5/45) and 4.4% (2/45) of cases, respectively.

In contrast, the recommended plan demonstrated substantially higher error rates, with severe errors (Likert score 1; “Unsafe / Incorrect / Dangerous”, and 2; “Inappropriate Overly Aggressive or Overly Conservative”) observed in 26.7% (12/45) of cases for both models. Moderate errors (Likert score 3; “Acceptable but Suboptimal”) were also more frequent in the plan section, occurring in 24.4% (11/45) of cases for Gemini 1.5 Pro and 15.6% (7/45) for OpenEvidence. Overall, these findings indicate that while both models rarely produced severely incorrect assessments, errors in treatment planning were considerably more common, with no difference between models in the rate of severe errors (Table 2).

**Table 2:**
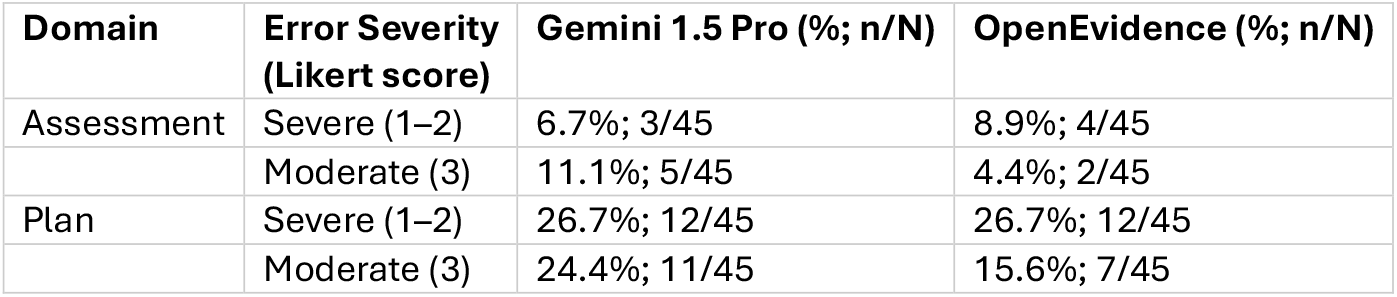
Error Severity Distribution by Model and Domain.

### Qualitative analysis of severe errors

A qualitative analysis was performed for all responses rated as clinically inappropriate (Likert scores 1–2). Individual errors were reviewed and categorized into three domains: diagnostic errors, treatment strategy errors, and execution errors, as summarized in Table 3 and detailed in Supplementary Tables 6-7.

**Table 3:** Classification and Frequency of Error Types Identified in the Qualitative Analysis of Clinically Inappropriate Responses (Likert Scores 1–2). Error categories were not mutually exclusive; individual cases could contribute to more than one category.

| Error Category | Definition / Included Errors | Gemini 1.5 Pro (n) | OpenEvidence (n) |
| --- | --- | --- | --- |
| Diagnostic Errors | Incorrect diagnosis, overestimation of severity, premature diagnostic closure | 4 | 2 |
| Treatment Strategy Errors | Suboptimal treatment choice, incorrect treatment sequencing, failure to optimize therapy | 7 | 9 |
| Execution Errors | Incorrect dosing, inappropriate duplication, titration/tapering errors | 4 | 4 |

Treatment strategy errors were the predominant error type in both models, identified in 7 cases for Gemini 1.5 Pro and 9 cases for OpenEvidence. Diagnostic errors were less frequent (4 vs. 2 cases, respectively), while execution errors occurred at similar frequencies in both models (4 cases each). A predominance of treatment strategy errors was observed in both models. Error categories were not mutually exclusive, and individual cases could be assigned to multiple categories.

### Error discordances

A case-level comparison of errors rated as clinically inappropriate (Likert scores 1–2) demonstrated limited overlap between models. Both Gemini 1.5 Pro and OpenEvidence produced clinically inappropriate responses in the same cases in only 11.1% of instances (5/45). In contrast, discordant errors were more common, occurring in 33.3% of cases (15/45), with OpenEvidence alone producing clinically inappropriate responses in 17.8% (8/45) and Gemini 1.5 Pro in 15.6% (7/45). These findings indicate that most clinically inappropriate responses were model-specific rather than shared.

## Discussion

This study found that when large language models were provided with vignettes based on common clinical scenarios in Parkinson’s disease, limitations were particularly evident in the generation of management plans. Some recommendations raised clinically relevant safety concerns if applied without physician oversight. A numerical trend favoring the specialized medical model was observed; however, no statistically significant difference was identified between models.

### Assessment–Plan Performance Discrepancy

The study consistently demonstrated a marked discrepancy between model performance in clinical assessment and treatment planning. While both models generally provided accurate assessments, their performance declined substantially when generating planning recommendations. This pattern was observed across all analyses and in both models, without a statistically significant difference between the general-purpose and the domain-specific medical model, although a trend favoring the latter was noted. These findings are consistent with the current literature findings (Rao et al. 2026; T.-H. Wang et al. 2025; Patel and Blumenthal 2026; Hager et al. 2024; F. Liu et al. 2025; Zhou et al. 2025).

### Error Types

Qualitative error analysis further supports these findings, demonstrating that treatment-related errors predominate over diagnostic inaccuracies. Beyond this, several additional insights emerge.

Both models exhibited limitations in therapeutic prioritization and sequencing, frequently selecting suboptimal strategies or misplacing treatments within the clinical hierarchy. Second, there was a consistent tendency toward “add-on prescribing,” whereby new therapies were introduced without prior optimization of existing treatment regimens. These patterns were similar across both models, suggesting shared limitations that are not substantially mitigated by domain-specific medical training (Hager et al. 2024). This observation is particularly relevant in Parkinson’s disease, where management requires individualized decision-making and careful balancing of treatment benefits and side effects in a complex disease with motor, neuropsychiatric, sleep, cognitive, and autonomic symptoms. (Foltynie et al. 2024; Pham Nguyen et al. 2024; Scott et al. 2013). Finally, model outputs may contain multiple simultaneous inaccuracies and should therefore be evaluated comprehensively rather than along a single dimension.

### Error Discordances

The low overlap in clinically inappropriate responses between models suggests that errors are primarily model-specific rather than case-dependent. If case characteristics were the main driver, a higher rate of shared failures would be expected. Instead, the findings indicate that each model applies distinct internal reasoning processes when interpreting the same input. This observation raises the theoretical possibility that combining outputs from multiple models may improve case-level accuracy by mitigating model-specific errors(Z. Wang et al. 2025).

### Overconfidence

Although the prompting strategy for Gemini explicitly allowed the expression of uncertainty, this option was not utilized by the model. While this may partially reflect prompt design, the consistent absence of uncertainty suggests a tendency toward overconfidence (Omar et al. 2024). This has important implications for clinical use. Unlike clinicians, who are trained to recognize and communicate uncertainty, LLMs may present responses with overly definitive phrasing. Therefore, clinicians should critically appraise model outputs, regardless of how definitive or authoritative they appear. This issue becomes especially problematic if patients are accessing advice directly, without mitigation by a clinician.

### Qualitative Characteristics of the Medical Model

Despite the absence of statistically significant differences in primary outcomes, several qualitative distinctions were observed. OpenEvidence generated more detailed responses, frequently including supporting evidence and accessible citations without explicit prompting. While this did not translate into superior clinical performance (Pearson et al. 2026; Ben Shitrit et al. 2026; Chen et al. 2025; Rosen et al. 2025; Jagarapu et al. 2025), the ability to directly review cited sources may facilitate more critical appraisal of model-generated recommendations by the clinician. The model also more commonly provided alternative therapeutic pathways, adverse effect considerations, and monitoring recommendations, reflecting a more structured management approach, albeit at the cost of longer outputs. Finally, unlike publicly accessible general-purpose LLMs, OpenEvidence is restricted to licensed medical professionals. In light of the present findings, this may represent an important practical safeguard, as appropriate use of current LLM systems still depends heavily on the clinician’s ability to critically evaluate generated outputs before applying them in practice.

### Limitations

This study has several limitations. Clinical appropriateness was evaluated against expert clinician judgment, which itself may be subject to variability and bias. There is subjectivity in rating error severity; what one physician may describe as a mild omission, another may find highly inappropriate. The process of adjudication was designed to mitigate these issues, and the specific models responses are provided in the msupplemental section so that readers can judge for themselves whether the model’s responses were accurate. The relatively small cohort limits statistical power. Model performance reflects a single time point and may change with ongoing updates (F. Liu et al. 2025). Additionally, prompting strategies may influence outputs, and the use ofn different prompts across models limits direct comparability (L. Wang et al. 2024; J. Liu et al. 2025). Moreover, models were evaluated using structured clinical vignettes with relatively clear clinical framing, and therefore their performance in more realistic free-flowing conversational or non-professional chat-based interactions remains uncertain. Finally, our findings are specific to Parkinson’s disease and may not generalize to other clinical domains.

## Conclusion

In summary, in generative clinical reasoning tasks involving Parkinson’s disease management vignettes, both models demonstrated generally good assessment performance, whereas treatment planning performance was comparatively suboptimal. A modest numerical advantage was observed for the specialized medical model, although no statistically significant difference was identified. These findings suggest that LLM-generated outputs should be interpreted with caution, particularly with respect to therapeutic recommendations. At present, these systems may serve as adjunctive tools for clinical reasoning when used by clinicians capable of critically evaluating their outputs. However, they are not sufficiently reliable to function as independent decision-makers, particularly in complex clinical domains such as Parkinson’s disease.

## Supporting information

see Supp. Table 1

## Data Availability

All data produced in the present work are contained in the manuscript

## Acknowledgments

The authors thank Tal Klap for technical support. No financial support or funding was received for this work.

## References

Bedi, Suhana, Yutong Liu, Lucy Orr-Ewing, et al. 2025. “Testing and Evaluation of Health Care Applications of Large Language Models: A Systematic Review.” JAMA 333 (4): 319–28. 10.1001/jama.2024.21700.

Ben Shitrit, Itamar, Daphna Idan, Mark Volevich, et al. 2026. “Real World Human-LLM Interactions – Prospective Blinded versus Unblinded Expert Physician Assessments of LLM Responses to Complex Medical Dilemmas.” PLOS Digital Health 5 (3): e0001278. 10.1371/journal.pdig.0001278.

Chen, Shan, Mingye Gao, Kuleen Sasse, et al. 2025. “When Helpfulness Backfires: LLMs and the Risk of False Medical Information Due to Sycophantic Behavior.” Npj Digital Medicine 8 (1): 605. 10.1038/s41746-025-02008-z.

Clusmann, Jan, Fiona R. Kolbinger, Hannah Sophie Muti, et al. 2023. “The Future Landscape of Large Language Models in Medicine.” Communications Medicine 3 (1): 141. 10.1038/s43856-023-00370-1.

Foltynie, Tom, Veronica Bruno, Susan Fox, Andrea A. Kühn, Fiona Lindop, and Andrew J. Lees. 2024. “Medical, Surgical, and Physical Treatments for Parkinson’s Disease.” The Lancet 403 (10423): 305–24. 10.1016/S0140-6736(23)01429-0.

Fonseca, Ângelo, Axel Ferreira, Luís Ribeiro, Sandra Moreira, and Cristina Duque. 2024. “Embracing the Future-Is Artificial Intelligence Already Better? A Comparative Study of Artificial Intelligence Performance in Diagnostic Accuracy and Decision-Making.” European Journal of Neurology 31 (4): e16195. 10.1111/ene.16195.

Gilson, Aidan, Conrad W. Safranek, Thomas Huang, et al. 2023. “How Does ChatGPT Perform on the United States Medical Licensing Examination (USMLE)? The Implications of Large Language Models for Medical Education and Knowledge Assessment.” JMIR Medical Education 9 (1): e45312.10.2196/45312.

Gong, Eun Jeong, Chang Seok Bang, Jae Jun Lee, and Gwang Ho Baik. 2025. “Knowledge-Practice Performance Gap in Clinical Large Language Models: Systematic Review of 39 Benchmarks.” Journal of Medical Internet Research 27 (1): e84120. 10.2196/84120.

Gorenshtein, Alon, Kamel Shihada, Mahmud Omar, Yiftach Barash, Girish N. Nadkarni, and Eyal Klang. 2026. “Large Language Models in Clinical Neurology: A Systematic Review.” Preprint, Research Square, February 18. 10.21203/rs.3.rs-8902070/v1.

Hager, Paul, Friederike Jungmann, Robbie Holland, et al. 2024. “Evaluation and Mitigation of the Limitations of Large Language Models in Clinical Decision-Making.” Nature Medicine 30 (9): 2613–22. 10.1038/s41591-024-03097-1.

Jagarapu, Jawahar, Kikelomo Babata, Surya Chamarthi, and Robert Hoyt. 2025. “The Accuracy and Repeatability of OpenEvidence on Complex Medical Subspecialty Scenarios: A Pilot Study.” Preprint, medRxiv, December 4.10.64898/2025.11.29.25341091.

Johri, Shreya, Jaehwan Jeong, Benjamin A. Tran, et al. 2025. “An Evaluation Framework for Clinical Use of Large Language Models in Patient Interaction Tasks.” Nature Medicine 31 (1): 77–86. 10.1038/s41591-024-03328-5.

Liu, Fenglin, Hongjian Zhou, Boyang Gu, et al. 2025. “Application of Large Language Models in Medicine.” Nature Reviews Bioengineering 3 (6): 445–64. 10.1038/s44222-025-00279-5.

Liu, Jialin, Fang Liu, Changyu Wang, and Siru Liu. 2025. “Prompt Engineering in Clinical Practice: Tutorial for Clinicians.” Journal of Medical Internet Research 27 (September): e72644. 10.2196/72644.

Liu, Xiaohong, Hao Liu, Guoxing Yang, et al. 2025. “A Generalist Medical Language Model for Disease Diagnosis Assistance.” Nature Medicine 31 (3): 932–42. 10.1038/s41591-024-03416-6.

Low, Yen Sia, Michael L. Jackson, Rebecca J. Hyde, et al. 2025. “Answering Real-World Clinical Questions Using Large Language Model, Retrieval-Augmented Generation, and Agentic Systems.” Digital Health 11 (June): 20552076251348850. 10.1177/20552076251348850.

Natour, Dania El, Mohamad Abou Alfa, Ahmad Chaaban, Reda Assi, Toufic Dally, and Bahaa Bou Dargham. 2026. “Performance of 5 AI Models on United States Medical Licensing Examination Step 1 Questions: Comparative Observational Study.” JMIR AI 5 (1): e76928. 10.2196/76928.

Omar, Mahmud, Benjamin S. Glicksberg, Girish N. Nadkarni, and Eyal Klang. 2024. “Overconfident AI? Benchmarking LLM Self-Assessment in Clinical Scenarios.” Preprint, medRxiv, August 11. 10.1101/2024.08.11.24311810.

OpenEvidence (OpenEvidence). n.d. “About.” Accessed January 6, 2026. https://www.openevidence.com.

Patel, Bakul, and David Blumenthal. 2026. “A Novel Approach to Overseeing the Clinical Application of Generative AI.” JAMA Health Forum 7 (3): e256947.10.1001/jamahealthforum.2025.6947.

Pearson, Joe, Itiel E. Dror, Emma Jayes, Grace-Rose Whordley, Georgina Mason, and Sophie Nightingale. 2026. “Examining Human Reliance on Artificial Intelligence in Decision Making.” Scientific Reports 16 (1): 5345 10.1038/s41598-026-34983-y.

Pham Nguyen, Thanh Phuong, Dylan Thibault, Ali G. Hamedani, and Allison W. Willis. 2024. “Attitudes and Beliefs towards Medication Burden and Deprescribing in Parkinson Disease.” BMC Neurology 24 (1): 325. 10.1186/s12883-024-03830-w.

Rao, Arya S., Kaiz P. Esmail, Richard S. Lee, et al. 2026. “Large Language Model Performance and Clinical Reasoning Tasks.” JAMA Network Open 9 (4): e264003. 10.1001/jamanetworkopen.2026.4003.

Ray, Sushant Kumar, Ebad Shabbir, Rafiq Ali, Abdullah Mohammed, and Samar Wazir. 2025. “Beyond Specialization: A Comprehensive Evaluation of General-Purpose Versus Medical Domain-Specific Large Language Models for Biomedical Question Answering.” 2025 IEEE International Conference on Data Mining Workshops (ICDMW), November, 478–84. 10.1109/ICDMW69685.2025.00060.

Rosen, Kyra L., Margaret Sui, Kimia Heydari, Elizabeth J. Enichen, and Joseph C. Kvedar. 2025. “The Perils of Politeness: How Large Language Models May Amplify Medical Misinformation.” NPJ Digital Medicine 8 (November): 644. 10.1038/s41746-025-02135-7.

Saab, Khaled, Tao Tu, Wei-Hung Weng, et al. (arXiv.Org). 2024. “Capabilities of Gemini Models in Medicine.” April 29. https://arxiv.org/abs/2404.18416v2.

Scott, Ian A., Leonard C. Gray, Jennifer H. Martin, Peter I. Pillans, and Charles A. Mitchell. 2013. “Deciding When to Stop: Towards Evidence-Based Deprescribing of Drugs in Older Populations.” Evidence Based Medicine 18 (4): 121–24. 10.1136/eb-2012-100930.

Singhal, Karan, Tao Tu, Juraj Gottweis, et al. 2025. “Toward Expert-Level Medical Question Answering with Large Language Models.” Nature Medicine 31 (3): 943–50. 10.1038/s41591-024-03423-7.

Thirunavukarasu, Arun James, Darren Shu Jeng Ting, Kabilan Elangovan, Laura Gutierrez, Ting Fang Tan, and Daniel Shu Wei Ting. 2023. “Large Language Models in Medicine.” Nature Medicine 29 (8): 1930–40. 10.1038/s41591-023-02448-8.

Wang, Li, Xi Chen, XiangWen Deng, et al. 2024. “Prompt Engineering in Consistency and Reliability with the Evidence-Based Guideline for LLMs.” Npj Digital Medicine 7 (1): 41. 10.1038/s41746-024-01029-4.

Wang, Te-Hao, Jing-Cheng Jheng, Yen-Ting Tseng, Li-Fu Chen, and Yu-Chun Chen. 2025. “Evaluating GPT-4’s Visual Interpretation and Clinical Reasoning on Emergency Settings: A 5-Year Analysis.” Journal of the Chinese Medical Association: JCMA 88 (9): 672–80. 10.1097/JCMA.0000000000001273.

Wang, Zifeng, Hanyin Wang, Benjamin Danek, et al. 2025. “A Perspective for Adapting Generalist AI to Specialized Medical AI Applications and Their Challenges.” Npj Digital Medicine 8 (1): 429. 10.1038/s41746-025-01789-7.

Wunsch III, Donald C., and Daniel B. Hier. 2026. “Large Language Models for Neurology: A Mini Review.” Frontiers in Digital Health 7 (January).10.3389/fdgth.2025.1732759.

Zhou, Shuang, Zidu Xu, Mian Zhang, et al. 2025. “Large Language Models for Disease Diagnosis: A Scoping Review.” Npj Artificial Intelligence 1 (1): 9. 10.1038/s44387-025-00011-z.

