## Supplementary material for "Benchmarking General-Purpose and Medical AI Large Language Models for Clinical Assessment and Management in Parkinson’s Disease": see Supp. Table 1

### Supplementary Tables

#### Supp. Table 1 -List of cases:

| case number | case |
|---|---|
| 1 | 65 m, ipd 5 years, tx levocarb 100/25 2 tab qid q4h, safinamide 100 mg qd. Reports involuntary movements of the limbs during the day. He usually does not notice this himself, but his wife asks him why he cannot sit comfortably in a chair. There is fluctuation during the day. It seems to him that it usually happens about an hour after taking LEVOCARB, but not necessarily. It does not interfere with his function, but it bothers his wife to see him like this. |
| 2 | 66 f, ipd 7 years, tx levocarb 100/25 1.5 tab qid q4h, Levocrab CR 200/50 QHS, rasagiline 1mg qd, entacapone 200 md QID. Myrbetriq 50 qd, laxaday 17 g qd. Describes a recurrence of the tremor in her right hand, a general bad feeling including anxiety, and FREEZING OF GAIT events quite regularly about 40 minutes before she is supposed to take her next dose of LEVOCARB. |
| 3 | 75 m, ipd 10 years, tx levocarb 100/25 2 tab qid q4h, Levocrab CR 200/50 QHS, rasagiline 1mg qd, entacapone 200 md QID. Myrbetriq 50 qd. Random events are reported during the day where after taking LEVOCARB it takes a long time - sometimes more than an hour - for the drug to start working. takes the LEVOCARB 30 min before meals. |
| 4 | 75 f, ipd 10 years, tx levocarb 100/25 2 tab qid q4h, Levocrab CR 200/50 QHS, rasagiline 1mg qd, entacapone 200 md QID. Myrbetriq 50 qd. Laxaday 17 gr QD. Describes that at least once a day and sometimes more often there are events in which she does not go ON after taking LEVOCARB. States that she takes LEVOCARB half an hour before meals. And has regular bowel movements every day. |

| | |
|---|---|
| 5 | 59 m, ipd 5 years, tx levocarb 100/25 1 tab qid q4h, rasagiline 1mg qd, Rotigotine patch 3 mg qd, Myrbetriq 25 qd. Laxaday 17 gr QD. Describes waking up in the morning with severe tremors, a feeling of muscle stiffness, slow walking, and a general feeling of ill health. |
| 6 | 59 m, ipd 2 years, tx levocarb 100/25 2 tab tid at 30 min before meals+ 1 tab HS, Laxaday 17 gr QD. The patient reports that since he began treatment with LEVOCRAB about a year ago, the doctors have tried to gradually increase his dose of LEVOCARB since there was no beneficial effect on the tremor. It should be noted that there was an improvement in other symptoms, such as the feeling of muscle stiffness decreasing, and walking becoming easier and faster. |
| 7 | 63 f, ipd 1 year, tx levocarb 100/25 1 tab tid at 30 min before meals+ 1 tab HS, Laxaday 17 gr QD. Reports 2 falls during the last six months. Occurred while walking on the street. Stumbled due to lack of attention to a prominent area on the sidewalk. Falls forward. No feeling of presyncope before the falls. Time relationship to drug treatment is unknown. |
| 8 | 68 f, ipd 6 years, tx levocarb 100/25 1.5 tab qid q4h, rotigotine patch 3 mg qd, amantadine 100 mg bid. She has a shuffling, unsteady gait. Family members note that she walks with short, hesitant steps and sometimes her feet seem to accelerate uncontrollably (festination). She has not fallen yet, but she feels very unstable especially when distracted or walking in crowded areas. |
| 9 | 78 m, ipd 12 years, tx levocarb 100/25 2 tab qid q4h, entacapone 200 mg qid, rasagiline 1 mg qd. He has increasing memory problems and difficulty following conversations. His wife reports that he often forgets recent events, misplaces items around the house, and needs repeated reminders for appointments. He also has trouble concentrating and planning simple tasks, which marks a decline from his previously sharp cognitive abilities. |
| 10 | 80 f, ipd 10 years, tx levocarb 100/25 2 tab qid q3h (8-11-14-17-20), safinamide 100 qd, amantadine 100 mg bid. She reports seeing children in her living room who aren't actually there, even in the middle of the day. Sometimes she also sees small animals darting around, or thinks she sees her deceased relatives momentarily. She is often able to recognize these visions are not real once she checks, but they are frightening and have caused her anxiety. |
| 11 | 65 f, ipd 6 years, tx levocarb 100/25 1 tab tid, selegiline 5 mg bid, sertraline 50 mg qd. She feels persistently sad and tearful with little enjoyment in activities she used to love. Her family notes she has become withdrawn, often sitting quietly and appearing downcast. She struggles with low energy and poor appetite, and sometimes she even wonders if life is worth living despite being on an antidepressant for the last year. The patient denies any suicidal thoughts. |
| 12 | 60 m, ipd 4 years, tx levocarb 100/25 1 tab qid q4h Rasagiline 1 mg qd, clonazepam 0.5 mg QHS prn. He constantly feels on edge and has episodes of intense anxiety, especially as his medication dose wears OFF. He worries excessively about small things, and before his next dose is due, he often becomes shaky, sweaty, and panic-stricken. |
| 13 | 70 m, ipd 8 years, tx levocarb 100/25 2 tab qid, entacapone 200 mg qid, Safinamide 100 qd. Spending most of the day sitting in his armchair doing nothing unless prompted. He does not initiate activities or hobbies he once enjoyed, but he does not report feeling sad or depressed — just indifferent. His wife reports she has to encourage him to get started on any task, as he otherwise lacks motivation to even get dressed or go for a short walk. |

| | |
|---|---|
| 14 | 55 m, ipd 7 years, tx levocarb 100/25 2 tab qid q4h, safinamide 100 mg qd, citalopram 20 mg qd, Laxaday 17 g qd. He tends to take more LEVOCARB than prescribed, often taking an extra half or full tablet whenever he feels his energy dipping. The family reports recurrent episodes of disinhibited speech, at times with sexual content, occasionally leading to problematic situations such as inappropriate remarks toward unfamiliar women, for example, a cashier at the supermarket. |
| 15 | 63 f, ipd 8 years, tx levocarb 100/25 1.5 tab qid q4h, Rasagiline 1 mg qd, citalopram 10 mg qd, laxaday 17 g qd prn. She has difficulty sleeping through the night, waking up every few hours and struggling to get back to sleep. She feels unrefreshed in the morning. There is no difficulty with initial sleep onset. The patient is unable to clearly identify the reason for awakening during the night; at times it is to use the bathroom, though not consistently. She does not experience any difficulty moving to the bathroom upon awakening. |
| 16 | 58 m, ipd 6 years, tx levocarb 100/25 2 tab qid q4h, entacapone 200 mg qid, citalopram 20 mg qd, myrbetriq 50 mg qd. He struggles with overwhelming sleepiness during the day and often nods off in the middle of activities. He has fallen asleep while watching TV and even briefly while eating lunch. Because he once nearly dozed off while driving, he no longer drives. |
| 17 | 64 f, ipd 4 years, tx levocarb 100/25 1.5 tab tid, gabapentin 300 mg qhs. She experiences aching muscle pain in her shoulders and arms that tends to worsen when her Parkinson's symptoms are more pronounced. The chronic pain significantly affect her quality of life, often requiring use of pain relievers, with various outcomes. |
| 18 | 71 m, ipd 9 years, tx levocarb 100/25 2 tab qid q4h, entacapone 200 mg qid, Laxaday 17 g qd. He has significant urinary urgency and frequency, needing to rush to the bathroom almost every 2 hours. He often cannot hold his urine long enough and has had several accidents. Nights are also troublesome, with him waking to urinate multiple times (nocturia), which disrupts his and his wife's sleep. |
| 19 | 75 f, ipd 8 years, tx levocarb 100/25 1.5 tab qid q4h, senna 2 tabs qhs, citalopram 10 md qd, rivastigmine patch 4.6 mg qd. She reports bowel movements only every 3–4 days despite taking daily laxatives, high-fiber diet and plenty of fluids. She often feels bloated and uncomfortable, and has to strain hard to pass stool. |
| 20 | 73 m, ipd 10 years, tx levocarb 100/25 2 tab 5 times a day q3h (7-10-13-16-19), Sin CR 200/50 mg QHS, Safinamide 100 mg qd, Rivastigmine patch 9.6 mg qd, Laxadya 17 g qd PRN. He feels light-headed and dizzy whenever he stands up from a sitting or lying position. On several occasions, he nearly blacked out after getting out of bed in the morning, needing to sit back down to avoid fainting. |
| 21 | 62 f, ipd 6 years, tx levocarb 100/25 1 tab qid q4h, Rasagiline 1 mg qd, amantadine 100 mg tid. She experiences overwhelming sense of exhaustion that is not relieved by rest. Even after a full night's sleep, she often feels she has no energy to do her usual activities or exercises. |
| 22 | 55 m, ipd 3 years, tx Levocarb 100/25 1.5 tab tid, citalopram 20 qd. Speaks with a very low volume and a hoarse, monotone voice that makes him difficult to hear. His coworkers often ask him to repeat himself, and he feels frustrated that his voice doesn't carry. he struggle sto speak loudly and clearly on the phone or in meetings. |
| 23 | 66-year-old, F IPD 10-year duration, Tx Levocarb 100/25 2 tabs 5 times a day, Sinemet CR 200/50 1 tab qhs, safinamide 100 md qd, Citalopram 20 mg qd, melatonin 10 mg qhs. complaining about bothersome drooling. no conection to wearing off times. No Treatment was tried. |

| | |
|---|---|
| 24 | 75 m, ipd 11 years, tx levocarb 100/25 2 tab qid q4h, levocarb CR 200/50 QHS, Safinamide 100 mg qd. He often coughs or chokes when swallowing, particularly with liquids. He needs his food cut into small pieces and sometimes avoids tough or dry foods that are hard to swallow. No connection for wearing off times. |
| 25 | 67 m, ipd 7 years, tx levocarb 100/25 1.5 tab qid q4h, entacapone 200 mg qid. She struggles with the basic tasks of eating, such as using utensils. Her hands tremble so much that food often falls off her fork before she can get it to her mouth. She also finds it hard to cut her food because of limited dexterity and strength in her hands. As a result, she often needs assistance or adaptive utensils to eat without making a mess. |
| 26 | 69 m, ipd 8 years, tx levocarb 100/25 2 tab qid q3h, Rasagiline 1 mg qd, myrbetriq 50 mg qd. laxaday 17 g qd prn. He often needs his wife's help to put on his shoes and socks, as balancing on one leg and bending over is challenging. What used to take him 5 minutes now can consume a half hour just to get dressed in the morning. |
| 27 | 72 f, ipd 10 years, tx levocarb 100/25 2 tab qid q3h, entacapone 200 mg qid. Safinamide 100 mg qd, laxaday 17 g qd. She finds it hard to shower; she's afraid of losing balance in the slippery tub and her slow movements make it exhausting to wash and dry herself. She has started relying on her daughter to help with bathing and other hygiene tasks several times a week. |
| 28 | 63 m, ipd 5 years, tx levocarb 100/25 2 tab tid, rasagiline 1 mg qd. His handwriting has become very small and cramped, to the point that it's difficult even for him to read it. When he tries to write a check or sign his name, the letters start out legible but quickly shrink in size and become hard to decipher. He finds it frustrating because he used to have neat handwriting, and now he often relies on typing or having his wife fill out forms for him. |
| 29 | 68 f, ipd 9 years, tx levocarb 100/25 1.5 tab qid q3h, selegiline 5 md bid, citalopram 20 mg qd. She used to enjoy knitting and gardening, but now her hands shake too much to knit, and her balance is too unsteady to work safely in the garden. She also loved painting, but the fine brush control and stamina required are no longer possible for her. This loss of enjoyable activities has affected her mood and sense of identity. |
| 30 | 74 m, ipd 10 years, tx levocarb 100/25 2 tab qid q4h, levocarb CR 200/50 QHS, melatonin 10 mg qhs, Escitalopram 10 mg qd. He finds it nearly impossible to turn over in bed at night. If he wants to shift positions, he has to wake up fully and sometimes even sit upright to reposition himself. |
| 31 | 59 f, ipd 3 years, tx levocarb 100/25 1.5 tab tid. Her primary symptom is a noticeable tremor in her left hand that appears at rest. The tremor improves somewhat after she takes her Parkinson's medication, but never fully goes away, and it worsens whenever she is stressed or anxious. She finds it embarrassing in social situations, often keeping her hand in her pocket to hide the shaking. |
| 32 | 65 m, ipd 6 years, tx levocarb 100/25 1.5 tab qid q4h, safinamide 100 mg qd, myrbetriq 50 mg qd. He has great difficulty rising from low positions — getting out of bed in the morning or lifting himself out of his car seat or a soft couch has become a slow, taxing process. In the mornings, he often needs to roll to the side and push up with his arms to sit at the edge of the bed, then gather strength to stand. When exiting his car, he has to rock back and forth a few times to gain momentum to stand up, and often requires someone to hold the door or give him a hand to pull him out of a deep chair. |
| 33 | 72 f, ipd 10 years, tx levocarb 100/25 2 tab qid q4h, entacapone 200 mg qid, rasagiline 1 mg qd. Her walking has become very impaired: she takes short, shuffling steps and has a stooped posture. She frequently has near-falls if she turns too |

| | |
|---|---|
| | quickly or if someone bumps into her, so she has to concentrate hard on every step to avoid stumbling. She now uses a walker for safety. |
| 34 | 69 m, ipd 9 years, tx levocarb 100/25 2 tab qid q4h, entacapone 200 mg qid. no history of cognitive problems. no history of hallucinations. He experiences episodes where his feet feel suddenly glued to the floor. This often happens when he's approaching a doorway or about to initiate walking, and it can last several seconds during which he simply cannot take a step despite trying. During these episodes, he is afraid he will fall because his upper body might lurch forward while his feet stay stuck. |
| 35 | 67 m, ipd 6 years, tx levocarb 100/25 2 tab qid q4h, safinamide 100 mg qd. Reports irregular, involuntary movements of the limbs and trunk occurring about 45–60 minutes after each levocarb dose. Movements are more noticeable in the afternoon. He remains functionally independent. In general, the patient's wife notices these movements more than he does himself. |
| 36 | 72 f, ipd 9 years, tx levocarb CR 200/50 qhs + levocarb 100/25 1.5 tab qid q4h, rasagiline 1 mg qd. Describes frequent lightheadedness and near-fainting when getting up in the morning or after prolonged sitting. No chest pain or palpitations. Blood pressure drop documented from 130/80 supine to 95/60 standing. |
| 37 | 70 m, ipd 8 years, tx prolopa 200mg 6 times a day (7-10-13-16-19-22), safinamide 100 mg qd. myrbetriq 50 mg qd, citalopram 20 mg qd. Reports seeing "small animals" or "children running" in the evenings while watching television. He recognizes they are not real and finds them undisturbing. |
| 38 | 68 f, ipd 1 year, tx stalevo 100 mg tid q4h . Complains of persistent loose stools and mild abdominal cramps for several weeks since recently starting stalevo. Denies blood or mucus. No other systemic symptoms. |
| 39 | 65 m, ipd 5 years, tx levocarb 100/25 1.5 tab qid q4h + rasagiline 1 mg qd. Reports an episode of severe headache, flushing, and elevated blood pressure (190/100 mmHg) shortly after eating aged cheese at a family event. No recurrent episodes since dietary modification. |
| 40 | 71 f, ipd 7 years, tx levocarb 100/25 1.5 tab qid q4h + selegiline 5 mg bid. Since starting selegiline, she reports difficulty falling asleep and frequent nighttime awakenings. No vivid dreams or anxiety. Daytime fatigue increased. |
| 41 | 63 m, ipd 7 years, tx levocarb 100/25 2 tab at 7-10-13-16-19-22, Sinemet CR 200/50 1 tab QHS, safinamide 100 mg qd. New episodes have emerged in which the patient reports seeing animals quickly passing in front of him. He recognizes that these are not real, and they do not cause him fear or distress. |
| 42 | 69 f, ipd 9 years, tx levocarb 100/25 2.5 tab qid q4h + entacapone 200 mg qid. Recently noticed that her urine turned dark orange but denies dysuria, frequency, or pain. Laboratory urinalysis normal. No other symptoms. |
| 43 | 62 m, ipd 3 years, tx levocarb 100/25 1 tab qid q4h + pramipexole 0.75 mg tid. His wife reports that he has fallen asleep suddenly during daytime activities, including while sitting at the table. denies prior warning or drowsiness. |
| 44 | 70 f, ipd 6 years, tx levocarb 100/25 2 tab qid q4h + ropinirole 4 mg tid. The patient recently replaced his car with a luxury vehicle beyond his financial means, using most of his savings to do so. He did not consult his wife beforehand and describes the decision as an impulsive act, stating that he "just felt like doing it." |
| 45 | 61 f, ipd 4 years, tx levocarb 100/25 1 tab qid q4h + rotigotine patch 4 mg qd. Reports redness, itching, and mild peeling of skin at the patch site after two weeks of use. No systemic rash or allergy history. |

| | |
|---|---|
| 46 | 78 f, ipd 10 years , tx levocarb 100/25 1 tab qid q4h + rivastigmine 6 mg bid. Since titration to current dose, she reports daily nausea and reduced appetite with a 2 kg weight loss. No vomiting. |
| 47 | 79 m, ipd 11 years , tx levocarb 100/25 2.5 tab qid q4h + donepezil 10 mg qd. His wife reports that he now experiences vivid and disturbing dreams almost every night. Sleep is fragmented, and he wakes up anxious. |
| 48 | 80 f, ipd 12 years , tx memantine 10 mg bid + levocarb 100/25 2.5 tab qid q4h. Family reports new-onset confusion and irritability after dose escalation from 10 to 20 mg daily. No infection or metabolic cause identified. |
| 49 | 69 m, ipd 8 years, tx amantadine 100 mg bid + levocarb 100/25 2 tab qid q4h. Reports bilateral leg swelling . No shortness of breath or cardiac history. |
| 50 | 62 f, ipd 7 years, tx levocarb 100/25 1.5 tab qid, entacapone 200 mg QID. She experiences clear wearing-off symptoms about 30–45 minutes before her next LEVOCARB dose is due. Her hands begin to tremble, her legs stiffen, and she feels anxious and fatigued. These motor and non-motor symptoms resolve shortly after taking the next dose, but the fluctuation disrupts her ability to plan activities and leaves her feeling vulnerable and dependent on the clock. |

*Supp Table 1: List of the clinical vignettes that were used with the LLM's.*

### Supp. Table 2- Likert Scale for Assessment rating:

| **Scale** | **Meaning** |
|---|---|
| 1 – Completely Incorrect | A clinical impression that does not match the case at all, contains an incorrect diagnosis, or ignores the main clinical problem. |
| 2 – Partially / Significantly Incorrect | Touches only a small portion of the problem, misinterprets important elements, or misses the core issue. |
| 3 – Too General / Acceptable but Not Focused | A general or partially correct impression, but lacking detail or failing to distinguish between similar clinical possibilities. |
| 4 – Good / Mostly Accurate | A clinically correct impression that identifies the primary issue but may lack specificity or subtyping. |
| 5 – Completely Accurate (Expert-Level) | A precise, focused, and clinically accurate definition of the primary issue relevant to the case. |

### Supp. Table 3- Likert Scale for Plan rating:

| **Scale** | **Meaning** |
|---|---|
| 1 – Unsafe / Incorrect / Dangerous | A recommendation that is medically unsound, potentially harmful, or contradicts clear clinical guidelines. |
| 2 – Inappropriate (Overly Aggressive or Overly Conservative) | A plan that shows poor clinical judgment by being excessively aggressive or too conservative to the point of missing appropriate treatment. |
| 3 – Acceptable but Suboptimal | Safe recommendations but not ideal; may be too general, insufficiently tailored, or |

| | |
|---|---|
| | missing a better evidence-based alternative. |
| 4 – Good and Clinically Appropriate | A correct and evidence based plan that fits the case well, with no major safety concerns, though not fully optimized. |
| 5 – Optimal, Precise, Evidence-Based (Expert-Level) | A fully appropriate, clear, safe, and optimal recommendation that aligns perfectly with the case details. |

### Supp. Table 4- Gemini response for each case:

| case number | Gemini response (2025-11-17) |
|---|---|
| 1 | A: The most likely diagnosis is Levodopa-Induced Dyskinesia (LID), characterized by involuntary movements fluctuating with Levodopa dosing.<br><br>P: Initiate Amantadine extended-release 274 mg PO daily to reduce LID severity by modulating glutamatergic pathways, while maintaining the current levocarb regimen. |
| 2 | A. The patient is experiencing motor and non-motor "wearing off" phenomena, which is a common complication of long-term Levodopa therapy in Parkinson's disease.<br><br>P. To stabilize Levodopa plasma levels and reduce "off" time, increase the frequency of immediate-release levodopa doses. Levocarb 100/25 1.5 tab per dose, administered 5 times a day (q4h wake time). |
| 3 | A: The patient is experiencing delayed onset of action of immediate-release levodopa, which is most likely due to food interference with absorption.<br><br>P: The most appropriate first-line intervention is to optimize medication timing by instructing the patient to take the immediate-release levocarb on an empty stomach: Levocap 100/25 two tablets by mouth four times daily, 60 minutes before meals. |
| 4 | A. The most likely clinical impression is refractory delayed or failed motor response (delayed or absent ON) to levodopa in a patient with fluctuating Parkinson's disease.<br><br>P. Add opicapone to the current regimen to enhance the beneficial effects of levodopa and reduce OFF time. Opicapone 50 mg PO once daily. |

| | |
|---|---|
| 5 | A. The patient is experiencing severe morning off symptoms, a common motor complication of Parkinson's disease, likely due to the overnight wearing off of levodopa/carbidopa.<br>P. Increase the dosing frequency of the current levodopa/carbidopa regimen to ensure consistent drug levels, which can be achieved by adding an early morning dose or adjusting the timing of the first dose. Rx: levodopa/carbidopa 100/25 mg, 1 tablet, oral, five times daily (q4h wakeful hours), with the last dose close to bedtime. |
| 6 | A: The most likely diagnosis is Parkinson's disease with inadequate control of rest tremor despite optimized levodopa/carbidopa dosing.<br>P: Given the poor response of tremor to levocarb, a monoamine oxidase type B (MAO-B) inhibitor should be added to the regimen to potentiate the dopaminergic effect. Rx: Selegiline 5 mg, PO, twice daily. |
| 7 | A. The most likely clinical impression is Parkinson’s disease of 1 year duration complicated by gait instability and falls, potentially related to inadequate levodopa dosing or motor fluctuations.<br><br>P. Increase the current Levocarb regimen to improve motor control: Carbidopa-Levodopa 25-100 mg, oral, five times daily (one tab 30 min before each of 4 meals/snacks during the day and one at bedtime) to increase 'on' time and stability. |
| 8 | A: The patient, a 68-year-old with established idiopathic Parkinson's disease, is presenting with advanced motor symptoms characterized by significant gait impairment, specifically festination and instability, putting her at high risk for falls despite triple therapy.<br><br>P: Initiate a structured physical therapy program focusing on gait training with visual cueing techniques, which is the most effective rehabilitation strategy to mitigate festination and improve stability. |
| 9 | A. The most likely clinical impression is Parkinson’s Disease Dementia (PDD), given the significant decline in multiple cognitive domains occurring 12 years after the initial diagnosis of Parkinson's disease.<br><br>P. Initiate treatment with rivastigmine to address the cognitive impairment: rivastigmine tartrate 1.5 mg PO BID, titrating up as tolerated every 4 weeks to a target dose of 6 mg PO BID. |
| 10 | A: The most likely diagnosis is Parkinson's Disease Psychosis (PDP), presenting as visual hallucinations with preserved insight, a common non-motor complication of Parkinson's disease.<br><br>P: Initiate treatment with the first FDA-approved medication for this condition to manage psychotic symptoms without worsening motor control: Pimavanserin 34 mg PO once daily. Continue current Parkinson's disease medications but closely |

| | |
|---|---|
| | monitor for any potential drug interactions or adverse effects, especially increased anxiety. |
| 11 | A. The patient is suffering from major depressive disorder refractory to selective serotonin reuptake inhibitor (SSRI) therapy in the context of Parkinson’s disease (PD).<br><br>P. Augment the treatment plan by switching the ineffective SSRI to a serotonin-norepinephrine reuptake inhibitor (SNRI) to target the persistent depressive symptoms. Prescribe venlafaxine 37.5 mg PO daily. |
| 12 | A. The patient is exhibiting clear signs of Levodopa wearing-off, manifesting as motor fluctuations (shaky, sweaty, panic-stricken) and non-motor symptoms (anxiety, feeling on edge) prior to his next medication dose.<br><br>P. Increase the frequency of levodopa/carbidopa dosing to better manage motor and non-motor fluctuations: Levodopa/carbidopa 100/25 mg 1 tablet PO Q3h during waking hours. |
| 13 | A. The most likely clinical impression is apathy, a common non-motor symptom in Parkinson’s disease, characterized by reduced motivation and goal-directed behavior.<br><br>P. Given the current complex medication regimen, a behavioral approach should be implemented, specifically creating and adhering to a daily schedule of activities and exercise to manage the apathy. |
| 14 | A: The most likely clinical impression is a Levodopa-Induced Impulse Control Disorder (ICD), presenting as disinhibition and problematic hypersexuality, exacerbated by the patient's pattern of levocarb overuse.<br><br>P: The primary treatment strategy is to reduce the patient's total dopaminergic load by lowering the dose of the most likely causative agent.<br><br>P: Adjust LEVOCARB (Carbidopa/Levodopa) 100/25 mg to 1.5 tablets PO QID Q4H, while closely monitoring for motor worsening and resolution of the ICD symptoms. |
| 15 | A: The most likely diagnosis is Insomnia/Fragmented Sleep, a common non-motor symptom of Parkinson's disease, with potential contributions from levodopa-related sleep disturbances or nocturnal motor symptoms.<br><br>P: Initiate a short course of a non-benzodiazepine sedative hypnotic to improve sleep maintenance. Rx: Zolpidem 5 mg PO QHS PRN, ensuring no significant nocturnal motor symptoms (like tremor or difficulty turning) are the primary cause of awakening before escalating therapy. |

| | |
|---|---|
| 16 | A. The most likely clinical impression is excessive daytime sleepiness (EDS) secondary to Parkinson's disease, significantly impacting daily function and safety.<br><br>P. Initiate treatment for EDS with a psychostimulant that has evidence in PD. Modafinil 100 mg PO BID. |
| 17 | A: The patient is experiencing musculoskeletal pain, a common non-motor symptom of Parkinson's disease, which appears related to motor fluctuations or "off" periods given its correlation with pronounced PD symptoms.<br><br>P: The primary treatment strategy should be optimizing the dopaminergic therapy to reduce "off" periods; increase the Levodopa/Carbidopa dose to 2 tablets by mouth three times daily (Levocarb 100/25 tab ii tid) to potentially improve muscle rigidity and associated pain. |
| 18 | A: The patient, who has established idiopathic Parkinson's disease (IPD), is experiencing significant, disruptive urinary urgency, frequency, and nocturia, which is a common non-motor symptom of Parkinson's disease, likely due to detrusor overactivity.<br><br>P: Initiate a selective anticholinergic medication to address the overactive bladder symptoms. Prescribe solifenacin 5 mg PO daily. |
| 19 | A: This patient presents with refractory chronic constipation, a common and potentially debilitating non-motor symptom of Parkinson's disease.<br><br>P: Increase the dose of the existing osmotic laxative, Senna, to address the refractory constipation, or switch to a high-dose polyethylene glycol (PEG) 3350 regimen, which is a safe and effective first-line treatment for chronic constipation in PD.<br><br>I am not sure. The two options I am considering are:1. Increase senna to 3 tabs qhs.<br>2. Switch to PEG 3350 17g (capful) PO daily. |
| 20 | A: This patient presents with symptoms highly suggestive of neurogenic orthostatic hypotension (nOH) secondary to Parkinson's disease, a common non-motor complication.<br>P: Initial management should prioritize non-pharmacological interventions, such as increasing salt and fluid intake and using compression stockings; if symptoms persist, initiate midodrine 5 mg by mouth three times daily. |
| 21 | A: The patient is experiencing fatigue, a common non-motor symptom of idiopathic Parkinson’s disease, despite current dopaminergic and adjunct therapies.<br><br>P: Implement a focused physical therapy program encouraging regular exercise, starting slowly and building up to at least 2.5 hours per week, which is the best |

| | |
|---|---|
| | known non-pharmacological therapy for improving PD-related fatigue and overall well-being. |
| 22 | A<br>The most likely clinical impression is Parkinson's disease complicated by severe hypophonia/dysphonia (speech difficulty) that is functionally limiting.<br>P<br>The recommended therapeutic strategy is referral for specialized speech therapy, specifically the Lee Silverman Voice Treatment (LSVT LOUD) program. This highly-intensive, evidence-based voice therapy focuses on increasing vocal loudness and is the gold standard for treating hypophonia in Parkinson's disease. |
| 23 | A: This patient presents with sialorrhea (drooling) secondary to Idiopathic Parkinson's Disease, which is unrelated to fluctuating motor symptoms or medication wearing off.<br><br>P: I recommend initiating an oral anticholinergic agent to reduce saliva production. The treatment plan is to start glycopyrrolate 1 mg PO TID, titrating carefully based on response and tolerability, particularly for potential side effects such as dry mouth or cognitive impairment. |
| 24 | A. The most likely clinical impression is advanced Parkinson’s disease complicated by oropharyngeal dysphagia.<br>P. Address the dysphagia with a referral for a comprehensive speech-language pathology (SLP) swallowing evaluation and initiate physical therapy focused on swallowing exercises. Continue current Parkinson's disease medications as they address motor symptoms, but prioritize non-pharmacological interventions for this specific complication. |
| 25 | A: The patient is likely experiencing advanced idiopathic Parkinson's disease with significant motor fluctuations and tremor leading to severe impairment in activities of daily living (eating).<br>P: Given the duration of disease, existing optimized medical therapy (levocarb/entacapone), and severe functional impairment, Deep Brain Stimulation (DBS) should be considered as the definitive, evidence-based therapeutic strategy to reduce motor complications and improve quality of life. |
| 26 | A. The patient is a 69-year-old male with confirmed idiopathic Parkinson's disease of 8 years duration, currently experiencing motor fluctuations ("off" periods) manifested as significant bradykinesia and difficulty with balance during self-care activities, despite current treatment with carbidopa-levodopa and a monoamine oxidase B inhibitor.<br>P. To address the functional decline and wearing-off symptoms, optimize the existing regimen by increasing the frequency of carbidopa-levodopa, aiming for more consistent dopaminergic stimulation throughout the day: increase Levocarb 100/25 tab PO to 2 tabs five times daily (q3h while awake). |

| | |
|---|---|
| 27 | I am not sure, considering either Deep Brain Stimulation (DBS) or a Levodopa-Carbidopa Intestinal Gel (LCIG) infusion. |
| 28 | A: The most likely clinical impression is Parkinson's disease, characterized by micrographia, which is a common motor symptom.<br>P: Increase the dosage of the current levodopa/carbidopa regimen to optimize symptomatic control of motor features including micrographia. Prescription: levocarb 100/25 two tabs qid, PO. |
| 29 | A. The most likely clinical impression is a 68-year-old female with Idiopathic Parkinson's Disease (IPD) experiencing inadequate motor symptom control, leading to functional decline and secondary mood symptoms.<br><br>P. Increase the levodopa-carbidopa dosage to optimize motor symptom control: levocarb 100/25 2 tablets PO qid q3h. |
| 30 | **A** The most likely diagnosis is nocturnal hypokinesia/off-periods, consistent with motor fluctuations in advanced Parkinson's disease.<br>**P** To specifically address nighttime "off" periods and improve nocturnal mobility, administer a dopamine agonist patch. Apply a Rotigotine transdermal patch (Rotigotine 4 mg/24h patch) once daily (qd). |
| 31 | A: The clinical impression is Parkinson's disease with fluctuating motor symptoms ("off" periods) despite current levodopa therapy.<br><br>P: Adjust the current levocarb dose to optimize motor control and specifically address the persistent "off" period tremor; prescribe carbidopa/levodopa 25/100 mg, take 2 tablets by mouth three times daily. |
| 32 | A. This patient is likely experiencing motor "off" periods, manifesting as severe levodopa-responsive bradykinesia and difficulty with transitions/axial movements, despite being on a complex dopaminergic regimen.<br>P. Increase the frequency of levocarb to maximize "on" time and reduce specific "off" period motor symptoms: levocarb 100/25 tab q4h (total 5 doses) PO. |
| 33 | A. The most likely clinical impression is advanced Parkinson's disease with refractory gait failure and postural instability despite optimized medical management.<br><br>P. Given the inadequate response to maximal medical therapy and the patient's gait impairment, a definitive treatment plan is to evaluate the patient for Deep Brain Stimulation (DBS) therapy to improve motor fluctuations and gait symptoms. |
| 34 | **A** The patient presents with classic Freezing of Gait (FOG), a common motor complication of advanced idiopathic Parkinson’s disease (IPD) likely related to inadequate dopaminergic coverage or motor fluctuations.<br><br>**P** Initiate Pramipexole 0.375 mg extended-release tablet PO once daily, |

| | |
|---|---|
| | titrating slowly every 5–7 days as tolerated, to maximize "on" time and reduce FOG episodes by providing continuous dopaminergic stimulation. Non-pharmacological management, including physical therapy incorporating visual and auditory cueing techniques, should also be strongly recommended. |
| 35 | **A** The patient is most likely experiencing levodopa-induced dyskinesia (LID), specifically peak-dose dyskinesia, based on the timing of irregular, involuntary movements 45–60 minutes after each levocarb dose.<br>**P** Reduce the current levocarb dose by 25% to minimize peak plasma concentration and dyskinesia, changing the prescription to levocarb 75/18.75 mg PO QID Q4H, while maintaining safinamide therapy. |
| 36 | A: The most likely diagnosis is symptomatic orthostatic hypotension, defined by a significant drop in blood pressure upon standing, in a patient with Parkinson's disease.<br>P: Initiate treatment with midodrine to manage the orthostatic symptoms. Midodrine 5 mg po tid at approximately 4-hour intervals during daytime hours (not later than 6 p.m.). |
| 37 | A. The patient is experiencing minor, non-disturbing visual hallucinations (likely visual misperceptions) as a symptom of Parkinson's Disease Psychosis (PDP), which is common in advanced PD and may be exacerbated by existing dopaminergic therapy and citalopram.<br><br>P. Given the hallucinations are non-disturbing and the patient is on high-dose levodopa, the initial evidence-based strategy is to safely reduce the dose of non-PD medications that may contribute, specifically tapering citalopram 20 mg PO daily. |
| 38 | **A** The patient is experiencing persistent non-bloody diarrhea and abdominal cramps highly consistent with a common gastrointestinal side effect of the entacapone component in Stalevo.<br>**P** Discontinue Stalevo immediately and transition to carbidopa/levodopa 100/25 mg PO tid q4h, as entacapone-related diarrhea necessitates removal of the COMT inhibitor. If motor fluctuations recur, consider adding Opicapone 50 mg PO once daily at bedtime. |
| 39 | A. The patient experienced a tyramine-induced hypertensive crisis precipitated by aged cheese due to concomitant treatment with the MAO-B inhibitor rasagiline.<br>P. The definitive plan is to discontinue rasagiline (1 mg qd) immediately to eliminate the risk of recurrent hypertensive crises; continue Levodopa/Carbidopa (100/25 mg, 1.5 tab QID Q4h) to maintain Parkinson's disease motor control. |
| 40 | **A** The patient is experiencing selegiline-induced insomnia, manifesting as difficulty falling asleep and frequent nighttime awakenings, leading to increased daytime fatigue.<br><br>**P** To address the sleep disturbance likely caused by selegiline, discontinue |

| | |
|---|---|
| | selegiline 5 mg bid and initiate a short-acting non-benzodiazepine for sleep-onset insomnia: zolpidem 5 mg PO qhs PRN. |
| 41 | A. The patient is experiencing minor visual hallucinations, a form of Parkinson's disease psychosis (PDP) with retained insight, which is often an early sign of progressing cognitive burden in PD.<br>P. Given the patient is on multiple dopaminergic agents, the first-line treatment is to cautiously reduce the non-carbidopa/levodopa dopaminergic medications; therefore, discontinue safinamide 100 mg qd. |
| 42 | A: The most likely clinical impression is a benign side effect of entacapone causing urine discoloration in a patient with idiopathic Parkinson's disease.<br>P: The patient should be reassured that the dark orange urine is a common and harmless side effect of entacapone, requiring no change in the current antiparkinsonian regimen. Continue levocarb 100/25 2.5 tab qid q4h and entacapone 200 mg qid PO. |
| 43 | A: The most likely clinical impression is sudden-onset sleep attacks, a known adverse effect of the dopamine agonist pramipexole in a patient with Parkinson's disease.<br><br>P: To manage the dangerous side effect of sudden sleep attacks, reduce the causative agent: Pramipexole (Mirapex) 0.375 mg PO TID. If motor symptoms worsen, consider adding Modafinil 100 mg PO daily. |
| 44 | A: The most likely diagnosis is impulse control disorder, specifically pathological spending, induced by dopamine agonist therapy (ropinirole) in a patient with Parkinson's disease.<br><br>P: Discontinue ropinirole 4 mg tid immediately and manage any withdrawal symptoms; levocarb should be continued at the current dose. |
| 45 | A. The most likely clinical impression is a common application site reaction related to the rotigotine transdermal system in a patient with Parkinson's disease.<br>P. Discontinue the current rotigotine transdermal patch due to the skin irritation and switch the patient to an equivalent dose of an oral nonergot dopamine agonist. The new therapeutic strategy is to initiate ropinirole 0.5 mg PO TID and titrate weekly as tolerated to maintain dopamine agonist effect. |
| 46 | A. The most likely clinical impression is gastrointestinal intolerance (nausea, reduced appetite, and weight loss) secondary to the recent titration of rivastigmine therapy.<br><br>P. To mitigate the adverse effects, decrease the current dose of the suspected causative agent; reduce rivastigmine oral capsule to 3 mg twice daily (BID) and monitor clinical status and weight. |
| 47 | A. The most likely clinical impression is REM Sleep Behavior Disorder (RBD) in the setting of long-standing Parkinson's disease, potentially exacerbated by existing |

| | |
|---|---|
| | dopaminergic therapy.<br>P. The treatment plan is to initiate clonazepam 0.5 mg PO at bedtime to manage the dream-related sleep disturbance and anxiety. The dose can be titrated based on clinical response, while monitoring for daytime sedation and balance. |
| 48 | **A** The most likely clinical impression is drug-induced confusion and irritability secondary to memantine dose escalation in an elderly patient with Parkinson's disease.<br>**P** The treatment plan is to immediately discontinue the recently increased dose of memantine and return to the prior dose to resolve the acute neurobehavioral symptoms. Reduce memantine to 10 mg PO BID, then consider gradual titration of levocarb 100/25 tab to manage motor symptoms if needed after symptoms resolve. |
| 49 | **A** The patient most likely has peripheral edema secondary to amantadine therapy.<br>**P** Discontinue amantadine 100 mg bid, and monitor for resolution of bilateral leg swelling. |
| 50 | A. The patient is experiencing end-of-dose "wearing-off" motor fluctuations, a common complication of long-term Levodopa therapy in Parkinson's disease, despite being treated with a COMT inhibitor (entacapone).<br>P. To address the predictable wearing-off, increase the frequency of the levocarb dosing: LEVOCARB (Carbidopa/Levodopa) 100/25 mg, 1.5 tab, PO, five times daily (instead of qid). |

*Supp Table 2: Gemini response for each case*

### Supp. Table 5- OpenEvidence response for each case

| case number | openevidance response (run 8/12/2025) | openevidance response shortened to A+P only for faster reading convenience (shown to raters) |
|---|---|---|
| 1 | This assessment and plan addresses peak-dose dyskinesia in a patient with established Parkinson's disease on levodopa therapy. The evidence strongly supports amantadine as the first-line pharmacological treatment for levodopa-induced dyskinesia, with extended-release formulations offering improved tolerability and once-daily dosing.[1-5]<br><br>Assessment and Plan<br><br>Assessment: | A: The patient exhibits involuntary limb movements occurring approximately one hour after levodopa dosing, consistent with peak-dose dyskinesia—the most common pattern of levodopa-induced dyskinesia.[6] The movements are non-disabling and do not interfere with function, though they cause caregiver distress. The temporal relationship to levodopa administration and the choreiform nature of the movements support this diagnosis.<br>P: Add amantadine extended-release 274 mg once daily at bedtime. |

| | | |
|---|---|---|
| | 65-year-old male with idiopathic Parkinson's disease (5-year duration) presenting with levodopa-induced peak-dose dyskinesia. Currently treated with levodopa/carbidopa 100/25 mg, 2 tablets four times daily (total daily dose: 800 mg levodopa/200 mg carbidopa), and safinamide 100 mg daily.<br><br>The patient exhibits involuntary limb movements occurring approximately one hour after levodopa dosing, consistent with peak-dose dyskinesia—the most common pattern of levodopa-induced dyskinesia.[6] The movements are non-disabling and do not interfere with function, though they cause caregiver distress. The temporal relationship to levodopa administration and the choreiform nature of the movements support this diagnosis.<br><br>Plan:<br><br>Add amantadine extended-release 274 mg once daily at bedtime.<br><br>Amantadine is the only FDA-approved medication with established efficacy for levodopa-induced dyskinesia.[2-3][5] A recent network meta-analysis of 85 randomized controlled trials identified immediate-release amantadine as the highest-ranking oral medication for dyskinesia reduction (SUCRA = 86.5%).[1] Extended-release formulations provide superior pharmacokinetics with once-daily evening dosing, offering higher and more continuous plasma levels compared to immediate-release formulations that require up to three times daily administration.[4-5]<br><br>The extended-release formulation has demonstrated efficacy in reducing dyskinesia and "off" time in pivotal trials.[4] Two extended-release | Amantadine is the only FDA-approved medication with established efficacy for levodopa-induced dyskinesia.[2-3][5] A recent network meta-analysis of 85 randomized controlled trials identified immediate-release amantadine as the highest-ranking oral medication for dyskinesia reduction (SUCRA = 86.5%).[1] Extended-release formulations provide superior pharmacokinetics with once-daily evening dosing, offering higher and more continuous plasma levels compared to immediate-release formulations that require up to three times daily administration.[4-5]<br>The extended-release formulation has demonstrated efficacy in reducing dyskinesia and "off" time in pivotal trials.[4] Two extended-release preparations are FDA-approved for this indication.[5] The recommended starting dose is 274 mg once daily at bedtime, which can be titrated based on response and tolerability. |

| | |
|---|---|
| | preparations are FDA-approved for this indication.[5] The recommended starting dose is 274 mg once daily at bedtime, which can be titrated based on response and tolerability.<br><br>Rationale for this approach:<br><br>Amantadine's anti-dyskinetic effects are mediated through N-methyl-D-aspartate (NMDA) receptor antagonism, addressing glutamate overactivity implicated in dyskinesia pathophysiology[7-8]<br><br>The patient's dyskinesias are mild and non-disabling, making pharmacological augmentation preferable to levodopa dose reduction, which could worsen parkinsonian symptoms<br><br>The current levodopa regimen provides adequate motor control, and the addition of safinamide suggests optimization of dopaminergic therapy has been attempted<br><br>Monitoring and follow-up:<br><br>Assess dyskinesia severity and frequency at 4-6 week follow-up<br><br>Monitor for potential adverse effects including peripheral edema, livedo reticularis, hallucinations, and confusion[2]<br><br>Caution regarding potential worsening of visual hallucinations with amantadine[2]<br><br>If dyskinesias worsen or become functionally limiting despite amantadine, consider neurosurgical consultation for deep brain stimulation evaluation, as GPi-DBS demonstrates the highest efficacy for dyskinesia reduction in advanced cases[1]<br><br>Patient and caregiver education: |

| | | |
|---|---|---|
| | Explain that mild dyskinesias are a common consequence of long-term levodopa therapy<br><br>Reassure that treatment aims to reduce movements while maintaining motor function<br><br>Discuss realistic expectations: amantadine provides moderate anti-dyskinetic benefit rather than complete elimination | |
| 2 | The provided references focus on evidence-based medicine methodology and do not contain specific clinical guidance for Parkinson's disease management. Therefore, this assessment and plan will be based on standard neurological practice for managing motor fluctuations in advanced Parkinson's disease, without citations from the provided sources.<br><br>Assessment and Plan<br><br>Assessment:<br><br>66-year-old female with 7-year history of idiopathic Parkinson's disease presenting with end-of-dose wearing-off phenomenon characterized by recurrent right hand tremor, malaise with anxiety, and freezing of gait occurring approximately 40 minutes before scheduled levodopa/carbidopa doses.<br><br>Current medication regimen:<br><br>Levodopa/carbidopa 100/25 mg, 1.5 tablets (150/37.5 mg) four times daily every 4 hours<br><br>Levodopa/carbidopa controlled release 200/50 mg at bedtime<br><br>Rasagiline 1 mg once daily | Assessment: The patient is experiencing classic motor fluctuations with predictable wearing-off despite being on combination therapy with levodopa, a MAO-B inhibitor (rasagiline), and a COMT inhibitor (entacapone). The symptoms occurring 40 minutes before the next dose suggest inadequate duration of levodopa effect despite the use of entacapone to prolong levodopa availability.<br><br>Plan: Add amantadine immediate-release 100 mg three times daily (morning, midday, and early evening). Rationale: Amantadine provides additional benefit for motor fluctuations and wearing-off symptoms through NMDA receptor antagonism. It can reduce off-time and may also help with dyskinesias if they develop. The three-times-daily dosing will provide coverage throughout the waking hours when wearing-off is problematic. Titration: Start with 100 mg once daily for 1 week, then increase to 100 mg twice daily for 1 week, then advance to the target dose of 100 mg three times daily. The last dose should be given no later than early evening to minimize insomnia risk. Monitoring: Reassess in 4-6 weeks for improvement in wearing- |

| | | |
|---|---|---|
| | Entacapone 200 mg four times daily<br><br>Mirabegron 50 mg once daily (for urinary symptoms)<br><br>Polyethylene glycol 17 g once daily (for constipation)<br><br>The patient is experiencing classic motor fluctuations with predictable wearing-off despite being on combination therapy with levodopa, a MAO-B inhibitor (rasagiline), and a COMT inhibitor (entacapone). The symptoms occurring 40 minutes before the next dose suggest inadequate duration of levodopa effect despite the use of entacapone to prolong levodopa availability.<br><br>Plan:<br><br>Add amantadine immediate-release 100 mg three times daily (morning, midday, and early evening).<br><br>Rationale: Amantadine provides additional benefit for motor fluctuations and wearing-off symptoms through NMDA receptor antagonism. It can reduce off-time and may also help with dyskinesias if they develop. The three-times-daily dosing will provide coverage throughout the waking hours when wearing-off is problematic.<br><br>Titration: Start with 100 mg once daily for 1 week, then increase to 100 mg twice daily for 1 week, then advance to the target dose of 100 mg three times daily. The last dose should be given no later than early evening to minimize insomnia risk.<br><br>Monitoring: Reassess in 4-6 weeks for improvement in wearing-off symptoms, freezing of gait, and anxiety associated | off symptoms, freezing of gait, and anxiety associated with off periods. Monitor for potential side effects including livedo reticularis, peripheral edema, hallucinations, and confusion. |

| | | |
|---|---|---|
| | with off periods. Monitor for potential side effects including livedo reticularis, peripheral edema, hallucinations, and confusion.<br><br>Alternative considerations if inadequate response: Consider referral to movement disorders specialist for evaluation of advanced therapies including deep brain stimulation, continuous levodopa-carbidopa intestinal gel, or subcutaneous apomorphine, given 7-year disease duration with motor fluctuations despite optimized oral therapy.<br><br>Continue current doses of all other medications unchanged at this time.<br><br>This assessment and plan addresses the patient's wearing-off phenomenon with a specific medication recommendation and dosing schedule. The treatment approach follows standard neurological practice for managing motor fluctuations in Parkinson's disease, though the provided references did not contain Parkinson's-specific clinical evidence.<br><br>Would you like me to review the latest evidence on the efficacy and safety of amantadine for motor fluctuations and freezing of gait in Parkinson's disease, particularly in patients already on levodopa, MAO-B, and COMT inhibitors? This could help clarify its expected benefits and risks in this specific clinical context. | |
| 3 | This assessment and plan will address the delayed levodopa response ("delayed-on") phenomenon in this patient with advanced Parkinson's disease. While the provided references focus on chronic disease self-management and other conditions rather than Parkinson's disease specifically, the clinical approach will draw on established principles of Parkinson's disease management, | Assessment: 75ב-year-old male with 10-year history of idiopathic Parkinson's disease experiencing delayed-on phenomenon characterized by prolonged time to levodopa effect (>1 hour) occurring randomly throughout the day. Current regimen includes levodopa/carbidopa 100/25 mg 2 tablets QID, levodopa/carbidopa CR 200/50 mg QHS, rasagiline 1 mg |

| | | |
|---|---|---|
| | particularly addressing medication absorption issues that commonly occur with delayed gastric emptying.<br><br>Assessment and Plan<br><br>Assessment:<br><br>75-year-old male with 10-year history of idiopathic Parkinson's disease experiencing delayed-on phenomenon characterized by prolonged time to levodopa effect (>1 hour) occurring randomly throughout the day. Current regimen includes levodopa/carbidopa 100/25 mg 2 tablets QID, levodopa/carbidopa CR 200/50 mg QHS, rasagiline 1 mg daily, entacapone 200 mg QID, and mirabegron 50 mg daily. Patient appropriately takes levodopa 30 minutes before meals.<br><br>The delayed and erratic levodopa absorption is most likely related to gastroparesis, a common autonomic complication in advanced Parkinson's disease that impairs gastric emptying and medication absorption. The patient is already on optimal timing (30 minutes before meals) and is taking entacapone as a COMT inhibitor to prolong levodopa effect, yet continues to experience unpredictable delays.<br><br>Plan:<br><br>Initiate subcutaneous apomorphine 3 mg as rescue therapy for delayed-on episodes. Apomorphine bypasses the gastrointestinal tract entirely, providing reliable dopaminergic stimulation within 10-20 minutes when oral levodopa fails to take effect in a timely manner. The patient should use apomorphine when levodopa has not produced benefit within 45-60 minutes of administration. | daily, entacapone 200 mg QID, and mirabegron 50 mg daily. Patient appropriately takes levodopa 30 minutes before meals. The delayed and erratic levodopa absorption is most likely related to gastroparesis, a common autonomic complication in advanced Parkinson's disease that impairs gastric emptying and medication absorption. The patient is already on optimal timing (30 minutes before meals) and is taking entacapone as a COMT inhibitor to prolong levodopa effect, yet continues to experience unpredictable delays.<br><br>Plan: Initiate subcutaneous apomorphine 3 mg as rescue therapy for delayed-on episodes. Apomorphine bypasses the gastrointestinal tract entirely, providing reliable dopaminergic stimulation within 10-20 minutes when oral levodopa fails to take effect in a timely manner. The patient should use apomorphine when levodopa has not produced benefit within 45-60 minutes of administration. Prior to initiating apomorphine, the patient must start trimethobenzamide 300 mg three times daily for 3 days to prevent nausea and vomiting. Apomorphine is contraindicated with 5-HT3 antagonists (ondansetron, granisetron) due to risk of profound hypotension.<br>The patient and caregiver will require training on proper subcutaneous injection technique. Starting dose is 2-3 mg subcutaneously, which can be titrated up to 6 mg per dose as needed, with maximum of 5 doses per day (20 mg total daily dose).<br>Continue current oral regimen unchanged, as the issue is absorption variability rather than inadequate total levodopa dosing. Monitor for orthostatic hypotension, hallucinations, |

| | | |
|---|---|---|
| | Prior to initiating apomorphine, the patient must start trimethobenzamide 300 mg three times daily for 3 days to prevent nausea and vomiting. Apomorphine is contraindicated with 5-HT3 antagonists (ondansetron, granisetron) due to risk of profound hypotension.<br><br>The patient and caregiver will require training on proper subcutaneous injection technique. Starting dose is 2-3 mg subcutaneously, which can be titrated up to 6 mg per dose as needed, with maximum of 5 doses per day (20 mg total daily dose).<br><br>Continue current oral regimen unchanged, as the issue is absorption variability rather than inadequate total levodopa dosing. Monitor for orthostatic hypotension, hallucinations, and dyskinesias as potential adverse effects of apomorphine therapy.<br><br>Follow-up in 4 weeks to assess response to apomorphine rescue therapy and evaluate need for further optimization of Parkinson's disease management.<br><br>This assessment and plan provides a specific, evidence-based treatment recommendation for the delayed-on phenomenon using subcutaneous apomorphine as rescue therapy. The recommendation includes precise dosing, necessary premedication with trimethobenzamide, contraindications, and monitoring parameters. This approach addresses the core problem of erratic gastrointestinal absorption while maintaining the current oral regimen.<br><br>Would you like me to review the latest evidence on long-term safety, efficacy, and patient satisfaction with subcutaneous apomorphine rescue | and dyskinesias as potential adverse effects of apomorphine therapy.<br>Follow-up in 4 weeks to assess response to apomorphine rescue therapy and evaluate need for further optimization of Parkinson's disease management. |

| | | |
|---|---|---|
| | therapy in advanced Parkinson’s disease, including real-world data and comparative studies with other rescue strategies? | |
| 4 | This assessment and plan addresses motor fluctuations with dose failures in advanced Parkinson's disease. The evidence supports optimizing COMT inhibition by switching from entacapone to opicapone, which has demonstrated superior efficacy in reducing OFF time, and adding a rescue medication for unpredictable OFF episodes.[1-5]<br><br>Assessment and Plan<br><br>Assessment:<br><br>75-year-old female with 10-year history of idiopathic Parkinson's disease experiencing motor fluctuations with dose failures—specifically, at least one daily episode where levodopa/carbidopa does not produce an ON response despite appropriate timing (30 minutes before meals) and adequate bowel function. She is currently on a complex regimen including levodopa/carbidopa immediate-release 100/25 mg two tablets four times daily, levodopa/carbidopa controlled-release 200/50 mg at bedtime, rasagiline 1 mg daily, and entacapone 200 mg four times daily.<br><br>The patient's dose failures represent unpredictable OFF periods, which differ from predictable end-of-dose wearing off.[3] These unpredictable responses likely reflect inconsistent gastrointestinal absorption of levodopa despite proper timing and normal bowel function.[1] Her current regimen already includes a MAO-B inhibitor (rasagiline) and COMT inhibitor (entacapone), but dose failures persist. | Assessment: The patient's dose failures represent unpredictable OFF periods, which differ from predictable end-of-dose wearing off.[3] These unpredictable responses likely reflect inconsistent gastrointestinal absorption of levodopa despite proper timing and normal bowel function.[1] Her current regimen already includes a MAO-B inhibitor (rasagiline) and COMT inhibitor (entacapone), but dose failures persist.<br><br>Plan: Switch entacapone to opicapone 50 mg once daily at bedtime (taken at least one hour before or after levodopa/carbidopa). Opicapone is a once-daily COMT inhibitor that has demonstrated superior efficacy compared to entacapone in reducing OFF time in patients with motor fluctuations.[1-2] This simplifies the regimen from four daily doses of entacapone to a single bedtime dose while potentially providing better control of motor fluctuations. When initiating opicapone, discontinue entacapone and start opicapone 50 mg once daily. Monitor for increased dyskinesia, which may require reduction of levodopa dose.<br>For the unpredictable dose failures, prescribe apomorphine sublingual film as rescue therapy, starting with 10 mg and titrating in 5 mg increments (maximum 35 mg) to achieve a full ON response.[5] This provides rapid relief (most patients achieve full ON response within 30 minutes) for unpredictable OFF episodes without requiring gastrointestinal absorption.[5-6] The patient should be instructed to hold the film under the |

| | | |
|---|---|---|
| | Plan:<br><br>Switch entacapone to opicapone 50 mg once daily at bedtime (taken at least one hour before or after levodopa/carbidopa). Opicapone is a once-daily COMT inhibitor that has demonstrated superior efficacy compared to entacapone in reducing OFF time in patients with motor fluctuations.[1-2] This simplifies the regimen from four daily doses of entacapone to a single bedtime dose while potentially providing better control of motor fluctuations.<br><br>When initiating opicapone, discontinue entacapone and start opicapone 50 mg once daily. Monitor for increased dyskinesia, which may require reduction of levodopa dose. Continue current doses of levodopa/carbidopa immediate-release (100/25 mg two tablets four times daily) and controlled-release (200/50 mg at bedtime), rasagiline 1 mg daily, mirabegron 50 mg daily, and polyethylene glycol 17 grams daily.<br><br>For the unpredictable dose failures, prescribe apomorphine sublingual film as rescue therapy, starting with 10 mg and titrating in 5 mg increments (maximum 35 mg) to achieve a full ON response.[5] This provides rapid relief (most patients achieve full ON response within 30 minutes) for unpredictable OFF episodes without requiring gastrointestinal absorption.[5-6] The patient should be instructed to hold the film under the tongue for 3 minutes without swallowing and may use up to 5 times daily as needed for OFF episodes.[5] Common side effects include oropharyngeal events (approximately 30% of patients), nausea, and dizziness, which are generally mild to moderate.[5]<br><br>Follow-up in 4-6 weeks to assess | tongue for 3 minutes without swallowing and may use up to 5 times daily as needed for OFF episodes.[5] Common side effects include oropharyngeal events (approximately 30% of patients), nausea, and dizziness, which are generally mild to moderate.[5] Follow-up in 4-6 weeks to assess response to opicapone, evaluate need for levodopa dose adjustments, and titrate apomorphine sublingual film to optimal dose for rescue therapy. |

| | | |
|---|---|---|
| | response to opicapone, evaluate need for levodopa dose adjustments, and titrate apomorphine sublingual film to optimal dose for rescue therapy. | |
| 5 | This assessment and plan addresses early morning OFF symptoms in a patient with advancing Parkinson's disease experiencing motor fluctuations. The treatment recommendation is based on evidence supporting COMT inhibitors, specifically opicapone, as an efficacious adjunct to levodopa for reducing OFF time in patients with motor fluctuations.[1-3]<br><br>Assessment and Plan<br><br>Assessment:<br><br>59-year-old male with idiopathic Parkinson's disease (5-year duration) presenting with early morning OFF symptoms characterized by severe tremors, muscle stiffness, bradykinesia, and general malaise upon waking. Current regimen includes levodopa/carbidopa 100/25 mg four times daily (total daily levodopa dose 400 mg), rasagiline 1 mg daily, and rotigotine patch 3 mg daily, along with mirabegron and polyethylene glycol for non-motor symptoms.<br><br>The patient's symptoms are consistent with nocturnal hypokinesia and early morning OFF episodes, which commonly develop after several years of levodopa therapy due to the medication's short half-life and progressive dopaminergic neuronal degeneration. Despite being on a dopamine agonist (rotigotine) and MAO-B inhibitor (rasagiline), the patient continues to experience significant early morning motor symptoms, indicating inadequate overnight dopaminergic coverage.[1] | Assessment: The patient's symptoms are consistent with nocturnal hypokinesia and early morning OFF episodes, which commonly develop after several years of levodopa therapy due to the medication's short half-life and progressive dopaminergic neuronal degeneration. Despite being on a dopamine agonist (rotigotine) and MAO-B inhibitor (rasagiline), the patient continues to experience significant early morning motor symptoms, indicating inadequate overnight dopaminergic coverage.[1]<br><br>Plan: Add opicapone 50 mg once daily at bedtime as adjunctive therapy to the current levodopa/carbidopa regimen.[2-4] Rationale: Opicapone is a once-daily COMT inhibitor that has been assessed as efficacious for treating motor fluctuations in patients with Parkinson's disease on levodopa therapy. COMT inhibitors enhance levodopa's effect by blocking its peripheral metabolism, thereby increasing its half-life and providing more sustained dopaminergic stimulation. Recent evidence specifically supports the use of COMT inhibitors like opicapone for early morning OFF symptoms in moderate to advanced Parkinson's disease. The once-daily dosing at bedtime is particularly advantageous for addressing nocturnal and early morning symptoms.[1-3][5]<br>The patient is already on a MAO-B inhibitor (rasagiline) and dopamine agonist (rotigotine), both of which are also efficacious for motor fluctuations. However, the persistence of early morning OFF symptoms despite these |

| | | |
|---|---|---|
| | Plan:<br><br>Add opicapone 50 mg once daily at bedtime as adjunctive therapy to the current levodopa/carbidopa regimen.[2-4]<br><br>Rationale: Opicapone is a once-daily COMT inhibitor that has been assessed as efficacious for treating motor fluctuations in patients with Parkinson's disease on levodopa therapy. COMT inhibitors enhance levodopa's effect by blocking its peripheral metabolism, thereby increasing its half-life and providing more sustained dopaminergic stimulation. Recent evidence specifically supports the use of COMT inhibitors like opicapone for early morning OFF symptoms in moderate to advanced Parkinson's disease. The once-daily dosing at bedtime is particularly advantageous for addressing nocturnal and early morning symptoms.[1-3][5]<br><br>The patient is already on a MAO-B inhibitor (rasagiline) and dopamine agonist (rotigotine), both of which are also efficacious for motor fluctuations. However, the persistence of early morning OFF symptoms despite these medications indicates the need for additional adjunctive therapy.[3] The strategy of using low to moderate doses of multiple agents rather than escalating single agents often provides optimal symptom control with fewer side effects.[2]<br><br>Monitoring: Assess response to opicapone after 2-4 weeks, monitoring for reduction in early morning OFF time and improvement in morning motor function. Monitor for potential adverse effects including dyskinesia (which may increase with enhanced levodopa effect), diarrhea, and orthostatic hypotension. If | medications indicates the need for additional adjunctive therapy.[3] The strategy of using low to moderate doses of multiple agents rather than escalating single agents often provides optimal symptom control with fewer side effects.[2] |

| | | |
|---|---|---|
| | dyskinesia emerges, consider reducing individual levodopa doses while maintaining dosing frequency.[1]<br><br>Alternative considerations: If opicapone provides insufficient benefit, other options for early morning OFF include rescue therapies such as inhaled or dispersible levodopa formulations, though these require on-demand administration rather than providing sustained overnight coverage.[5-6]<br><br>Continue current doses of levodopa/carbidopa, rasagiline, rotigotine, mirabegron, and polyethylene glycol. Counsel the patient to take levodopa doses at least 1 hour separated from meals to optimize absorption, particularly as disease advances.[2][7] | |
| 6 | This assessment and plan addresses a patient with tremor-dominant Parkinson's disease showing partial response to levodopa. The evidence indicates that tremor is often less responsive to levodopa than bradykinesia and rigidity, and the current dose escalation strategy may not be optimal.[1] Based on the medical literature supporting adjunctive therapies for motor symptoms and the specific challenge of tremor management, the following clinical note will recommend adding a COMT inhibitor to enhance levodopa efficacy while addressing the tremor-dominant presentation.[1-3]<br><br>Assessment and Plan<br><br>Assessment:<br><br>59-year-old male with idiopathic Parkinson's disease (diagnosed 2 years ago) presenting with tremor-dominant disease showing partial response to levodopa/carbidopa. Current regimen includes levodopa/carbidopa 100/25 mg, | Assessment: The patient reports progressive dose escalation over the past year due to inadequate tremor control, though experiencing improvement in rigidity and bradykinesia. This clinical pattern is consistent with the known differential responsiveness of parkinsonian symptoms to levodopa, with tremor often being less responsive than bradykinesia and rigidity.[1] The good response in rigidity and bradykinesia confirms the diagnosis of idiopathic Parkinson's disease, as lack of response to levodopa would warrant reconsideration of the diagnosis.[1-2] The patient is not yet experiencing motor fluctuations or dyskinesia, which typically emerge after approximately 4 years of levodopa therapy.[1] However, the current strategy of escalating levodopa monotherapy may increase future risk of motor complications.<br><br>Plan: Add opicapone 50 mg once daily at bedtime as adjunctive therapy to the current levodopa/carbidopa regimen. |

| | | |
|---|---|---|
| | 2 tablets three times daily 30 minutes before meals plus 1 tablet at bedtime (total daily levodopa dose: 700 mg), and Laxaday 17 grams once daily for constipation.<br><br>The patient reports progressive dose escalation over the past year due to inadequate tremor control, though experiencing improvement in rigidity and bradykinesia. This clinical pattern is consistent with the known differential responsiveness of parkinsonian symptoms to levodopa, with tremor often being less responsive than bradykinesia and rigidity.[1] The good response in rigidity and bradykinesia confirms the diagnosis of idiopathic Parkinson's disease, as lack of response to levodopa would warrant reconsideration of the diagnosis.[1-2]<br><br>The patient is not yet experiencing motor fluctuations or dyskinesia, which typically emerge after approximately 4 years of levodopa therapy.[1] However, the current strategy of escalating levodopa monotherapy may increase future risk of motor complications.<br><br>Plan:<br><br>Add opicapone 50 mg once daily at bedtime as adjunctive therapy to the current levodopa/carbidopa regimen.<br><br>Rationale: Opicapone is a once-daily COMT inhibitor that is FDA-approved as adjunctive treatment to levodopa/carbidopa in patients with Parkinson's disease and has been shown to be efficacious for motor symptoms in patients on levodopa therapy.[4-5] COMT inhibitors enhance the effect of levodopa by blocking dopamine metabolism, thereby prolonging levodopa's benefits.[1][6] This approach allows for | Rationale: Opicapone is a once-daily COMT inhibitor that is FDA-approved as adjunctive treatment to levodopa/carbidopa in patients with Parkinson's disease and has been shown to be efficacious for motor symptoms in patients on levodopa therapy.[4-5] COMT inhibitors enhance the effect of levodopa by blocking dopamine metabolism, thereby prolonging levodopa's benefits.[1][6] This approach allows for improved symptom control without further escalating the levodopa dose, which may help minimize future risk of motor complications.[2] |

| | |
|---|---|
| | improved symptom control without further escalating the levodopa dose, which may help minimize future risk of motor complications.[2]<br><br>The addition of adjunctive therapy rather than continued levodopa dose escalation is supported by evidence that patients often achieve optimal symptom control with fewer side effects using low or moderate doses of multiple agents rather than higher doses of any single class.[2] Recent evidence demonstrates that COMT inhibitors, including opicapone, are efficacious treatments for motor symptoms in Parkinson's disease, and opicapone specifically offers the convenience of once-daily dosing.[2][5]<br><br>While tremor may remain partially refractory even with optimized dopaminergic therapy, this strategy provides the best evidence-based approach to enhance overall motor function.[1][3] If tremor remains significantly problematic despite this intervention, alternative or additional strategies may be considered in the future.<br><br>Continue levodopa/carbidopa at current dose (100/25 mg, 2 tablets three times daily plus 1 tablet at bedtime) and Laxaday 17 grams once daily.<br><br>Monitor for potential side effects of COMT inhibitor therapy including dyskinesia, nausea, orthostatic hypotension, and diarrhea. Counsel patient that levodopa absorption can be delayed by slow gastric emptying and blocked by concurrent protein intake; doses should be separated from meals by at least 1 hour when possible.[2]<br><br>Follow-up in 4-6 weeks to assess treatment response and tolerability. |

| | | |
|---|---|---|
| 7 | This assessment and plan will focus on fall prevention in a patient with early-stage Parkinson's disease, emphasizing the most evidence-based intervention: referral to physical therapy for a supervised exercise program. The American Geriatrics Society recommends that all people with PD at risk for falls should work with a physical therapist, ideally one specializing in PD, starting at an early stage of disease, and exercise interventions have been shown to reduce fall rates by approximately 26% in patients with mild to moderate PD.[1-2]<br><br>Assessment and Plan<br><br>Assessment:<br><br>63-year-old female with idiopathic Parkinson's disease (diagnosed one year ago) presenting with recurrent falls—two falls in the past six months, both occurring while walking outdoors due to inattention to environmental hazards (sidewalk irregularities). Falls were forward in direction without presyncope, consistent with gait and attention deficits rather than orthostatic hypotension or syncope. The temporal relationship to levodopa/carbidopa dosing is unknown.<br><br>Current medications include levodopa/carbidopa 100/25 mg four times daily (total daily levodopa dose 400 mg) and Laxaday 17 grams once daily for constipation.<br><br>Fall Risk Factors:<br><br>History of falls (strong predictor of future falls)[3]<br><br>Likely gait impairment (shuffling/small-scaled gait increases stumbling risk)[3]<br><br>Possible attention/executive dysfunction | Assessment: Fall Risk Factors: History of falls (strong predictor of future falls)[3]ת Likely gait impairment (shuffling/small-scaled gait increases stumbling risk)[3], Possible attention/executive dysfunction (falls due to inattention to environmental hazards)[3], Early-stage PD with progressive disease severity[3]<br><br>Plan: Primary Intervention - Physical Therapy Referral: Refer to physical therapy, preferably a therapist with expertise in Parkinson's disease, for a supervised multicomponent exercise program. Exercise interventions probably reduce the rate of falls by 26% and probably slightly reduce the number of people experiencing falls in patients with mild to moderate PD.[2] The American Geriatrics Society recommends that all people with PD at risk for falls should work with a physical therapist starting at an early stage of disease, as this strategy reduces disease-related complications and is associated with lower healthcare costs and fewer fractures.[1]<br><br>The physical therapy program should include: Gait training with cueing techniques (visual, auditory, or tactile cues)[3-4] Balance training exercises (such as Tai Chi)[3] Strength training, particularly of lower extremities and trunk[3] Task-specific training for functional activities[5] Dual-task training to address attention deficits during walking[3] Fall prevention strategies and environmental modification recommendations[5] |

| | |
|---|---|
| | (falls due to inattention to environmental hazards)[3]<br><br>Early-stage PD with progressive disease severity[3]<br><br>Plan:<br><br>Primary Intervention - Physical Therapy Referral:<br><br>Refer to physical therapy, preferably a therapist with expertise in Parkinson's disease, for a supervised multicomponent exercise program. Exercise interventions probably reduce the rate of falls by 26% and probably slightly reduce the number of people experiencing falls in patients with mild to moderate PD.[2] The American Geriatrics Society recommends that all people with PD at risk for falls should work with a physical therapist starting at an early stage of disease, as this strategy reduces disease-related complications and is associated with lower healthcare costs and fewer fractures.[1]<br><br>The physical therapy program should include:<br><br>Gait training with cueing techniques (visual, auditory, or tactile cues)[3-4]<br><br>Balance training exercises (such as Tai Chi)[3]<br><br>Strength training, particularly of lower extremities and trunk[3]<br><br>Task-specific training for functional activities[5]<br><br>Dual-task training to address attention deficits during walking[3]<br><br>Fall prevention strategies and |

| | |
|---|---|
| | environmental modification recommendations[5]<br><br>Medication Management:<br><br>Continue current levodopa/carbidopa regimen at 100/25 mg four times daily (total 400 mg levodopa daily). The current dose is within the recommended range for early PD (150-400 mg/day) and below the threshold associated with increased dyskinesia risk.[6] The American Academy of Neurology recommends prescribing the lowest effective dose of levodopa that provides adequate symptomatic benefit.[6]<br><br>Monitor for:<br><br>Motor response to treatment and presence of dyskinesia<br><br>Motor fluctuations or OFF periods that may contribute to falls[3]<br><br>Orthostatic hypotension (particularly during transfers)[3]<br><br>Impulse control disorders, excessive daytime sleepiness, and hallucinations[6]<br><br>If falls are determined to be related to OFF periods with slow mobility or shuffling gait, consider increasing levodopa dose.[3] Conversely, if falls are related to ON-phase dyskinesias or orthostatic hypotension, dose reduction may be warranted.[3]<br><br>Additional Assessments:<br><br>Formal gait speed assessment (self-selected gait speed <1.1 m/s predicts fall risk)[1]<br><br>Screening for freezing of gait using detailed questionnaires and specific |

| | | |
|---|---|---|
| | provocation tasks (360-degree axial turning)[3]<br><br>Cognitive screening, particularly executive function assessment, as part of multifactorial fall risk evaluation[1]<br><br>Timed Up and Go test or similar balance assessment tools[3]<br><br>Orthostatic vital signs, particularly during transfers[3]<br><br>Patient Education:<br><br>Discuss fall prevention strategies, including environmental awareness and modification<br><br>Counsel on importance of adherence to exercise program<br><br>Advise on proper footwear[3]<br><br>Educate regarding potential medication-related fall risks<br><br>Follow-up:<br><br>Reassess in 3 months to evaluate response to physical therapy intervention, monitor for medication-related adverse effects, and adjust treatment plan as needed based on fall frequency and motor symptom control. | |
| 8 | This assessment and plan addresses gait dysfunction with freezing and festination in a patient with established Parkinson's disease on triple therapy. The National Parkinson Foundation guidelines recommend optimizing dopaminergic medication for freezing of gait and festination, with specific guidance to increase levodopa dose or add an MAO-B inhibitor for OFF-phase freezing.[1] Given the patient's current regimen and gait instability, the evidence supports adding | Assessment: The clinical presentation is consistent with freezing of gait (FOG) and festination, which represent one of the most common causes of falls in parkinsonism.[1] The shuffling gait with short, hesitant steps and uncontrolled acceleration (festination) suggests inadequate dopaminergic coverage, particularly given the 4-hour dosing interval which may be leading to OFF periods.[2-3] The worsening with distraction and crowded areas |

| | | |
|---|---|---|
| | a COMT inhibitor to extend levodopa duration and reduce OFF periods, combined with physical therapy interventions that have demonstrated efficacy for freezing of gait.[2-4]<br><br>Assessment and Plan<br><br>Assessment:<br><br>68-year-old female with 6-year history of idiopathic Parkinson's disease presenting with progressive gait dysfunction characterized by shuffling gait, festination, and postural instability without falls to date. Current medications include levodopa/carbidopa 100/25 mg 1.5 tablets four times daily (total daily levodopa dose 600 mg), rotigotine patch 3 mg daily, and amantadine 100 mg twice daily.<br><br>The clinical presentation is consistent with freezing of gait (FOG) and festination, which represent one of the most common causes of falls in parkinsonism.[1] The shuffling gait with short, hesitant steps and uncontrolled acceleration (festination) suggests inadequate dopaminergic coverage, particularly given the 4-hour dosing interval which may be leading to OFF periods.[2-3] The worsening with distraction and crowded areas indicates dual-task interference, a characteristic feature of gait dysfunction in Parkinson's disease.[5]<br><br>Plan:<br><br>Pharmacological Management:<br><br>Add entacapone 200 mg with each levodopa/carbidopa dose (four times daily). Entacapone is a catechol-O-methyltransferase (COMT) inhibitor that prolongs levodopa duration and is | indicates dual-task interference, a characteristic feature of gait dysfunction in Parkinson's disease.[5]<br><br>Plan: Add entacapone 200 mg with each levodopa/carbidopa dose (four times daily). Entacapone is a catechol-O-methyltransferase (COMT) inhibitor that prolongs levodopa duration and is clinically useful for treating motor fluctuations.[6-7] The National Parkinson Foundation guidelines specifically recommend optimizing dopaminergic medication for freezing of gait and festination, including adding an MAO-B inhibitor for OFF-phase freezing.[1] While the patient is already on rasagiline-equivalent coverage via rotigotine, adding entacapone will extend each levodopa dose duration and reduce OFF periods that may be contributing to gait dysfunction.[2][8] |

| | |
|---|---|
| | clinically useful for treating motor fluctuations.[6-7] The National Parkinson Foundation guidelines specifically recommend optimizing dopaminergic medication for freezing of gait and festination, including adding an MAO-B inhibitor for OFF-phase freezing.[1] While the patient is already on rasagiline-equivalent coverage via rotigotine, adding entacapone will extend each levodopa dose duration and reduce OFF periods that may be contributing to gait dysfunction.[2][8]<br><br>Alternative consideration: If inadequate response to entacapone after 4-6 weeks, consider opicapone 50 mg once daily at bedtime as an alternative COMT inhibitor, which is FDA-approved for adjunctive treatment in patients experiencing OFF episodes.[6][9]<br><br>Non-Pharmacological Management:<br><br>Refer to physical therapy for gait training with cueing strategies (visual and auditory cues), which have Level A evidence for reducing freezing of gait.[4] The National Parkinson Foundation guidelines recommend gait training using cues or cognitive movement strategies for freezing of gait and festination.[1] Specific interventions should include:<br><br>Visual and auditory cueing techniques (evidence level A)[4]<br><br>Treadmill training (evidence level A)[4][10]<br><br>Balance training and dual-task training to address instability in complex environments[1]<br><br>Home safety evaluation and environmental modifications, including optimizing lighting and creating larger |

| | | |
|---|---|---|
| | spaces for turning[1]<br><br>Monitoring and Follow-up:<br><br>Reassess in 4-6 weeks to evaluate response to entacapone and physical therapy<br><br>Monitor for dyskinesia development with increased dopaminergic medication[2]<br><br>Assess for orthostatic hypotension at follow-up visits[1]<br><br>Consider formal gait assessment using Timed Up and Go test or Tinetti mobility index[1]<br><br>Educate patient and family about fall prevention strategies given high fall risk with festination[1]<br><br>Safety Considerations:<br><br>The patient has not yet fallen but demonstrates significant fall risk factors including festination, postural instability, and dual-task interference.[1] Falls in Parkinson's disease most commonly occur while patients are on medication, though this may reflect disease severity rather than a direct medication effect.[2] Close monitoring during medication optimization is essential. | |
| 9 | The following assessment and plan addresses cognitive impairment in a patient with advanced Parkinson's disease. The treatment recommendation is grounded in evidence from multiple randomized controlled trials demonstrating efficacy of rivastigmine for Parkinson's disease dementia, which is the only FDA-approved medication for this indication and is designated as clinically useful by the International Parkinson and Movement Disorder Society.[1-3] | Assessment: The clinical presentation is consistent with PDD rather than mild cognitive impairment given the multi-domain cognitive deficits and functional impairment affecting activities of daily living.[4-5] The cognitive profile—featuring memory problems, executive dysfunction, and attentional difficulties—is typical for PDD.[4][6]<br><br>Plan: Initiate rivastigmine for treatment of Parkinson's disease dementia. |

| | | |
|---|---|---|
| | Assessment and Plan<br><br>Assessment:<br><br>78-year-old male with 12-year history of idiopathic Parkinson's disease presenting with progressive cognitive decline consistent with Parkinson's disease dementia (PDD). Current symptoms include:<br><br>Memory impairment: forgetting recent events, misplacing items, requiring repeated reminders for appointments<br><br>Executive dysfunction: difficulty planning simple tasks, trouble concentrating<br><br>Language/communication difficulties: difficulty following conversations<br><br>Functional decline: marked deterioration from baseline cognitive abilities<br><br>Current Parkinson's disease medications:<br><br>Levodopa/carbidopa 100/25 mg, 2 tablets four times daily (total daily levodopa dose: 800 mg)<br><br>Entacapone 200 mg four times daily<br><br>Rasagiline 1 mg once daily<br><br>The clinical presentation is consistent with PDD rather than mild cognitive impairment given the multi-domain cognitive deficits and functional impairment affecting activities of daily living.[4-5] The cognitive profile—featuring memory problems, executive dysfunction, and attentional difficulties—is typical for PDD.[4][6]<br><br>Plan: | Rivastigmine is the only medication with consistent evidence of efficacy for PDD and is FDA-approved for this indication.[1][7] In the pivotal EXPRESS trial of 362 patients with PDD, rivastigmine (3-12 mg daily) demonstrated significant improvement in global cognition (mean 2.1-point improvement on the Alzheimer Disease Assessment Scale vs. 0.7-point decline with placebo, $p<0.001$), as well as improvements in attention, executive functions, neuropsychiatric symptoms, and activities of daily living.[1-2] The International Parkinson and Movement Disorder Society designates rivastigmine as clinically useful based on this high-level evidence.[1-2]<br><br>Dosing regimen: Start rivastigmine 1.5 mg twice daily (oral capsule or solution) Titrate by 1.5 mg twice daily every 4 weeks as tolerated. Target dose: 3-6 mg twice daily (maximum 6 mg twice daily) Alternative: rivastigmine transdermal patch may be used if oral formulation is not tolerated, starting at 4.6 mg/24 hours and titrating to 9.5 mg/24 hours or 13.3 mg/24 hours<br><br>Monitoring:<br>Assess for gastrointestinal side effects (nausea, vomiting, diarrhea), which are common but often transient[2-3]<br>Monitor for potential transient worsening of tremor during initial treatment, though sustained motor worsening is not expected[2][8]<br>Reassess cognitive function, neuropsychiatric symptoms, and functional status at follow-up visits |

| | |
|---|---|
| | Initiate rivastigmine for treatment of Parkinson's disease dementia. Rivastigmine is the only medication with consistent evidence of efficacy for PDD and is FDA-approved for this indication.[1][7] In the pivotal EXPRESS trial of 362 patients with PDD, rivastigmine (3-12 mg daily) demonstrated significant improvement in global cognition (mean 2.1-point improvement on the Alzheimer Disease Assessment Scale vs. 0.7-point decline with placebo, p<0.001), as well as improvements in attention, executive functions, neuropsychiatric symptoms, and activities of daily living.[1-2] The International Parkinson and Movement Disorder Society designates rivastigmine as clinically useful based on this high-level evidence.[1-2]<br><br>Dosing regimen:<br><br>Start rivastigmine 1.5 mg twice daily (oral capsule or solution)<br><br>Titrate by 1.5 mg twice daily every 4 weeks as tolerated<br><br>Target dose: 3-6 mg twice daily (maximum 6 mg twice daily)<br><br>Alternative: rivastigmine transdermal patch may be used if oral formulation is not tolerated, starting at 4.6 mg/24 hours and titrating to 9.5 mg/24 hours or 13.3 mg/24 hours<br><br>Monitoring:<br><br>Assess for gastrointestinal side effects (nausea, vomiting, diarrhea), which are common but often transient[2-3]<br><br>Monitor for potential transient worsening of tremor during initial treatment, though sustained motor worsening is not |

| | |
|---|---|
| | expected[2][8]<br><br>Reassess cognitive function, neuropsychiatric symptoms, and functional status at follow-up visits<br><br>Consider dose adjustment based on response and tolerability<br><br>Additional considerations:<br><br>Continue current Parkinson's disease medications as prescribed; dopaminergic therapy optimization may help some cognitive symptoms but is not a primary treatment for dementia[4][6]<br><br>Avoid anticholinergic medications, which are a substantial risk factor for worsening dementia in Parkinson's disease[8]<br><br>Address modifiable vascular risk factors (blood pressure, diabetes, hyperlipidemia) as dementia in PD may involve mixed pathologies including vascular disease[9]<br><br>Consider referral to neuropsychology for formal cognitive assessment if not already completed<br><br>Provide caregiver education and support resources given the significant impact of PDD on both patients and families[7]<br><br>Prognosis and expectations:<br><br>Rivastigmine provides modest symptomatic benefit and may stabilize cognitive function, but does not modify disease progression.[4-6] Response varies among individuals, with some patients showing marked or moderate improvement (approximately 20% combined) while others remain stable or decline.[2] Benefits may be sustained for |

| | | |
|---|---|---|
| | up to 48 weeks based on open-label extension data.[2] | |
| 10 | The assessment and plan will address visual hallucinations in this patient with advanced Parkinson's disease, focusing on medication optimization. The evidence supports acetylcholinesterase inhibitors as first-line pharmacological treatment for visual hallucinations in Parkinson's disease, with rivastigmine having the strongest evidence base in this population.[1-4]<br><br>Assessment and Plan<br><br>Assessment:<br><br>80-year-old female with 10-year history of idiopathic Parkinson's disease presenting with distressing visual hallucinations consisting of well-formed complex images (children, small animals, deceased relatives). The patient demonstrates preserved insight as she can often recognize these visions as unreal upon checking, though they cause significant anxiety. She is currently on optimized dopaminergic therapy including levodopa/carbidopa 100/25 mg 2 tablets four times daily, safinamide 100 mg daily, and amantadine 100 mg twice daily.<br><br>Visual hallucinations are common in Parkinson's disease and represent dysfunction in cholinergic, serotonergic, and dopaminergic systems.[1] The presence of well-formed visual hallucinations with retained insight is characteristic of Parkinson's disease psychosis.[1][5] Medical triggers such as infection, constipation, or metabolic disturbances have been considered and excluded.[1]<br><br>Plan: | Assessment: Visual hallucinations are common in Parkinson's disease and represent dysfunction in cholinergic, serotonergic, and dopaminergic systems.[1] The presence of well-formed visual hallucinations with retained insight is characteristic of Parkinson's disease psychosis.[1][5] Medical triggers such as infection, constipation, or metabolic disturbances have been considered and excluded.[1]<br><br>Plan: Initiate rivastigmine 1.5 mg twice daily with food, titrating up by 1.5 mg twice daily every 4 weeks as tolerated to a target dose of 6 mg twice daily (or maximum tolerated dose up to 6 mg twice daily).<br>Rationale: Acetylcholinesterase inhibitors, particularly rivastigmine and donepezil, are effective for visual hallucinations in Parkinson's disease even in the absence of dementia.[1-2] Rivastigmine has demonstrated efficacy in reducing visual hallucinations in patients with Parkinson's disease dementia and may have a preventive effect on incident visual hallucinations.[2] Cholinesterase inhibitors are recommended as first-line pharmacological treatment for well-formed visual hallucinations in Parkinson's disease.[3] The cholinergic deficit contributes to the pathophysiology of visual hallucinations in synucleinopathies.[6] Continue current dopaminergic regimen unchanged at this time, as the patient's motor symptoms appear adequately controlled and reduction of dopaminergic medications is not feasible without compromising motor function.[2] Monitor for common side effects of rivastigmine including nausea, vomiting, diarrhea, decreased |

| | | |
|---|---|---|
| | Initiate rivastigmine 1.5 mg twice daily with food, titrating up by 1.5 mg twice daily every 4 weeks as tolerated to a target dose of 6 mg twice daily (or maximum tolerated dose up to 6 mg twice daily).<br><br>Rationale: Acetylcholinesterase inhibitors, particularly rivastigmine and donepezil, are effective for visual hallucinations in Parkinson's disease even in the absence of dementia.[1-2] Rivastigmine has demonstrated efficacy in reducing visual hallucinations in patients with Parkinson's disease dementia and may have a preventive effect on incident visual hallucinations.[2] Cholinesterase inhibitors are recommended as first-line pharmacological treatment for well-formed visual hallucinations in Parkinson's disease.[3] The cholinergic deficit contributes to the pathophysiology of visual hallucinations in synucleinopathies.[6]<br><br>Continue current dopaminergic regimen unchanged at this time, as the patient's motor symptoms appear adequately controlled and reduction of dopaminergic medications is not feasible without compromising motor function.[2]<br><br>Monitor for common side effects of rivastigmine including nausea, vomiting, diarrhea, decreased appetite, and dizziness. Advise taking with food to minimize gastrointestinal side effects.<br><br>Follow-up in 4-6 weeks to assess response to treatment and tolerability. If visual hallucinations persist despite adequate trial of rivastigmine at therapeutic doses, consider adding low-dose quetiapine (12.5-50 mg daily) or referral for consideration of pimavanserin, which is FDA-approved for Parkinson's disease psychosis.[1-2][7] | appetite, and dizziness. Advise taking with food to minimize gastrointestinal side effects.<br>Follow-up in 4-6 weeks to assess response to treatment and tolerability. If visual hallucinations persist despite adequate trial of rivastigmine at therapeutic doses, consider adding low-dose quetiapine (12.5-50 mg daily) or referral for consideration of pimavanserin, which is FDA-approved for Parkinson's disease psychosis.[1-2][7] Provide education and reassurance to patient and caregiver about the nature of visual hallucinations in Parkinson's disease and the expected timeline for treatment response.[5][8] |

| | | |
|---|---|---|
| | Provide education and reassurance to patient and caregiver about the nature of visual hallucinations in Parkinson's disease and the expected timeline for treatment response.[5][8] | |
| 11 | The following assessment and plan addresses treatment-resistant depression in Parkinson's disease. The evidence supports optimizing antidepressant therapy, with tricyclic antidepressants (particularly nortriptyline) and SNRIs showing superior efficacy compared to SSRIs in this population.[1-2] The recommendation is grounded in randomized controlled trial data demonstrating significant improvement in depressive symptoms without worsening motor function.<br><br>Assessment and Plan<br><br>Assessment:<br><br>65-year-old female with idiopathic Parkinson's disease (6-year duration) presenting with treatment-resistant major depressive disorder despite 12 months of sertraline 50 mg daily. Current symptoms meet criteria for major depressive disorder, including persistent sadness, anhedonia, social withdrawal, low energy, poor appetite, and passive suicidal ideation without active suicidal thoughts. Motor symptoms are currently managed with levodopa/carbidopa 100/25 mg three times daily and selegiline 5 mg twice daily.<br><br>The patient's depression has not adequately responded to SSRI monotherapy at the current dose. Evidence suggests that SSRIs may have limited efficacy in Parkinson's disease-related depression, with tricyclic antidepressants and SNRIs demonstrating superior outcomes in this population.[1- | Assessment: Current symptoms meet criteria for major depressive disorder, including persistent sadness, anhedonia, social withdrawal, low energy, poor appetite, and passive suicidal ideation without active suicidal thoughts. Motor symptoms are currently managed with levodopa/carbidopa 100/25 mg three times daily and selegiline 5 mg twice daily.<br>The patient's depression has not adequately responded to SSRI monotherapy at the current dose. Evidence suggests that SSRIs may have limited efficacy in Parkinson's disease-related depression, with tricyclic antidepressants and SNRIs demonstrating superior outcomes in this population.[1-5]<br><br>Plan: Switch from sertraline to nortriptyline for treatment of depression in Parkinson's disease. This recommendation is based on Class I evidence demonstrating that nortriptyline significantly improves depressive symptoms in Parkinson's disease patients, with a mean reduction in Hamilton Depression Rating Scale scores of 6.2 points greater than placebo and a response rate of 53% compared to 11% for paroxetine (an SSRI).[1-2] Nortriptyline has been shown to improve not only depression but also anxiety, sleep, and social functioning without worsening motor symptoms.[2]<br><br>Specific dosing regimen:<br>Taper sertraline 50 mg daily to 25 mg |

| | | |
|---|---|---|
| | 5]<br><br>Plan:<br><br>Switch from sertraline to nortriptyline for treatment of depression in Parkinson's disease. This recommendation is based on Class I evidence demonstrating that nortriptyline significantly improves depressive symptoms in Parkinson's disease patients, with a mean reduction in Hamilton Depression Rating Scale scores of 6.2 points greater than placebo and a response rate of 53% compared to 11% for paroxetine (an SSRI).[1-2] Nortriptyline has been shown to improve not only depression but also anxiety, sleep, and social functioning without worsening motor symptoms.[2]<br><br>Specific dosing regimen:<br><br>Taper sertraline 50 mg daily to 25 mg daily for 1 week, then discontinue<br><br>Begin nortriptyline 25 mg at bedtime after sertraline discontinuation<br><br>Titrate nortriptyline by 25 mg every 3-7 days as tolerated<br><br>Target dose: nortriptyline 50-75 mg at bedtime (maximum studied dose 100 mg daily)[2]<br><br>Monitor for anticholinergic side effects (dry mouth, constipation, urinary retention, cognitive effects)<br><br>Obtain baseline ECG given tricyclic use<br><br>Rationale: Multiple randomized controlled trials and network meta-analyses support the use of dual reuptake inhibitors (tricyclics and SNRIs) over SSRIs for Parkinson's disease-related depression.[3-7] Nortriptyline specifically | daily for 1 week, then discontinue<br>Begin nortriptyline 25 mg at bedtime after sertraline discontinuation<br>Titrate nortriptyline by 25 mg every 3-7 days as tolerated<br>Target dose: nortriptyline 50-75 mg at bedtime (maximum studied dose 100 mg daily)[2]<br>Monitor for anticholinergic side effects (dry mouth, constipation, urinary retention, cognitive effects)<br>Obtain baseline ECG given tricyclic use<br>Rationale: Multiple randomized controlled trials and network meta-analyses support the use of dual reuptake inhibitors (tricyclics and SNRIs) over SSRIs for Parkinson's disease-related depression.[3-7] Nortriptyline specifically demonstrated superiority over both paroxetine and placebo in the largest head-to-head comparison.[2] While SNRIs such as venlafaxine are also effective, nortriptyline may offer additional benefits for sleep and anxiety that are common comorbidities in this patient.[1-2] |

| | |
|---|---|
| | demonstrated superiority over both paroxetine and placebo in the largest head-to-head comparison.[2] While SNRIs such as venlafaxine are also effective, nortriptyline may offer additional benefits for sleep and anxiety that are common comorbidities in this patient.[1-2]<br><br>Additional considerations:<br><br>Continue current Parkinson's disease medications (levodopa/carbidopa and selegiline) as dopaminergic optimization is an important first step in managing depression[5][8]<br><br>Monitor for potential serotonin syndrome during transition, though risk is low with appropriate tapering<br><br>Reassess depressive symptoms at 4 weeks and 8 weeks using validated depression scales<br><br>Consider cognitive behavioral therapy as adjunctive treatment, which has demonstrated benefit in Parkinson's disease depression[4][8]<br><br>If inadequate response to nortriptyline at 8 weeks, consider switching to venlafaxine XR (target dose 150-225 mg daily) as an alternative evidence-based option[1]<br><br>Safety monitoring:<br><br>Follow up in 1-2 weeks to assess tolerability during titration<br><br>Monitor for orthostatic hypotension, which may be exacerbated by tricyclic use in the context of Parkinson's disease-related autonomic dysfunction<br><br>Assess suicidal ideation at each visit given |

| | | |
|---|---|---|
| | passive thoughts about life's value<br><br>Screen for emergent impulse control disorders, though these are more commonly associated with dopamine agonists than current regimen | |
| 12 | This patient presents with anxiety symptoms that appear to be both generalized and related to medication wearing-off periods. The medical literature supports optimizing dopaminergic therapy as the first-line approach for anxiety associated with motor fluctuations, followed by consideration of SSRIs if anxiety persists.[1-3] The assessment and plan below will address both the motor fluctuation-related anxiety and the generalized anxiety symptoms with evidence-based recommendations.<br><br>Assessment and Plan<br><br>Assessment:<br><br>60-year-old male with idiopathic Parkinson's disease (4-year duration) presenting with anxiety symptoms that appear to be predominantly related to medication wearing-off periods. The patient describes episodes of intense anxiety, shakiness, sweating, and panic occurring specifically as his levodopa/carbidopa dose wears off, along with baseline excessive worry. He is currently on levodopa/carbidopa 100/25 mg four times daily every 4 hours, rasagiline 1 mg daily, and clonazepam 0.5 mg at bedtime as needed.<br><br>The temporal relationship between anxiety symptoms and medication wearing-off strongly suggests non-motor fluctuations (off-period anxiety), which is a common manifestation of Parkinson's disease.[1-3] The patient also exhibits features of generalized anxiety with | A: The temporal relationship between anxiety symptoms and medication wearing-off strongly suggests non-motor fluctuations (off-period anxiety), which is a common manifestation of Parkinson's disease.[1-3] The patient also exhibits features of generalized anxiety with excessive worry. Current dopaminergic therapy appears suboptimal given the 4-hour dosing interval with breakthrough symptoms.<br><br>P: Optimize dopaminergic therapy - This is the first-line approach for anxiety associated with motor fluctuations.[1-3]<br>Increase levodopa/carbidopa to 100/25 mg 1.5 tablets four times daily (total daily levodopa dose increasing from 400 mg to 600 mg)<br>Rationale: Anxiety occurring as an off-period symptom should prompt review and optimization of levodopa timing and dosing. Increasing dopaminergic therapy can address both depression and anxiety related to off-phases and non-motor fluctuations.[1][4-5] |

| | |
|---|---|
| | excessive worry. Current dopaminergic therapy appears suboptimal given the 4-hour dosing interval with breakthrough symptoms.<br><br>Plan:<br><br>1. Optimize dopaminergic therapy - This is the first-line approach for anxiety associated with motor fluctuations.[1-3]<br><br>Increase levodopa/carbidopa to 100/25 mg 1.5 tablets four times daily (total daily levodopa dose increasing from 400 mg to 600 mg)<br><br>Rationale: Anxiety occurring as an off-period symptom should prompt review and optimization of levodopa timing and dosing. Increasing dopaminergic therapy can address both depression and anxiety related to off-phases and non-motor fluctuations.[1][4-5]<br><br>Monitor for dyskinesias or other dopaminergic side effects at follow-up<br><br>2. Initiate SSRI therapy for persistent anxiety<br><br>Start escitalopram 10 mg once daily in the morning<br><br>Rationale: SSRIs are the front-line psychopharmacology for anxiety in Parkinson's disease and have demonstrated safety and efficacy in this population. Escitalopram is well-tolerated and is being studied specifically in Parkinson's disease depression/anxiety.[1-2][6]<br><br>The combination of rasagiline (MAO-B inhibitor) with SSRIs carries a theoretical risk of serotonin syndrome, but this risk is rare and should not lead to under-prescribing[1] |

| | | |
|---|---|---|
| | May titrate to 20 mg daily after 4-6 weeks if inadequate response<br><br>3. Continue current medications<br><br>Rasagiline 1 mg daily - continue<br><br>Clonazepam 0.5 mg at bedtime as needed - continue for now, but consider tapering once SSRI reaches therapeutic effect given fall risk; low-dose benzodiazepines may be needed for non-motor fluctuations in some patients[2]<br><br>4. Non-pharmacological interventions<br><br>Refer for cognitive behavioral therapy tailored for Parkinson's disease, which has demonstrated statistically and clinically significant improvements in anxiety[2]<br><br>Consider telemedicine-based CBT given potential mobility limitations[2]<br><br>5. Follow-up<br><br>Return visit in 4 weeks to assess response to medication adjustments<br><br>Monitor for improvement in off-period anxiety symptoms and generalized anxiety<br><br>Assess for adverse effects including dyskinesias, serotonin syndrome symptoms, and fall risk<br><br>Consider further dopaminergic optimization or SSRI dose adjustment based on response | |
| 13 | This assessment and plan addresses apathy in Parkinson's disease, a distinct neuropsychiatric syndrome characterized by lack of motivation without depression. The evidence supports pharmacological | A: Clinical features are consistent with parkinsonian apathy: patient demonstrates reduced self-initiated activity, requires external prompting for basic tasks (dressing, walking), and |

| | | |
|---|---|---|
| | intervention with dopaminergic agents, particularly dopamine D2/D3 agonists like piribedil, which have demonstrated efficacy in randomized controlled trials for apathy in Parkinson's disease. Rivastigmine also shows benefit but is typically reserved for patients with cognitive impairment.[1-2]<br><br>Assessment and Plan<br><br>Assessment:<br><br>70-year-old male with 8-year history of idiopathic Parkinson's disease presenting with apathy—characterized by lack of motivation, reduced goal-directed behavior, and diminished initiation of activities without accompanying sadness or depressed mood. Current dopaminergic regimen includes levodopa/carbidopa 100/25 mg 2 tablets four times daily (total daily levodopa dose 800 mg), entacapone 200 mg four times daily, and safinamide 100 mg once daily.<br><br>Clinical features are consistent with parkinsonian apathy: patient demonstrates reduced self-initiated activity, requires external prompting for basic tasks (dressing, walking), and has lost interest in previously enjoyed hobbies while explicitly denying depressive symptoms. This presentation distinguishes apathy from depression, as the patient reports indifference rather than sadness.[1]<br><br>Apathy is a prevalent non-motor symptom in Parkinson's disease that significantly impacts quality of life and caregiver burden. The underlying pathophysiology involves mesolimbic dopaminergic denervation affecting motivation circuits.[1-3]<br><br>Plan: | has lost interest in previously enjoyed hobbies while explicitly denying depressive symptoms. This presentation distinguishes apathy from depression, as the patient reports indifference rather than sadness.[1]<br>Apathy is a prevalent non-motor symptom in Parkinson's disease that significantly impacts quality of life and caregiver burden. The underlying pathophysiology involves mesolimbic dopaminergic denervation affecting motivation circuits.[1-3]<br><br>P: Pharmacological intervention with piribedil, a dopamine D2/D3 receptor agonist, is recommended based on Class I evidence demonstrating efficacy for apathy in Parkinson's disease.[1-2]<br><br>Specific Treatment Recommendation:<br>Piribedil 50 mg three times daily initially, titrated up to 100 mg three times daily (total 300 mg/day) over 4-6 weeks<br>Rationale: A randomized, double-blind, placebo-controlled trial in 37 patients with Parkinson's disease and apathy (Starkstein Apathy Scale score >14) demonstrated that piribedil reduced apathy scores by 34.6% compared to 3.2% with placebo (p=0.015) over 12 weeks. This represents Class I evidence for efficacy. The study also showed trends toward improvement in quality of life and anhedonia.[1-2]<br><br>Titration schedule:<br>Week 1-2: 50 mg three times daily<br>Week 3-4: 50 mg twice daily + 100 mg once daily<br>Week 5-6: 100 mg three times daily (target dose) |

| | |
|---|---|
| | Pharmacological intervention with piribedil, a dopamine D2/D3 receptor agonist, is recommended based on Class I evidence demonstrating efficacy for apathy in Parkinson's disease.[1-2]<br><br>Specific Treatment Recommendation:<br><br>Piribedil 50 mg three times daily initially, titrated up to 100 mg three times daily (total 300 mg/day) over 4-6 weeks<br><br>Rationale: A randomized, double-blind, placebo-controlled trial in 37 patients with Parkinson's disease and apathy (Starkstein Apathy Scale score >14) demonstrated that piribedil reduced apathy scores by 34.6% compared to 3.2% with placebo (p=0.015) over 12 weeks. This represents Class I evidence for efficacy. The study also showed trends toward improvement in quality of life and anhedonia.[1-2]<br><br>Titration schedule:<br><br>Week 1-2: 50 mg three times daily<br><br>Week 3-4: 50 mg twice daily + 100 mg once daily<br><br>Week 5-6: 100 mg three times daily (target dose)<br><br>Alternative considerations if piribedil is not available or not tolerated:<br><br>Rivastigmine 4.6 mg/24-hour patch, titrated to 9.5 mg/24-hour patch, showed significant improvement in apathy (LARS score) in a double-blind placebo-controlled study of 31 patients with Parkinson's disease without dementia (p=0.031). However, this cholinesterase inhibitor is typically reserved for patients with cognitive impairment.[1] |

| | | |
|---|---|---|
| | Methylphenidate has shown preliminary benefit in small studies but lacks robust Class I evidence specifically for apathy in Parkinson's disease.[1]<br><br>Monitoring:<br><br>Assess apathy severity at baseline and 12 weeks using validated scale (Starkstein Apathy Scale or Lille Apathy Rating Scale)<br><br>Monitor for dopaminergic side effects (dyskinesia, impulse control disorders, hallucinations)<br><br>Evaluate functional improvement and caregiver burden<br><br>Non-pharmacological adjuncts:<br><br>Encourage structured daily activities and exercise-based interventions, which show moderate-quality evidence for benefit.[4-5]<br><br>Consider referral for behavioral interventions adapted from dementia care strategies.[6]<br><br>The current levodopa regimen should be continued as optimizing dopaminergic therapy plays an important role in managing apathy. However, there is no evidence to support antidepressants for apathy in Parkinson's disease, and selective serotonin reuptake inhibitors may paradoxically worsen apathy.[1][6-7] | |
| 14 | This assessment and plan addresses impulse control and related behavioral disorders (ICBDs) in Parkinson's disease, specifically disinhibited speech with sexual content. The evidence strongly supports that dopamine agonist reduction is the primary treatment approach, though this patient is not on a dopamine agonist.[1-3] The literature | This assessment and plan addresses impulse control and related behavioral disorders (ICBDs) in Parkinson's disease, specifically disinhibited speech with sexual content. The evidence strongly supports that dopamine agonist reduction is the primary treatment approach, though this patient is not on a dopamine |

| | | |
|---|---|---|
| | indicates that excessive levodopa use itself can contribute to ICBDs, and the patient's self-escalating levodopa dosing pattern is concerning for dopamine dysregulation syndrome, which requires specific management strategies.[1][4]<br><br>Assessment and Plan<br><br>Assessment:<br><br>55-year-old male with 7-year history of idiopathic Parkinson's disease presenting with impulse control and related behavioral disorder (ICBD) manifesting as disinhibited speech with sexual content and inappropriate social behavior. The patient demonstrates concerning features of dopamine dysregulation syndrome evidenced by self-escalating levodopa/carbidopa use beyond prescribed dosing (taking extra half to full tablets when feeling energy dip).<br><br>Current medications:<br><br>Levodopa/carbidopa 100/25 mg, 2 tablets four times daily (prescribed dose: 800 mg levodopa/day)<br><br>Safinamide 100 mg once daily<br><br>Citalopram 20 mg once daily<br><br>Laxaday 17 g once daily<br><br>The disinhibited behavior with sexual content represents a significant ICBD that is impacting social functioning and creating problematic situations. While dopamine agonists are the most common culprit for ICBDs in Parkinson's disease, this patient is not on a dopamine agonist.[1-3] However, the pattern of compulsive levodopa use and behavioral disinhibition suggests that excessive pulsatile dopaminergic stimulation from | agonist.[1-3] The literature indicates that excessive levodopa use itself can contribute to ICBDs, and the patient's self-escalating levodopa dosing pattern is concerning for dopamine dysregulation syndrome, which requires specific management strategies.[1][4]<br><br>Assessment and Plan<br><br>Assessment:<br><br>55-year-old male with 7-year history of idiopathic Parkinson's disease presenting with impulse control and related behavioral disorder (ICBD) manifesting as disinhibited speech with sexual content and inappropriate social behavior. The patient demonstrates concerning features of dopamine dysregulation syndrome evidenced by self-escalating levodopa/carbidopa use beyond prescribed dosing (taking extra half to full tablets when feeling energy dip).<br><br>Current medications:<br><br>Levodopa/carbidopa 100/25 mg, 2 tablets four times daily (prescribed dose: 800 mg levodopa/day)<br><br>Safinamide 100 mg once daily<br><br>Citalopram 20 mg once daily<br><br>Laxaday 17 g once daily<br><br>The disinhibited behavior with sexual content represents a significant ICBD that is impacting social functioning and creating problematic situations. While dopamine agonists are the most common culprit for ICBDs in Parkinson's disease, this patient is not on a dopamine agonist.[1-3] However, |

| | | |
|---|---|---|
| | self-escalating levodopa dosing is likely contributing to the ICBD presentation.[4]<br><br>Plan:<br><br>Primary intervention: Structured reduction of total daily levodopa dose with transition to more continuous dopaminergic stimulation<br><br>Given the absence of dopamine agonist therapy and evidence that excessive levodopa use can contribute to ICBDs, the treatment approach focuses on reducing pulsatile dopaminergic stimulation while maintaining motor control:[4]<br><br>Reduce levodopa/carbidopa to 100/25 mg, 1.5 tablets four times daily (total 600 mg levodopa/day), representing a 25% reduction from prescribed dose and likely greater reduction from actual intake<br><br>Implement strict dosing schedule with patient and family education about the risks of self-escalating doses and the connection between excessive dopaminergic medication and behavioral symptoms[1]<br><br>Close monitoring with follow-up in 2-3 weeks to assess both motor symptoms and behavioral changes, as reduction in dopaminergic therapy may improve ICBDs but risks motor symptom worsening[2-3]<br><br>Caregiver involvement is essential for monitoring medication adherence and behavioral symptoms[1]<br><br>Consider cognitive behavioral therapy referral, as this has shown efficacy for ICBDs in Parkinson's disease[2]<br><br>If behavioral symptoms persist despite levodopa reduction or if motor symptoms significantly worsen, second-line options | the pattern of compulsive levodopa use and behavioral disinhibition suggests that excessive pulsatile dopaminergic stimulation from self-escalating levodopa dosing is likely contributing to the ICBD presentation.[4]<br><br>Plan:<br><br>Primary intervention: Structured reduction of total daily levodopa dose with transition to more continuous dopaminergic stimulation<br><br>Given the absence of dopamine agonist therapy and evidence that excessive levodopa use can contribute to ICBDs, the treatment approach focuses on reducing pulsatile dopaminergic stimulation while maintaining motor control:[4]<br><br>Reduce levodopa/carbidopa to 100/25 mg, 1.5 tablets four times daily (total 600 mg levodopa/day), representing a 25% reduction from prescribed dose and likely greater reduction from actual intake<br><br>Implement strict dosing schedule with patient and family education about the risks of self-escalating doses and the connection between excessive dopaminergic medication and behavioral symptoms[1]<br><br>Close monitoring with follow-up in 2-3 weeks to assess both motor symptoms and behavioral changes, as reduction in dopaminergic therapy may improve ICBDs but risks motor symptom worsening[2-3]<br><br>Caregiver involvement is essential for monitoring medication adherence and behavioral symptoms[1] |

| | | |
|---|---|---|
| | include quetiapine 25-50 mg at bedtime or consideration of subthalamic nucleus deep brain stimulation evaluation for advanced disease with refractory ICBDs.[1][4] The current SSRI (citalopram) may be continued as antidepressants have been suggested as potential adjunctive therapy for ICBDs, though evidence is limited.[1]<br><br>Monitoring for dopamine agonist withdrawal syndrome is not applicable in this case given absence of dopamine agonist therapy, but monitoring for worsening motor symptoms, mood changes, and autonomic symptoms during levodopa reduction is essential.[2] | Consider cognitive behavioral therapy referral, as this has shown efficacy for ICBDs in Parkinson's disease[2]<br><br>If behavioral symptoms persist despite levodopa reduction or if motor symptoms significantly worsen, second-line options include quetiapine 25-50 mg at bedtime or consideration of subthalamic nucleus deep brain stimulation evaluation for advanced disease with refractory ICBDs.[1][4] The current SSRI (citalopram) may be continued as antidepressants have been suggested as potential adjunctive therapy for ICBDs, though evidence is limited.[1]<br><br>Monitoring for dopamine agonist withdrawal syndrome is not applicable in this case given absence of dopamine agonist therapy, but monitoring for worsening motor symptoms, mood changes, and autonomic symptoms during levodopa reduction is essential.[2] |
| 15 | The following assessment and plan prioritizes cognitive behavioral therapy for insomnia (CBT-I) as first-line treatment, consistent with current guidelines from the American College of Physicians and the U.S. Department of Veterans Affairs/Department of Defense.[1-2] Given that CBT-I may not be immediately accessible and the patient has sleep maintenance insomnia specifically, a dual orexin receptor antagonist is recommended as the pharmacologic option with the strongest evidence for this presentation.[3-4]<br><br>Assessment and Plan<br><br>Assessment:<br><br>63-year-old female with idiopathic Parkinson's disease (8-year duration) | A: The patient meets criteria for chronic insomnia disorder with predominant sleep maintenance difficulty. While Parkinson's disease commonly causes sleep fragmentation, her preserved mobility upon awakening and inconsistent relationship to nocturia suggest the insomnia may have a significant primary component amenable to targeted treatment.<br><br>Plan: Cognitive behavioral therapy for insomnia (CBT-I) is recommended as first-line treatment for chronic insomnia disorder.[1-2] CBT-I includes sleep restriction therapy, stimulus control, relaxation techniques, and cognitive therapy targeting maladaptive beliefs about sleep.[5-6] This multicomponent approach |

| | | |
|---|---|---|
| | presenting with chronic insomnia disorder, specifically sleep maintenance insomnia. She experiences frequent nocturnal awakenings every few hours with difficulty returning to sleep, resulting in unrefreshing sleep. Sleep onset is preserved. Awakenings are not consistently related to nocturia or motor symptoms (she reports no difficulty moving to the bathroom). Current medications include levodopa/carbidopa 100/25 mg 1.5 tablets four times daily, rasagiline 1 mg daily, citalopram 10 mg daily, and Laxaday 17 g as needed.<br><br>The patient meets criteria for chronic insomnia disorder with predominant sleep maintenance difficulty. While Parkinson's disease commonly causes sleep fragmentation, her preserved mobility upon awakening and inconsistent relationship to nocturia suggest the insomnia may have a significant primary component amenable to targeted treatment.<br><br>Plan:<br><br>Cognitive behavioral therapy for insomnia (CBT-I) is recommended as first-line treatment for chronic insomnia disorder.[1-2] CBT-I includes sleep restriction therapy, stimulus control, relaxation techniques, and cognitive therapy targeting maladaptive beliefs about sleep.[5-6] This multicomponent approach produces substantial and sustained improvements in sleep maintenance, with evidence showing superiority over pharmacotherapy in long-term outcomes.[2] CBT-I can be delivered via individual therapy, group sessions, or telehealth platforms.[2][5]<br><br>Refer to behavioral sleep medicine specialist or trained clinician for CBT-I | produces substantial and sustained improvements in sleep maintenance, with evidence showing superiority over pharmacotherapy in long-term outcomes.[2] CBT-I can be delivered via individual therapy, group sessions, or telehealth platforms.[2][5]<br>Refer to behavioral sleep medicine specialist or trained clinician for CBT-I |

| | |
|---|---|
| | If CBT-I is not immediately available, consider brief behavioral treatment for insomnia (BBT-I) focusing on sleep restriction and stimulus control[2][6]<br><br>Given the patient's sleep maintenance insomnia and potential need for pharmacologic intervention while awaiting or supplementing behavioral therapy, the committed medication recommendation is:<br><br>Suvorexant 10 mg orally at bedtime<br><br>Suvorexant is a dual orexin receptor antagonist specifically effective for sleep maintenance insomnia.[3-4] Professional society guidelines identify suvorexant as a preferred option for patients with difficulty maintaining sleep.[3] It is FDA-approved for insomnia characterized by difficulties with sleep onset and/or sleep maintenance.[7] The starting dose of 10 mg can be increased to 20 mg if needed for efficacy, with dose adjustment based on response.[7]<br><br>Rationale for suvorexant over alternatives:<br><br>Mechanism suited to sleep maintenance: Orexin receptor antagonists reduce nocturnal awakenings and improve sleep continuity[3-4]<br><br>Safety profile: Lower risk of residual sedation compared to benzodiazepines and Z-drugs, which should be avoided when possible due to short- and long-term risks[4]<br><br>Evidence base: Demonstrated efficacy specifically for sleep maintenance insomnia in clinical trials[3]<br><br>Alternative considerations if suvorexant is not tolerated or effective: |

| | | |
|---|---|---|
| | Low-dose doxepin (3-6 mg at bedtime) has strong evidence for sleep maintenance insomnia[3][8]<br><br>Nonbenzodiazepine receptor agonists (zolpidem, eszopiclone) at lowest effective dose for shortest duration[8]<br><br>Additional recommendations:<br><br>Review sleep hygiene practices, though sleep hygiene education alone is insufficient as monotherapy[2]<br><br>Assess for other contributors to sleep fragmentation in Parkinson's disease (REM sleep behavior disorder, restless legs syndrome, nocturnal akinesia)<br><br>Monitor for medication side effects and efficacy at 2-4 week follow-up<br><br>Shared decision-making approach regarding benefits, risks, and costs of pharmacotherapy[1]<br><br>The goal is to use medication as a bridge or adjunct to CBT-I, which provides the most durable long-term benefit.[5][9] Sequential therapy combining behavioral treatment followed by medication (or vice versa) yields higher response and remission rates than either modality alone.[9] | |
| 16 | This assessment and plan will address excessive daytime sleepiness (EDS) in a patient with Parkinson's disease on dopaminergic therapy. The medical literature supports a stepwise approach: first optimizing dopaminergic medications (particularly reducing sedating agents like dopamine agonists if present), then considering wake-promoting agents such as modafinil, which has moderate-quality evidence for EDS in Parkinson's disease.[1-2] | A: 58-year-old male with 6-year history of idiopathic Parkinson's disease presenting with severe excessive daytime sleepiness (EDS) manifesting as involuntary sleep episodes during sedentary activities and near-miss events while driving. The patient's EDS is likely multifactorial, with contributing factors including: Dopaminergic medication effects (levodopa can contribute to somnolence)[3] Possible underlying sleep architecture |

| | | |
|---|---|---|
| | Assessment and Plan<br><br>Assessment:<br><br>58-year-old male with 6-year history of idiopathic Parkinson's disease presenting with severe excessive daytime sleepiness (EDS) manifesting as involuntary sleep episodes during sedentary activities and near-miss events while driving. Current medications include levodopa/carbidopa 100/25 mg 2 tablets four times daily, entacapone 200 mg four times daily, citalopram 20 mg daily, and mirabegron 50 mg daily.<br><br>The patient's EDS is likely multifactorial, with contributing factors including:<br><br>Dopaminergic medication effects (levodopa can contribute to somnolence)[3]<br><br>Possible underlying sleep architecture disruption from Parkinson's disease itself[1]<br><br>Potential contribution from citalopram (SSRIs can affect sleep quality)[4]<br><br>Need to exclude obstructive sleep apnea and other primary sleep disorders[3]<br><br>The severity of symptoms, particularly near-sleep episodes while driving, represents a significant safety concern requiring immediate intervention.<br><br>Plan:<br><br>Sleep Study Evaluation<br><br>Order polysomnography to exclude obstructive sleep apnea and other primary sleep disorders before initiating wake-promoting therapy, as these | disruption from Parkinson's disease itself[1] Potential contribution from citalopram (SSRIs can affect sleep quality)[4] Need to exclude obstructive sleep apnea and other primary sleep disorders[3]<br>Plan: Sleep Study Evaluation: Order polysomnography to exclude obstructive sleep apnea and other primary sleep disorders before initiating wake-promoting therapy, as these conditions must be addressed first[3]<br>If sleep apnea is identified, initiate CPAP therapy as this alone may improve EDS[3]<br><br>Medication Optimization<br>Review timing and dosing of levodopa/carbidopa to ensure adequate nighttime coverage while minimizing daytime sedation<br>Consider whether citalopram is contributing to sleep disturbance; evaluate risk-benefit of continuing versus alternative antidepressant if indicated[4]<br>Ensure good sleep hygiene practices[1][4]<br><br>Pharmacologic Treatment for EDS<br>Initiate modafinil 100 mg once daily in the morning, with plan to titrate to 200 mg daily after one week if tolerated and needed for symptom control[1-2]<br>Modafinil has moderate-quality evidence supporting its use for EDS in Parkinson's disease and is suggested by the American Academy of Sleep Medicine[2]<br><br>Common adverse effects include insomnia, nausea, headache, and dry mouth; monitor for these symptoms[1] |

| | |
|---|---|
| | conditions must be addressed first[3]<br><br>If sleep apnea is identified, initiate CPAP therapy as this alone may improve EDS[3]<br><br>Medication Optimization<br><br>Review timing and dosing of levodopa/carbidopa to ensure adequate nighttime coverage while minimizing daytime sedation<br><br>Consider whether citalopram is contributing to sleep disturbance; evaluate risk-benefit of continuing versus alternative antidepressant if indicated[4]<br><br>Ensure good sleep hygiene practices[1][4]<br><br>Pharmacologic Treatment for EDS<br><br>Initiate modafinil 100 mg once daily in the morning, with plan to titrate to 200 mg daily after one week if tolerated and needed for symptom control[1-2]<br><br>Modafinil has moderate-quality evidence supporting its use for EDS in Parkinson's disease and is suggested by the American Academy of Sleep Medicine[2]<br><br>Common adverse effects include insomnia, nausea, headache, and dry mouth; monitor for these symptoms[1]<br><br>Alternative agents if modafinil is ineffective or not tolerated include caffeine or solriamfetol[1][3][5]<br><br>Safety Counseling<br><br>Strict driving restriction until EDS is adequately controlled and confirmed by clinical reassessment<br><br>Educate patient about avoiding high-risk activities when sleepy |

| | | |
|---|---|---|
| | Consider referral to occupational therapy for safety assessment<br><br>Follow-up<br><br>Reassess in 2-4 weeks after polysomnography results and modafinil initiation<br><br>Use Epworth Sleepiness Scale to objectively track improvement<br><br>Adjust treatment based on response and tolerability<br><br>If modafinil provides insufficient benefit, consider sodium oxybate (though this requires careful monitoring given FDA black box warning for respiratory depression and potential to induce obstructive sleep apnea)[2][6] | |
| 17 | The assessment and plan will follow evidence-based guidelines for managing Parkinson's disease-related pain, with optimization of dopaminergic therapy as the first-line approach.[1] Given the temporal relationship between pain and motor symptoms, this appears to be fluctuation-related musculoskeletal pain, which should initially be addressed by adjusting the current levodopa regimen before considering additional analgesics.[1-3]<br><br>Assessment and Plan<br><br>Assessment:<br><br>64-year-old female with idiopathic Parkinson's disease (4-year duration) presenting with chronic musculoskeletal pain in shoulders and arms that worsens with motor symptom fluctuations, consistent with Parkinson's disease-related fluctuation-associated nociceptive pain. Currently managed with | A: 64-year-old female with idiopathic Parkinson's disease (4-year duration) presenting with chronic musculoskeletal pain in shoulders and arms that worsens with motor symptom fluctuations, consistent with Parkinson's disease-related fluctuation-associated nociceptive pain.<br>P: Optimize dopaminergic therapy as the first-line intervention for Parkinson's disease-related pain.[1-2] The current levodopa dosing schedule may be inadequate to maintain stable dopaminergic levels throughout the day, contributing to fluctuation-related pain.<br><br>Treatment Recommendation:<br><br>Increase levodopa/carbidopa to 25/100 mg, 2 tablets four times daily (total daily levodopa dose: 800 mg divided into more frequent dosing). This adjustment provides both increased total daily levodopa and more frequent |

| | | |
|---|---|---|
| | levodopa/carbidopa 100/25 mg, 1.5 tablets three times daily (total daily levodopa dose: 450 mg), and gabapentin 300 mg at bedtime. The temporal relationship between pain exacerbation and worsening Parkinson's symptoms suggests dopaminergic deficiency as a key contributor to her pain syndrome.[1]<br><br>The pain significantly impairs quality of life and responds variably to over-the-counter analgesics. Current gabapentin dosing is subtherapeutic for neuropathic pain management and unlikely to provide meaningful benefit for this pain phenotype.<br><br>Plan:<br><br>Optimize dopaminergic therapy as the first-line intervention for Parkinson's disease-related pain.[1-2] The current levodopa dosing schedule may be inadequate to maintain stable dopaminergic levels throughout the day, contributing to fluctuation-related pain.<br><br>Treatment Recommendation:<br><br>Increase levodopa/carbidopa to 25/100 mg, 2 tablets four times daily (total daily levodopa dose: 800 mg divided into more frequent dosing). This adjustment provides both increased total daily levodopa and more frequent dosing intervals to reduce motor fluctuations and associated pain.[1]<br><br>Rationale: Optimization of dopaminergic treatment should always be the initial step in managing Parkinson's disease-related chronic pain, as dopaminergic deficiency is often a key contributor to pain syndromes.[1] More frequent dosing intervals help maintain stable dopaminergic levels and reduce off-period symptoms, including fluctuation- | dosing intervals to reduce motor fluctuations and associated pain.[1]<br><br>Rationale: Optimization of dopaminergic treatment should always be the initial step in managing Parkinson's disease-related chronic pain, as dopaminergic deficiency is often a key contributor to pain syndromes.[1] More frequent dosing intervals help maintain stable dopaminergic levels and reduce off-period symptoms, including fluctuation-related musculoskeletal pain.[1] This approach is supported by current treatment algorithms that prioritize dopaminergic optimization before considering non-dopaminergic analgesics.[1]<br><br>Additional Considerations:<br><br>Discontinue gabapentin 300 mg at bedtime, as this dose is subtherapeutic and unlikely to provide benefit for her pain type<br><br>Monitor for hyperdopaminergic effects including dyskinesia, orthostatic hypotension, and neuropsychiatric symptoms with increased levodopa dosing[1]<br><br>Reassess pain severity in 2-4 weeks using a standardized pain scale<br><br>If pain persists despite optimized dopaminergic therapy, consider adding a COMT inhibitor (e.g., entacapone) or MAO-B inhibitor to further stabilize dopaminergic levels[1]<br><br>Non-pharmacological interventions including physical therapy, stretching exercises, and massage should be incorporated as adjunctive measures[2][4] |

| | | |
|---|---|---|
| | related musculoskeletal pain.[1] This approach is supported by current treatment algorithms that prioritize dopaminergic optimization before considering non-dopaminergic analgesics.[1]<br><br>Additional Considerations:<br><br>Discontinue gabapentin 300 mg at bedtime, as this dose is subtherapeutic and unlikely to provide benefit for her pain type<br><br>Monitor for hyperdopaminergic effects including dyskinesia, orthostatic hypotension, and neuropsychiatric symptoms with increased levodopa dosing[1]<br><br>Reassess pain severity in 2-4 weeks using a standardized pain scale<br><br>If pain persists despite optimized dopaminergic therapy, consider adding a COMT inhibitor (e.g., entacapone) or MAO-B inhibitor to further stabilize dopaminergic levels[1]<br><br>Non-pharmacological interventions including physical therapy, stretching exercises, and massage should be incorporated as adjunctive measures[2][4]<br><br>If pain remains refractory after dopaminergic optimization, consider trial of duloxetine or prolonged-release oxycodone/naloxone, though evidence for the latter is mixed[1-3][5]<br><br>Follow-up:<br><br>Schedule return visit in 2-4 weeks to assess treatment response, monitor for adverse effects, and adjust therapy as needed. | If pain remains refractory after dopaminergic optimization, consider trial of duloxetine or prolonged-release oxycodone/naloxone, though evidence for the latter is mixed[1-3][5]<br><br>Follow-up:<br><br>Schedule return visit in 2-4 weeks to assess treatment response, monitor for adverse effects, and adjust therapy as needed. |

| | | |
|---|---|---|
| 18 | The assessment and plan will address this patient's overactive bladder symptoms in the context of Parkinson's disease, with a treatment recommendation grounded in recent evidence. While behavioral therapy has shown noninferiority to pharmacotherapy in a 2025 trial, the patient's severe symptoms with multiple daily accidents and significant nocturia warrant initial pharmacological intervention.[1] The recommendation will focus on mirabegron as first-line therapy, given its efficacy in Parkinson's disease patients and favorable safety profile compared to antimuscarinics.[2-3]<br><br>Assessment and Plan<br><br>Assessment:<br><br>71-year-old male with 9-year history of idiopathic Parkinson's disease presenting with severe overactive bladder (OAB) symptoms characterized by urinary urgency and frequency every 2 hours, urge incontinence with multiple accidents, and bothersome nocturia disrupting sleep for both patient and spouse. OAB symptoms occur in 30-50% of patients with Parkinson's disease, with urgency affecting approximately 60%, nocturia 50%, and incontinence 40%.[1] These symptoms significantly impact quality of life and are associated with increased fall risk and potential for institutionalization.[1]<br><br>Current Parkinson's disease management includes levodopa/carbidopa 100/25 mg 2 tablets four times daily, entacapone 200 mg four times daily, and Laxaday 17 g daily for constipation. The severity of urinary symptoms—with accidents occurring multiple times and nocturia causing sleep disruption—warrants pharmacological intervention. | A: 71-year-old male with 9-year history of idiopathic Parkinson's disease presenting with severe overactive bladder (OAB) symptoms characterized by urinary urgency and frequency every 2 hours, urge incontinence with multiple accidents, and bothersome nocturia disrupting sleep for both patient and spouse. Plan: Initiate mirabegron 25 mg orally once daily, with plan to increase to 50 mg daily after 4-8 weeks if symptoms persist and medication is well-tolerated. Mirabegron, a β3-adrenergic receptor agonist, has demonstrated efficacy in treating OAB symptoms in Parkinson's disease patients in randomized controlled trials.[3] In a double-blind, placebo-controlled study of 117 Parkinson's disease patients with OAB, mirabegron significantly reduced OAB symptom scores at 4 and 8 weeks compared to placebo.[3] Meta-analysis of randomized trials shows that pharmacological treatment for OAB in Parkinson's disease significantly reduces micturition episodes per 24 hours (mean difference -1.33), nocturia episodes (mean difference -0.33), and urinary incontinence episodes (mean difference -0.72), while improving quality of life scores.[2]<br>Mirabegron is preferred over antimuscarinic agents (such as solifenacin or fesoterodine) in this patient for several important reasons. First, antimuscarinics carry risk of cognitive impairment, which is particularly concerning in Parkinson's disease patients.[4-5] Second, antimuscarinics can worsen constipation, which this patient already manages with daily Laxaday.[4][6] Third, mirabegron demonstrated acceptable adverse events in Parkinson's disease patients, with only mild increases in post-void residual |

| | | |
|---|---|---|
| | Plan:<br><br>Initiate mirabegron 25 mg orally once daily, with plan to increase to 50 mg daily after 4-8 weeks if symptoms persist and medication is well-tolerated.<br><br>Mirabegron, a β3-adrenergic receptor agonist, has demonstrated efficacy in treating OAB symptoms in Parkinson's disease patients in randomized controlled trials.[3] In a double-blind, placebo-controlled study of 117 Parkinson's disease patients with OAB, mirabegron significantly reduced OAB symptom scores at 4 and 8 weeks compared to placebo.[3] Meta-analysis of randomized trials shows that pharmacological treatment for OAB in Parkinson's disease significantly reduces micturition episodes per 24 hours (mean difference -1.33), nocturia episodes (mean difference -0.33), and urinary incontinence episodes (mean difference -0.72), while improving quality of life scores.[2]<br><br>Mirabegron is preferred over antimuscarinic agents (such as solifenacin or fesoterodine) in this patient for several important reasons. First, antimuscarinics carry risk of cognitive impairment, which is particularly concerning in Parkinson's disease patients.[4-5] Second, antimuscarinics can worsen constipation, which this patient already manages with daily Laxaday.[4][6] Third, mirabegron demonstrated acceptable adverse events in Parkinson's disease patients, with only mild increases in post-void residual urine volume and no significant difference in treatment-related adverse events compared to placebo.[3]<br><br>Additional management considerations:<br><br>Non-pharmacological measures: Recommend limiting fluid intake 2-3 | urine volume and no significant difference in treatment-related adverse events compared to placebo.[3]<br><br>Additional management considerations:<br>Non-pharmacological measures: Recommend limiting fluid intake 2-3 hours before bedtime and reducing caffeine consumption to help reduce nocturia and urgency[4]<br>Monitor for adverse effects: Watch for hypertension, urinary retention (though risk is low), and any worsening of Parkinson's disease symptoms<br>Optimize dopaminergic therapy: Current levodopa/carbidopa regimen may be contributing to or ameliorating bladder symptoms; no changes recommended at this time unless motor symptoms warrant adjustment[4]<br>Consider pelvic floor muscle training: If mirabegron provides insufficient benefit, behavioral therapy with pelvic floor muscle exercises has shown noninferiority to drug therapy and may be added as adjunctive treatment[1][7] |

| | | |
|---|---|---|
| | hours before bedtime and reducing caffeine consumption to help reduce nocturia and urgency[4]<br><br>Monitor for adverse effects: Watch for hypertension, urinary retention (though risk is low), and any worsening of Parkinson's disease symptoms<br><br>Optimize dopaminergic therapy: Current levodopa/carbidopa regimen may be contributing to or ameliorating bladder symptoms; no changes recommended at this time unless motor symptoms warrant adjustment[4]<br><br>Consider pelvic floor muscle training: If mirabegron provides insufficient benefit, behavioral therapy with pelvic floor muscle exercises has shown noninferiority to drug therapy and may be added as adjunctive treatment[1][7]<br><br>Follow-up: Reassess symptoms in 4-6 weeks using validated questionnaires (OAB Symptom Score) and voiding diary to objectively measure treatment response<br><br>Escalation options if inadequate response: Increase mirabegron to 50 mg daily, add behavioral therapy, or consider referral to urology for evaluation of intravesical botulinum toxin injection in refractory cases[4]<br><br>Rationale for starting dose: Initiating at 25 mg daily allows assessment of tolerability before escalating to the standard 50 mg daily dose, which was the dose used in the pivotal Parkinson's disease trial.[3] | |
| 19 | This assessment and plan will address refractory constipation in a patient with Parkinson's disease who has failed conservative measures and stimulant laxatives. The American Geriatrics Society | A: Constipation is a prevalent non-motor symptom in Parkinson's disease that can precede motor symptoms and significantly impacts quality of life.[3-4] This patient has appropriately trialed |

| | | |
|---|---|---|
| | recommends escalating to prescription agents like prucalopride when first-line therapies are insufficient, and recent evidence specifically demonstrates prucalopride's efficacy and safety in Parkinson's disease-related constipation.[1-2]<br><br>Assessment and Plan<br><br>Assessment:<br><br>75-year-old female with 8-year history of idiopathic Parkinson's disease presenting with refractory constipation despite conservative management. Currently experiencing bowel movements only every 3-4 days with associated bloating, discomfort, and straining, despite daily senna, high-fiber diet, and adequate fluid intake.<br><br>Current medications include levodopa/carbidopa 100/25 mg 1.5 tablets four times daily, rivastigmine patch 4.6 mg daily for cognitive symptoms, citalopram 10 mg daily, and senna 2 tablets at bedtime.<br><br>Constipation is a prevalent non-motor symptom in Parkinson's disease that can precede motor symptoms and significantly impacts quality of life.[3-4] This patient has appropriately trialed lifestyle modifications (fiber, hydration) and stimulant laxatives without adequate response, warranting escalation of therapy.<br><br>Plan:<br><br>Constipation Management:<br><br>Initiate prucalopride 1 mg orally once daily, with option to increase to 2 mg daily after 2-4 weeks if needed and tolerated. A recent randomized controlled | lifestyle modifications (fiber, hydration) and stimulant laxatives without adequate response, warranting escalation of therapy.<br><br>Plan: Constipation Management: Initiate prucalopride 1 mg orally once daily, with option to increase to 2 mg daily after 2-4 weeks if needed and tolerated. A recent randomized controlled trial demonstrated that prucalopride 2 mg daily significantly increased spontaneous bowel movements in Parkinson's disease patients with chronic constipation (mean increase of 3.2 weekly bowel movements vs 1.6 with placebo, p=0.02), with good tolerability and no worsening of motor symptoms. Given the patient's age, starting at the lower 1 mg dose is appropriate per clinical trial protocols in older adults.[2][5]<br><br>Continue senna 2 tablets at bedtime as adjunctive therapy. The American Geriatrics Society recommends combining stimulant laxatives with other agents as needed.[1]<br><br>Consider adding polyethylene glycol (macrogol) as an osmotic laxative if response to prucalopride is suboptimal, as this has demonstrated efficacy in Parkinson's disease and has minimal drug interactions.[4-5] |

| | |
|---|---|
| | trial demonstrated that prucalopride 2 mg daily significantly increased spontaneous bowel movements in Parkinson's disease patients with chronic constipation (mean increase of 3.2 weekly bowel movements vs 1.6 with placebo, p=0.02), with good tolerability and no worsening of motor symptoms. Given the patient's age, starting at the lower 1 mg dose is appropriate per clinical trial protocols in older adults.[2][5]<br><br>Continue senna 2 tablets at bedtime as adjunctive therapy. The American Geriatrics Society recommends combining stimulant laxatives with other agents as needed.[1]<br><br>Consider adding polyethylene glycol (macrogol) as an osmotic laxative if response to prucalopride is suboptimal, as this has demonstrated efficacy in Parkinson's disease and has minimal drug interactions.[4-5]<br><br>Monitoring:<br><br>Counsel patient and caregivers about potential side effects of prucalopride including nausea, diarrhea, and abdominal pain.[5]<br><br>Assess renal function given age-related considerations with prucalopride use.[5]<br><br>Follow up in 2-4 weeks to assess response and consider dose titration to 2 mg daily if bowel movement frequency remains suboptimal.<br><br>Parkinson's Disease Management:<br><br>Continue current levodopa/carbidopa regimen.<br><br>Continue rivastigmine patch and |

| | | |
|---|---|---|
| | citalopram as prescribed.<br><br>Note that optimizing dopaminergic therapy may have modest effects on gastrointestinal motility.[3]<br><br>Additional Considerations:<br><br>Ensure adequate hydration continues, particularly when initiating prucalopride.<br><br>Consider gastroenterology referral if constipation remains refractory to prucalopride and combination laxative therapy.[3]<br><br>Evaluate for potential pelvic floor dysfunction if symptoms persist, though this would require specialized assessment.[4] | |
| 20 | This assessment and plan addresses orthostatic hypotension in a patient with advanced Parkinson's disease, incorporating both non-pharmacological and pharmacological interventions. The treatment recommendation prioritizes midodrine as first-line therapy based on evidence from multiple sources demonstrating its efficacy and FDA approval for orthostatic hypotension.[1-5]<br><br>Assessment and Plan<br><br>Assessment:<br><br>73-year-old male with 10-year history of idiopathic Parkinson's disease presenting with symptomatic orthostatic hypotension manifesting as lightheadedness, dizziness upon standing, and near-syncope episodes, particularly severe in the morning after rising from bed.<br><br>Current medications include levodopa/carbidopa 100/25 mg (2 tablets five times daily), Sinemet CR 200/50 mg | A: 73-year-old male with 10-year history of idiopathic Parkinson's disease presenting with symptomatic orthostatic hypotension manifesting as lightheadedness, dizziness upon standing, and near-syncope episodes, particularly severe in the morning after rising from bed. The severity of symptoms, particularly the near-syncope episodes requiring immediate sitting to prevent falls, indicates significant functional impairment requiring intervention beyond non-pharmacological measures alone.<br><br>Plan: Non-pharmacological interventions:<br>Stand slowly from sitting or lying positions[7]<br>Elevate head of bed 30-45 degrees to reduce morning orthostatic hypotension[1][7]<br>Increase fluid intake to 2-2.5 liters daily[7]<br>Increase dietary salt intake (unless contraindicated by other conditions)[2][7] |

| | | |
|---|---|---|
| | at bedtime, safinamide 100 mg daily, rivastigmine patch 9.6 mg daily, and Laxadya 17 g as needed. The patient's dopaminergic therapy, particularly levodopa, may be contributing to or exacerbating orthostatic hypotension through both peripheral and central hypotensive mechanisms. Rivastigmine, while beneficial for cognitive symptoms, has been associated with increased risk of syncope and bradycardia in patients with dementia.[6]<br><br>The severity of symptoms, particularly the near-syncope episodes requiring immediate sitting to prevent falls, indicates significant functional impairment requiring intervention beyond non-pharmacological measures alone.<br><br>Plan:<br><br>Non-pharmacological interventions:<br><br>Stand slowly from sitting or lying positions[7]<br><br>Elevate head of bed 30-45 degrees to reduce morning orthostatic hypotension[1][7]<br><br>Increase fluid intake to 2-2.5 liters daily[7]<br><br>Increase dietary salt intake (unless contraindicated by other conditions)[2][7]<br><br>Avoid large meals that may worsen postprandial hypotension[1]<br><br>Consider compression stockings, though patient tolerance may be limited[1]<br><br>Pharmacological intervention:<br><br>Initiate midodrine 5 mg three times daily (morning, midday, and early afternoon | Avoid large meals that may worsen postprandial hypotension[1]<br>Consider compression stockings, though patient tolerance may be limited[1]<br><br>Pharmacological intervention:<br>Initiate midodrine 5 mg three times daily (morning, midday, and early afternoon doses), with the last dose no later than 4-5 hours before bedtime to minimize risk of supine hypertension. Midodrine is an FDA-approved alpha-1 adrenergic agonist that increases peripheral vascular resistance through vasoconstriction and has demonstrated significant improvement in standing systolic blood pressure and orthostatic symptoms in randomized controlled trials.[1-5] |

| | |
|---|---|
| | doses), with the last dose no later than 4-5 hours before bedtime to minimize risk of supine hypertension. Midodrine is an FDA-approved alpha-1 adrenergic agonist that increases peripheral vascular resistance through vasoconstriction and has demonstrated significant improvement in standing systolic blood pressure and orthostatic symptoms in randomized controlled trials.[1-5]<br><br>If symptoms persist after one week, may titrate midodrine to 10 mg three times daily. Maximum dose is 10 mg three times daily.[4]<br><br>Monitoring:<br><br>Monitor supine and standing blood pressure regularly, particularly supine blood pressure given risk of supine hypertension with midodrine[1][5][7]<br><br>Assess for common adverse effects including piloerection, scalp itching, and urinary retention[4]<br><br>Avoid midodrine within 3-5 hours of bedtime to prevent nocturnal supine hypertension[1][4]<br><br>Monitor for therapeutic response within 2-3 weeks<br><br>Alternative considerations if inadequate response:<br><br>If midodrine provides insufficient symptom control, consider adding droxidopa 100 mg three times daily (titrate up to 600 mg three times daily as needed) or fludrocortisone 0.1 mg daily (maximum 0.2 mg daily), though fludrocortisone requires monitoring of electrolytes and has less favorable long-term safety profile.[1-3][5][7] |

| | | |
|---|---|---|
| | Medication review:<br><br>Continue current Parkinson's disease regimen as dopaminergic therapy adjustment is not recommended solely for orthostatic hypotension management, as the benefits of motor symptom control typically outweigh the hypotensive effects. Continue rivastigmine for cognitive benefits despite potential contribution to orthostatic symptoms, as the cognitive benefits are established.[2][6] | |
| 21 | The assessment and plan will address fatigue in Parkinson's disease with a specific treatment recommendation. While evidence for fatigue management in PD is limited, rasagiline has the strongest support for reducing physical fatigue, and this patient is already on rasagiline 1 mg daily, which may be suboptimal for fatigue management based on available evidence.[1-3]<br><br>Assessment and Plan<br><br>Assessment:<br><br>62-year-old female with idiopathic Parkinson's disease (6-year duration) presenting with severe, persistent fatigue unrelieved by rest. Current medications include levodopa/carbidopa 100/25 mg four times daily, rasagiline 1 mg daily, and amantadine 100 mg three times daily. The patient describes overwhelming exhaustion that persists despite adequate sleep and significantly impairs her ability to perform usual activities and exercise.<br><br>Fatigue is a common and debilitating non-motor symptom affecting 40-50% of patients with Parkinson's disease.[3-4] This symptom significantly impacts quality of life and represents a leading cause of disability in PD.[3] The patient's current medication regimen includes | Assessment: Fatigue is a common and debilitating non-motor symptom affecting 40-50% of patients with Parkinson's disease.[3-4] This symptom significantly impacts quality of life and represents a leading cause of disability in PD.[3] The patient's current medication regimen includes rasagiline, which has demonstrated efficacy in reducing physical fatigue in PD, though evidence suggests potential dose-dependent effects.[1-3]<br>Plan: Primary recommendation: Increase rasagiline from 1 mg daily to 2 mg daily (maximum approved dose).<br><br>Rationale: High-quality evidence demonstrates that rasagiline is associated with reduced progression of physical fatigue in PD.[1-2] A large randomized controlled trial (N=1176) showed that rasagiline significantly slowed fatigue progression compared to placebo (SMD -0.27, 95% CI -0.39 to -0.16).[1-2] A pilot study specifically examining rasagiline for fatigue treatment showed significant improvement in fatigue scores at 1 mg daily, and the pivotal trials included both 1 mg and 2 mg daily dosing with combined benefit.[2-3] Rasagiline represents the intervention with the strongest evidence base for fatigue management in PD, and dose |

| | | |
|---|---|---|
| | rasagiline, which has demonstrated efficacy in reducing physical fatigue in PD, though evidence suggests potential dose-dependent effects.[1-3]<br><br>Differential considerations for fatigue:<br><br>Primary PD-related fatigue (most likely given clinical presentation)<br><br>Suboptimal dopaminergic therapy<br><br>Comorbid depression or apathy<br><br>Sleep disorders<br><br>Medication side effects<br><br>Plan:<br><br>Primary recommendation: Increase rasagiline from 1 mg daily to 2 mg daily (maximum approved dose).<br><br>Rationale: High-quality evidence demonstrates that rasagiline is associated with reduced progression of physical fatigue in PD.[1-2] A large randomized controlled trial (N=1176) showed that rasagiline significantly slowed fatigue progression compared to placebo (SMD -0.27, 95% CI -0.39 to -0.16).[1-2] A pilot study specifically examining rasagiline for fatigue treatment showed significant improvement in fatigue scores at 1 mg daily, and the pivotal trials included both 1 mg and 2 mg daily dosing with combined benefit.[2-3] Rasagiline represents the intervention with the strongest evidence base for fatigue management in PD, and dose optimization may provide additional benefit.[2][5-6]<br><br>Additional considerations:<br><br>Assess for comorbid depression, as | optimization may provide additional benefit.[2][5-6] |

| | | |
|---|---|---|
| | patients with depressive symptoms may respond differently to fatigue treatment[2]<br><br>Evaluate sleep quality and consider polysomnography if sleep disorder suspected<br><br>Continue current levodopa/carbidopa and amantadine regimen (note: amantadine has not shown benefit for fatigue in PD)[2]<br><br>Consider behavioral interventions, particularly structured exercise programs, as adjunctive therapy, though evidence remains limited[1][6-7]<br><br>Monitoring:<br><br>Reassess fatigue severity in 8-12 weeks using validated scale (Fatigue Severity Scale or Parkinson Fatigue Scale)<br><br>Monitor for adverse effects of increased rasagiline dose<br><br>If inadequate response, consider alternative pharmacological options such as methylphenidate 10 mg three times daily or referral to movement disorders specialist for further management[8]<br><br>Patient education:<br><br>Discuss realistic expectations, as complete resolution of fatigue may not be achievable<br><br>Encourage maintenance of physical activity as tolerated, as some patients report exercise alleviates fatigue[7]<br><br>Optimize sleep hygiene | |
| 22 | The following assessment and plan is based on evidence demonstrating that Lee Silverman Voice Treatment (LSVT | A: The patient's presentation is consistent with hypokinetic dysarthria, which affects approximately 89% of |

| | | |
|---|---|---|
| | LOUD) is the most effective speech therapy intervention for hypophonic dysarthria in Parkinson's disease, with multiple randomized controlled trials showing significant improvements in vocal loudness and communication effectiveness that are maintained for months after treatment.[1-3]<br><br>Assessment and Plan<br><br>Assessment:<br><br>55-year-old male with idiopathic Parkinson's disease (3-year duration) presenting with hypophonic dysarthria characterized by reduced vocal volume, hoarse voice quality, and monotone speech. Current symptoms significantly impair functional communication in workplace settings, with colleagues frequently requesting repetition and patient experiencing frustration with inability to project voice adequately during phone calls and meetings. Currently managed with levodopa/carbidopa 100/25 mg, 1.5 tablets three times daily, and citalopram 20 mg daily.<br><br>The patient's presentation is consistent with hypokinetic dysarthria, which affects approximately 89% of individuals with Parkinson's disease. Voice and speech impairments in Parkinson's disease include reduced loudness, weak or breathy voice quality, and monotone speech, which typically do not respond adequately to dopaminergic medications alone.[2][4-6]<br><br>Plan:<br><br>Referral for Lee Silverman Voice Treatment (LSVT LOUD) is recommended as the primary intervention for this patient's dysarthria. LSVT LOUD has the | individuals with Parkinson's disease. Voice and speech impairments in Parkinson's disease include reduced loudness, weak or breathy voice quality, and monotone speech, which typically do not respond adequately to dopaminergic medications alone.[2][4-6]<br><br>Plan: Referral for Lee Silverman Voice Treatment (LSVT LOUD) is recommended as the primary intervention for this patient's dysarthria. LSVT LOUD has the strongest evidence base among speech therapy interventions for Parkinson's disease-related voice disorders.[1-3] |

| | |
|---|---|
| | strongest evidence base among speech therapy interventions for Parkinson's disease-related voice disorders.[1-3]<br><br>The treatment protocol consists of:<br><br>16 individual sessions delivered over 4 consecutive weeks<br><br>Four 50-minute sessions per week (face-to-face or via telehealth)<br><br>Daily home practice: 5-10 minutes on treatment days, 15 minutes twice daily on non-treatment days[1]<br><br>LSVT LOUD specifically targets vocal loudness through high-intensity, high-effort exercises focused on the respiratory-laryngeal system. Multiple randomized controlled trials demonstrate that LSVT LOUD significantly increases vocal sound pressure level by 3-9 dB across various speech tasks, with improvements maintained for up to 24 months post-treatment. A recent pragmatic trial showed that LSVT LOUD reduced voice handicap index scores by 8.0 points compared to no therapy and by 9.6 points compared to standard NHS speech therapy at 3 months.[1-3]<br><br>The treatment works by addressing the reduced perception of effort characteristic of Parkinson's disease, teaching patients to recalibrate their internal sense of "normal" loudness and maintain increased vocal effort. Neuroimaging studies demonstrate that LSVT LOUD induces cortical activation changes, particularly in superior temporal regions, which correlate with improved speech intelligibility.[4][7]<br><br>Continue current Parkinson's disease medications without adjustment at this time, as the patient's motor symptoms |

| | | |
|---|---|---|
| | appear adequately controlled on levodopa/carbidopa. Speech and voice problems in Parkinson's disease typically require behavioral intervention rather than medication adjustment.[6][8]<br><br>Expected outcomes: Improvements in vocal loudness, speech intelligibility, voice quality, and patient-reported communication effectiveness. Secondary benefits may include improvements in facial expression and swallowing function. The patient should be counseled that LSVT LOUD requires significant physical and mental effort during the intensive treatment period, but this high-intensity approach is essential for achieving sustained therapeutic benefit.[4][9]<br><br>Follow-up: Reassess speech and communication function 1 month and 6 months post-treatment completion to monitor maintenance of treatment gains and determine need for periodic refresher sessions. | |
| 23 | The following assessment and plan addresses bothersome drooling in a patient with advanced Parkinson's disease. The treatment recommendation is based on evidence demonstrating efficacy and safety of botulinum toxin injections for sialorrhea in Parkinson's disease, with multiple randomized controlled trials supporting this approach.<br>Lancet<br>+ 3<br>[1-4]<br><br>Assessment and Plan<br><br>Assessment:<br><br>66-year-old female with 10-year history of idiopathic Parkinson's disease presenting with bothersome sialorrhea unrelated to medication wearing-off | Assessment: Sialorrhea affects up to 74% of patients with Parkinson's disease and results from inefficient elimination of oral saliva rather than increased production. This symptom can cause significant embarrassment, social stigma, perioral skin excoriation, and aspiration risk.<br>Plan:<br>Botulinum toxin injection into the parotid and submandibular glands is recommended as first-line therapy for this patient's sialorrhea. Specifically, incobotulinumtoxinA 100 units total dose (divided between bilateral parotid and submandibular glands) is the committed treatment recommendation.<br>IncobotulinumtoxinA 100 units has demonstrated significant reduction in unstimulated salivary flow rate and |

| | | |
|---|---|---|
| | periods. Current antiparkinsonian regimen includes levodopa/carbidopa 100/25 mg two tablets five times daily, Sinemet CR 200/50 mg at bedtime, and safinamide 100 mg daily. Also taking citalopram 20 mg daily and melatonin 10 mg at bedtime. No prior treatment attempted for sialorrhea.<br><br>Sialorrhea affects up to 74% of patients with Parkinson's disease and results from inefficient elimination of oral saliva rather than increased production. This symptom can cause significant embarrassment, social stigma, perioral skin excoriation, and aspiration risk.<br>Lancet<br>+ 1<br>[1][3]<br><br>Plan:<br><br>Botulinum toxin injection into the parotid and submandibular glands is recommended as first-line therapy for this patient's sialorrhea. Specifically, incobotulinumtoxinA 100 units total dose (divided between bilateral parotid and submandibular glands) is the committed treatment recommendation.<br>AAN<br>[2]<br><br>IncobotulinumtoxinA 100 units has demonstrated significant reduction in unstimulated salivary flow rate and improvement in patient-reported outcomes at 4 weeks post-injection, with therapeutic effects lasting approximately 16 weeks. This represents FDA-approved therapy for chronic sialorrhea in adults. The treatment is well-tolerated with the most common adverse events being mild to moderate dry mouth (2.7%) and dysphagia (0% at this dose).<br>AAN<br>+ 1 | improvement in patient-reported outcomes at 4 weeks post-injection, with therapeutic effects lasting approximately 16 weeks. This represents FDA-approved therapy for chronic sialorrhea in adults. The treatment is well-tolerated with the most common adverse events being mild to moderate dry mouth (2.7%) and dysphagia (0% at this dose).<br><br>Alternative botulinum toxin formulations with evidence for sialorrhea include rimabotulinumtoxinB 2500-3500 units, which showed significant efficacy with onset as early as 1 week post-injection. However, incobotulinumtoxinA is preferred given its FDA approval specifically for this indication and favorable safety profile at the 100-unit dose.<br><br>Oral anticholinergic agents such as glycopyrrolate (1 mg three times daily) represent an alternative approach but carry risks of systemic anticholinergic effects including dry mouth, constipation, urinary retention, and potential cognitive impairment—particularly problematic in patients with Parkinson's disease. Given these concerns and the patient's advanced disease duration, botulinum toxin injection is the superior choice. |

| | | |
|---|---|---|
| | [2][5]<br><br>Alternative botulinum toxin formulations with evidence for sialorrhea include rimabotulinumtoxinB 2500-3500 units, which showed significant efficacy with onset as early as 1 week post-injection. However, incobotulinumtoxinA is preferred given its FDA approval specifically for this indication and favorable safety profile at the 100-unit dose.<br>JAMA<br>[3]<br><br>Oral anticholinergic agents such as glycopyrrolate (1 mg three times daily) represent an alternative approach but carry risks of systemic anticholinergic effects including dry mouth, constipation, urinary retention, and potential cognitive impairment—particularly problematic in patients with Parkinson's disease. Given these concerns and the patient's advanced disease duration, botulinum toxin injection is the superior choice.<br>Lancet<br>+ 1<br>[1][6]<br><br>The patient should be counseled that injections will need to be repeated approximately every 3-4 months based on symptom recurrence. Referral to a neurologist or other physician trained in botulinum toxin administration for salivary gland injections should be arranged.<br><br>Continue current antiparkinsonian medications without adjustment, as the sialorrhea is not related to wearing-off periods. | |
| 24 | This assessment and plan will address oropharyngeal dysphagia in advanced Parkinson's disease, incorporating evidence-based diagnostic and | A: Dysphagia occurs in over 80% of patients with Parkinson's disease and represents a significant risk factor for aspiration pneumonia, malnutrition, |

| | | |
|---|---|---|
| | therapeutic approaches. The literature supports instrumental swallowing assessment as the gold standard and identifies expiratory muscle strength training as having the strongest evidence for treatment efficacy, with emerging data supporting neurostimulation approaches.<br>Lancet<br>+ 6<br>[1-7]<br><br>Assessment and Plan<br><br>Assessment:<br><br>75-year-old male with 11-year history of idiopathic Parkinson's disease presenting with oropharyngeal dysphagia characterized by coughing/choking with liquids, need for modified food textures (small pieces), and avoidance of tough or dry foods. Symptoms are not temporally related to medication wearing-off periods, suggesting persistent dysphagia independent of dopaminergic fluctuations.<br><br>Current antiparkinsonian regimen: levodopa/carbidopa 100/25 mg 2 tablets four times daily every 4 hours, levodopa/carbidopa CR 200/50 mg at bedtime, safinamide 100 mg daily.<br><br>Dysphagia occurs in over 80% of patients with Parkinson's disease and represents a significant risk factor for aspiration pneumonia, malnutrition, and mortality.<br>Dysphagia<br>[8]<br>The patient's symptoms suggest both oral phase dysfunction (requiring food modification) and pharyngeal phase impairment (liquid aspiration risk).<br><br>Plan: | and mortality.<br>The patient's symptoms suggest both oral phase dysfunction (requiring food modification) and pharyngeal phase impairment (liquid aspiration risk).<br><br>Plan: Instrumental Swallowing Assessment<br>Refer for videofluoroscopic swallowing study (VFSS), which is the gold standard for evaluating oropharyngeal dysfunction in Parkinson's disease<br>VFSS will assess aspiration risk, identify specific swallowing phase impairments, and guide targeted therapy<br>Consider fiberoptic endoscopic evaluation of swallowing (FEES) as alternative if VFSS unavailable<br>Speech-Language Pathology Referral |

| | |
|---|---|
| | Instrumental Swallowing Assessment<br><br>Refer for videofluoroscopic swallowing study (VFSS), which is the gold standard for evaluating oropharyngeal dysfunction in Parkinson's disease<br>Lancet<br>+ 1<br>[1-2]<br><br>VFSS will assess aspiration risk, identify specific swallowing phase impairments, and guide targeted therapy<br><br>Consider fiberoptic endoscopic evaluation of swallowing (FEES) as alternative if VFSS unavailable<br>Current Opinion in Neurology<br>[2]<br><br>Speech-Language Pathology Referral<br><br>Initiate expiratory muscle strength training (EMST) as primary rehabilitative intervention<br><br>EMST has Class I evidence demonstrating improved swallow safety (penetration-aspiration scores) and hyolaryngeal movement in Parkinson's disease<br>AAN<br>[4]<br><br>A 4-week EMST protocol (5 days/week, 20 minutes/day) significantly reduces dysphagia severity with sustained effects at 3 months<br>Movement Disorders<br>[3]<br><br>EMST specifically improves pharyngeal residue clearance and overall swallowing efficiency<br>Movement Disorders<br>[3]<br><br>Additional therapies to consider based on |

| | |
|---|---|
| | VFSS findings: video-assisted swallowing therapy, compensatory strategies, and postural techniques<br>Lancet<br>+ 1<br>[1][9]<br><br>Adjunctive Neurostimulation Therapies<br><br>Consider referral for neuromuscular electrical stimulation (NMES) or repetitive transcranial magnetic stimulation (rTMS) as adjunctive therapy<br><br>Meta-analyses demonstrate medium-to-large effect sizes for neurostimulation approaches in Parkinson's disease dysphagia<br>Neurogastroenterology and Motility<br>+ 1<br>[5][7]<br><br>NMES shows significant improvement in standardized swallowing assessments and penetration-aspiration scores<br>Neuropsychiatric Disease and Treatment<br>[7]<br><br>Medication Optimization<br><br>Current regimen appears optimized with continuous dopaminergic coverage (regular levodopa plus CR formulation, MAO-B inhibitor)<br><br>While dopaminergic effects on dysphagia remain controversial, continuous dopaminergic stimulation may benefit swallowing function<br>Lancet<br>+ 2<br>[1][10-11]<br><br>Monitor for any temporal relationship between medication timing and swallowing symptoms despite current report of no wearing-off correlation |

| | | |
|---|---|---|
| | Dietary Modifications and Safety<br><br>Continue current texture modifications (small pieces, avoiding tough/dry foods)<br><br>Formal dysphagia diet recommendations pending VFSS results<br><br>Consider thickened liquids if thin liquid aspiration confirmed on instrumental assessment<br><br>Nutritional assessment to ensure adequate caloric intake given food avoidance patterns<br><br>Multidisciplinary Follow-up<br><br>Re-evaluate swallowing function in 3 months following initiation of EMST<br><br>Monitor for signs of aspiration pneumonia (fever, cough, respiratory symptoms)<br><br>Consider gastroenterology referral if esophageal dysphagia symptoms develop, as esophageal dysmotility occurs in approximately 67% of Parkinson's disease patients<br>Lancet<br>[1]<br><br>Patient and Caregiver Education<br><br>Counsel on aspiration pneumonia risk and warning signs<br><br>Emphasize importance of adherence to swallowing therapy protocols<br><br>Discuss safe swallowing strategies pending formal speech-language pathology evaluation | |
| 25 | This assessment and plan will address medication-refractory tremor in a patient | A: Tremor is often less responsive to levodopa than bradykinesia and |

| | | |
|---|---|---|
| | with advanced Parkinson's disease experiencing significant functional impairment despite optimized dopaminergic therapy. The evidence supports that tremor can be less responsive to levodopa than other motor symptoms, and this patient's current regimen may benefit from optimization with adjunctive therapy targeting motor fluctuations.<br>NEJM<br>+ 2<br>[1-3]<br><br>Assessment and Plan<br><br>Assessment:<br><br>67-year-old male with idiopathic Parkinson's disease for 7 years presenting with functionally disabling tremor despite treatment with levodopa/carbidopa 100/25 mg, 1.5 tablets four times daily (total daily levodopa dose 600 mg), and entacapone 200 mg four times daily. The patient demonstrates significant functional impairment with activities of daily living, specifically eating, requiring assistance or adaptive utensils due to severe tremor affecting hand dexterity and strength.<br><br>Tremor is often less responsive to levodopa than bradykinesia and rigidity, and this patient appears to have tremor-predominant symptoms that are inadequately controlled despite a substantial levodopa regimen.<br>NEJM<br>[1]<br>The current medication schedule suggests possible motor fluctuations given the 4-hour dosing interval. While the patient is already on a COMT inhibitor (entacapone), opicapone is a once-daily COMT inhibitor that may provide more consistent coverage for motor | rigidity, and this patient appears to have tremor-predominant symptoms that are inadequately controlled despite a substantial levodopa regimen.<br>The current medication schedule suggests possible motor fluctuations given the 4-hour dosing interval. Plan: Switch from entacapone 200 mg four times daily to opicapone 50 mg once daily at bedtime (at least one hour before or after levodopa/carbidopa doses). Continue current levodopa/carbidopa 100/25 mg, 1.5 tablets four times daily. Rationale: Opicapone provides once-daily COMT inhibition and is indicated for adjunctive treatment in patients with Parkinson's disease experiencing "off" episodes. |

| | |
|---|---|
| | fluctuations and is FDA-approved as adjunctive treatment to levodopa/carbidopa in patients experiencing "off" episodes.<br>Lancet<br>+ 1<br>[2][4]<br><br>The patient's functional impairment with eating represents a significant quality of life issue. While adaptive feeding utensils can provide some benefit, optimizing pharmacologic management remains the priority for tremor control.<br>American Journal of Occupational Therapy<br>+ 1<br>[5-6]<br><br>Plan:<br><br>Switch from entacapone 200 mg four times daily to opicapone 50 mg once daily at bedtime (at least one hour before or after levodopa/carbidopa doses). Continue current levodopa/carbidopa 100/25 mg, 1.5 tablets four times daily.<br><br>Rationale: Opicapone provides once-daily COMT inhibition and is indicated for adjunctive treatment in patients with Parkinson's disease experiencing "off" episodes.<br>Lancet<br>+ 1<br>[2][4]<br>This simplification may improve medication adherence while potentially providing more consistent dopaminergic coverage throughout the day. Meta-analysis demonstrates that COMT inhibitors effectively reduce off time in patients with motor fluctuations, and opicapone's pharmacokinetic profile allows for once-daily dosing separate from levodopa administration.<br>Lancet |

| | | |
|---|---|---|
| | [2]<br><br>Additional recommendations:<br><br>Ensure levodopa doses are taken at least 1 hour separated from protein-containing meals to optimize absorption<br>Lancet<br>[2]<br><br>Consider trial of adaptive feeding utensils (weighted spoon with standard handle or active tremor cancellation device) as adjunctive non-pharmacologic intervention<br>American Journal of Occupational Therapy<br>[5]<br><br>Referral to occupational therapy for assessment and training with adaptive equipment<br><br>Monitor for dyskinesia or other dopaminergic side effects with medication adjustment<br><br>If tremor remains refractory to optimized medical management after 4-6 weeks, consider referral to movement disorders specialist for evaluation of advanced therapies including deep brain stimulation, which can effectively reduce medication-refractory tremor<br>JAMA<br>[7]<br><br>Follow-up in 4-6 weeks to assess tremor control, functional status with eating, and medication tolerability. | |
| 26 | This patient presents with motor fluctuations characteristic of advanced Parkinson's disease, with prolonged morning off time significantly impacting activities of daily living. The evidence supports adding a COMT inhibitor to his current regimen, with opicapone being | A: The patient demonstrates functional impairment in activities of daily living, with morning dressing time increased from 5 minutes to 30 minutes, requiring assistance with shoes and socks due to balance difficulties and bradykinesia. This represents classic |

| | | |
|---|---|---|
| | the preferred agent given its once-daily dosing and efficacy in reducing off time.[1-3]<br><br>Assessment and Plan<br><br>Assessment:<br><br>69-year-old male with idiopathic Parkinson's disease (8-year duration) presenting with significant motor fluctuations manifesting as prolonged morning off time. Currently on levodopa/carbidopa 100/25 mg, 2 tablets four times daily every 3 hours (total daily levodopa dose 800 mg), rasagiline 1 mg daily, mirabegron 50 mg daily, and Laxaday 17 g as needed.<br><br>The patient demonstrates functional impairment in activities of daily living, with morning dressing time increased from 5 minutes to 30 minutes, requiring assistance with shoes and socks due to balance difficulties and bradykinesia. This represents classic end-of-dose wearing off with early morning off symptoms, consistent with the complex phase of Parkinson's disease.[2]<br><br>His current regimen includes appropriate levodopa dosing with MAO-B inhibitor adjunctive therapy. However, the persistent motor fluctuations despite optimized oral levodopa frequency (every 3 hours) and rasagiline therapy indicate need for additional adjunctive treatment to extend levodopa benefit and reduce off time.[1][3]<br><br>Plan:<br><br>Add opicapone 50 mg once daily at bedtime as adjunctive therapy to current levodopa/carbidopa regimen.[1-4]<br><br>Rationale: Opicapone is a once-daily | end-of-dose wearing off with early morning off symptoms, consistent with the complex phase of Parkinson's disease.[2]<br><br>Plan: Add opicapone 50 mg once daily at bedtime as adjunctive therapy to current levodopa/carbidopa regimen.[1-4]<br>Rationale: Opicapone is a once-daily COMT inhibitor that has demonstrated efficacy in reducing off time in patients with Parkinson's disease experiencing motor fluctuations. It is FDA-approved as adjunctive treatment to levodopa/carbidopa in patients experiencing "off" episodes. Recent evidence confirms opicapone is efficacious for treating motor fluctuations in patients already on levodopa therapy. The once-daily dosing offers practical advantages over entacapone (which requires dosing with each levodopa dose) and may improve medication adherence.[1-5] |

| | |
|---|---|
| | COMT inhibitor that has demonstrated efficacy in reducing off time in patients with Parkinson's disease experiencing motor fluctuations. It is FDA-approved as adjunctive treatment to levodopa/carbidopa in patients experiencing "off" episodes. Recent evidence confirms opicapone is efficacious for treating motor fluctuations in patients already on levodopa therapy. The once-daily dosing offers practical advantages over entacapone (which requires dosing with each levodopa dose) and may improve medication adherence.[1-5]<br><br>COMT inhibitors work by blocking the degradation of levodopa, thereby prolonging its duration of action and extending on time. This mechanism directly addresses the patient's early morning off symptoms and end-of-dose wearing off.[1][6]<br><br>Continue current medications:<br><br>Levodopa/carbidopa 100/25 mg, 2 tablets four times daily every 3 hours<br><br>Rasagiline 1 mg once daily<br><br>Mirabegron 50 mg once daily<br><br>Laxaday 17 g once daily as needed<br><br>Monitoring and counseling:<br><br>Counsel patient that opicapone may increase dyskinesia; if this occurs, levodopa dose reduction may be necessary[1]<br><br>Monitor for other potential dopaminergic side effects including nausea, orthostatic hypotension, hallucinations, or behavioral changes[1] |

| | | |
|---|---|---|
| | Advise patient to take opicapone at bedtime, at least 1 hour before or after levodopa doses<br><br>Separate opicapone from protein-containing meals by at least 1 hour to optimize absorption[2]<br><br>Follow up in 4-6 weeks to assess response (reduction in morning off time and improvement in dressing independence)<br><br>Additional considerations:<br><br>Recommend referral to physical therapy for gait and balance training to address functional limitations with dressing and reduce fall risk[6-7]<br><br>If inadequate response to opicapone after appropriate trial, consider additional adjunctive therapies or evaluation for advanced treatments (deep brain stimulation, continuous subcutaneous levodopa, or levodopa-carbidopa intestinal gel) at a specialty center[1][3][6] | |
| 27 | This assessment and plan will focus on optimizing pharmacologic management for motor fluctuations and addressing functional decline through physical therapy referral. The patient is already on a comprehensive regimen including levodopa/carbidopa, entacapone, and safinamide, but her functional limitations suggest inadequate control of motor symptoms. The medical literature supports adding opicapone as the next step, given its efficacy in reducing off-time when added to existing COMT inhibitor therapy, along with physical therapy to specifically address balance and activities of daily living.[1-5]<br><br>Assessment and Plan<br><br>Assessment: | A: Despite this comprehensive regimen including levodopa, a COMT inhibitor (entacapone), and an MAO-B inhibitor (safinamide), the patient demonstrates persistent motor fluctuations with bradykinesia significantly impairing function. Her fear of falling and need for assistance with bathing indicate both motor symptom inadequacy and safety concerns that warrant intervention.<br><br>Plan: Medication Adjustment:<br>Add opicapone 50 mg once daily at bedtime, and discontinue entacapone. Opicapone is a once-daily COMT inhibitor that has demonstrated efficacy in reducing off-time in patients with Parkinson's disease experiencing motor fluctuations.[1-3] In the |

| | | |
|---|---|---|
| | 72-year-old female with 10-year history of idiopathic Parkinson's disease presenting with progressive functional decline manifesting as difficulty with activities of daily living, specifically bathing. She reports fear of losing balance in the slippery tub and exhaustion from slow movements during hygiene tasks, now requiring daughter's assistance several times weekly.<br><br>Current medication regimen includes:<br><br>Levodopa/carbidopa 100/25 mg, 2 tablets four times daily every 3 hours (total daily levodopa dose: 800 mg)<br><br>Entacapone 200 mg four times daily<br><br>Safinamide 100 mg once daily<br><br>Laxaday 17 g once daily<br><br>Despite this comprehensive regimen including levodopa, a COMT inhibitor (entacapone), and an MAO-B inhibitor (safinamide), the patient demonstrates persistent motor fluctuations with bradykinesia significantly impairing function. Her fear of falling and need for assistance with bathing indicate both motor symptom inadequacy and safety concerns that warrant intervention.<br><br>Plan:<br><br>Medication Adjustment:<br><br>Add opicapone 50 mg once daily at bedtime, and discontinue entacapone. Opicapone is a once-daily COMT inhibitor that has demonstrated efficacy in reducing off-time in patients with Parkinson's disease experiencing motor fluctuations.[1-3] In the International Parkinson and Movement Disorder | International Parkinson and Movement Disorder Society evidence-based review, opicapone was assessed as efficacious for treating motor fluctuations.[3] Opicapone offers the advantage of once-daily dosing compared to entacapone's four-times-daily regimen, which may improve medication adherence and provide more consistent COMT inhibition throughout the day.[2][6]<br>The starting dose of opicapone is 50 mg once daily, taken at bedtime at least one hour before or after levodopa/carbidopa.[6-7] This can be titrated to 25 mg daily if tolerability issues arise, though 50 mg is the standard therapeutic dose.<br><br>Physical Therapy Referral:<br>Refer for comprehensive physical therapy evaluation with focus on gait and balance training. Exercise interventions including gait and balance training, progressive resistance training, and strength training have demonstrated benefit for motor symptoms, balance, gait, and function in Parkinson's disease.[4] The American Geriatrics Society recommends optimizing exercise, strengthening, balance, and physical therapy at time of diagnosis and throughout the course of care.[5] Physical therapy can provide strategies to improve safety during bathing and other activities of daily living, potentially reducing fall risk and improving independence.[4]<br><br>Occupational Therapy Consideration:<br>Consider occupational therapy consultation to assess home safety modifications, particularly bathroom adaptations (grab bars, non-slip mats, shower chair) and to develop compensatory strategies for hygiene tasks.[4] |

| | |
|---|---|
| | Society evidence-based review, opicapone was assessed as efficacious for treating motor fluctuations.[3] Opicapone offers the advantage of once-daily dosing compared to entacapone's four-times-daily regimen, which may improve medication adherence and provide more consistent COMT inhibition throughout the day.[2][6]<br><br>The starting dose of opicapone is 50 mg once daily, taken at bedtime at least one hour before or after levodopa/carbidopa.[6-7] This can be titrated to 25 mg daily if tolerability issues arise, though 50 mg is the standard therapeutic dose.<br><br>Physical Therapy Referral:<br><br>Refer for comprehensive physical therapy evaluation with focus on gait and balance training. Exercise interventions including gait and balance training, progressive resistance training, and strength training have demonstrated benefit for motor symptoms, balance, gait, and function in Parkinson's disease.[4] The American Geriatrics Society recommends optimizing exercise, strengthening, balance, and physical therapy at time of diagnosis and throughout the course of care.[5] Physical therapy can provide strategies to improve safety during bathing and other activities of daily living, potentially reducing fall risk and improving independence.[4]<br><br>Occupational Therapy Consideration:<br><br>Consider occupational therapy consultation to assess home safety modifications, particularly bathroom adaptations (grab bars, non-slip mats, shower chair) and to develop compensatory strategies for hygiene tasks.[4] |

| | | |
|---|---|---|
| | Follow-up:<br><br>Schedule follow-up in 4-6 weeks to assess response to opicapone, monitor for adverse effects (dyskinesia, orthostatic hypotension, hallucinations), and evaluate functional improvement with physical therapy. If inadequate response to medication optimization and therapy interventions, consider referral to movement disorders specialist for evaluation of device-aided therapies. | |
| 28 | This assessment and plan addresses progressive micrographia in a patient with established Parkinson's disease on levodopa/carbidopa and rasagiline. The evidence indicates that progressive micrographia (where letters shrink during writing) is less responsive to levodopa optimization than consistent micrographia, making intensive amplitude training the most evidence-based intervention.[1-3] The treatment recommendation will focus on physical therapy with specific handwriting rehabilitation.<br><br>Assessment and Plan<br><br>Assessment:<br><br>63-year-old male with 5-year history of idiopathic Parkinson's disease presenting with progressive micrographia—handwriting that starts legible but progressively shrinks and becomes illegible during writing tasks. Currently managed with levodopa/carbidopa 100/25 mg, 2 tablets three times daily (total daily levodopa dose 600 mg), and rasagiline 1 mg daily.<br><br>The patient's description of letters that "start out legible but quickly shrink" is characteristic of progressive micrographia, a manifestation of the | Assessment:<br>The patient's description of letters that "start out legible but quickly shrink" is characteristic of progressive micrographia, a manifestation of the sequence effect in bradykinesia.[1-2] This pattern differs from consistent micrographia (uniformly small writing) and has distinct neural correlates, involving dysfunction of the basal ganglia motor circuit combined with disconnections between the rostral supplementary motor area, rostral cingulate motor area, and cerebellum.[1] Importantly, progressive micrographia demonstrates limited responsiveness to levodopa optimization, with studies showing that while levodopa improves writing speed, the decremental features of progressive micrographia remain largely unaltered.[1-2]<br><br>Plan:<br>Referral for intensive amplitude-focused handwriting training with physical or occupational therapy. The evidence demonstrates that intensive amplitude training (30 minutes daily, 5 days per week for 6 weeks) produces significant improvements in writing size with consolidation, automatization, and retention of gains after training.[3] This approach has shown effect sizes of |

| | | |
|---|---|---|
| | sequence effect in bradykinesia.[1-2] This pattern differs from consistent micrographia (uniformly small writing) and has distinct neural correlates, involving dysfunction of the basal ganglia motor circuit combined with disconnections between the rostral supplementary motor area, rostral cingulate motor area, and cerebellum.[1] Importantly, progressive micrographia demonstrates limited responsiveness to levodopa optimization, with studies showing that while levodopa improves writing speed, the decremental features of progressive micrographia remain largely unaltered.[1-2]<br><br>Plan:<br><br>Referral for intensive amplitude-focused handwriting training with physical or occupational therapy. The evidence demonstrates that intensive amplitude training (30 minutes daily, 5 days per week for 6 weeks) produces significant improvements in writing size with consolidation, automatization, and retention of gains after training.[3] This approach has shown effect sizes of 7-17% improvement in amplitude with benefits maintained at 6-week follow-up and transfer to daily life writing tasks.[3]<br><br>Specific treatment recommendation: Physical therapy or occupational therapy consultation for amplitude-based handwriting rehabilitation program consisting of 30 minutes of daily practice, 5 days per week for 6 weeks, focused on increasing writing size through targeted exercises with visual feedback (such as writing within progressively larger target zones).[3]<br><br>The patient should be counseled that while amplitude training effectively increases writing size, there may be | 7-17% improvement in amplitude with benefits maintained at 6-week follow-up and transfer to daily life writing tasks.[3] |

| | | |
|---|---|---|
| | trade-offs in writing speed and fluency during the learning phase.[4] However, the improvements in size are consolidated and retained over time.[3] Attention-based strategies during writing may provide additional benefit by recruiting the anterior putamen and dorsolateral prefrontal cortex.[1]<br><br>Continue current dopaminergic regimen (levodopa/carbidopa 100/25 mg, 2 tablets three times daily, and rasagiline 1 mg daily) as the patient's overall motor symptoms appear adequately controlled. Given that progressive micrographia shows limited response to levodopa dose adjustments, medication optimization is not the primary intervention at this time.[1-2]<br><br>Follow-up in 8-10 weeks to assess response to handwriting rehabilitation and reassess overall motor symptom control. | |
| 29 | This assessment and plan addresses motor fluctuations and mood symptoms in a patient with advanced Parkinson's disease experiencing "off" episodes despite optimized levodopa therapy. The treatment recommendation is supported by evidence showing efficacy of COMT inhibitors as adjunctive therapy for motor fluctuations, with opicapone specifically approved for this indication and demonstrating once-daily dosing convenience.<br>NEJM<br>+ 3<br>[1-4]<br><br>Assessment and Plan<br><br>Assessment:<br><br>68-year-old female with 9-year history of idiopathic Parkinson's disease experiencing motor fluctuations and | A: Motor symptoms: Patient demonstrates classic signs of advancing Parkinson's disease with inadequate tremor control and balance instability limiting previously enjoyed activities (knitting, gardening, painting). The tremor interfering with fine motor tasks and the balance issues suggest suboptimal symptom control, consistent with motor fluctuations that typically emerge after approximately 4 years of levodopa therapy.<br>Her current levodopa dosing interval of every 3 hours reflects attempts to manage wearing-off effects.<br>Mood symptoms: Patient exhibits depression related to loss of meaningful activities and sense of identity. Depression is common in Parkinson's disease and may be exacerbated by motor fluctuations.<br>Plan: Add opicapone 50 mg once daily at bedtime as adjunctive therapy to |

| | | |
|---|---|---|
| | functional decline despite current therapy with levodopa/carbidopa 100/25 mg 1.5 tablets four times daily (total daily levodopa dose 600 mg), selegiline 5 mg twice daily, and citalopram 20 mg daily.<br><br>Motor symptoms: Patient demonstrates classic signs of advancing Parkinson's disease with inadequate tremor control and balance instability limiting previously enjoyed activities (knitting, gardening, painting). The tremor interfering with fine motor tasks and the balance issues suggest suboptimal symptom control, consistent with motor fluctuations that typically emerge after approximately 4 years of levodopa therapy.<br>NEJM<br>[1]<br>Her current levodopa dosing interval of every 3 hours reflects attempts to manage wearing-off effects.<br><br>Mood symptoms: Patient exhibits depression related to loss of meaningful activities and sense of identity. Depression is common in Parkinson's disease and may be exacerbated by motor fluctuations.<br>Lancet<br>+ 1<br>[2][5]<br>Current treatment with citalopram 20 mg daily represents appropriate first-line therapy with an SSRI.<br>Lancet<br>+ 2<br>[2][5-6]<br><br>Current medication assessment: The patient is on a moderate total daily levodopa dose (600 mg) with frequent dosing intervals. She is already receiving MAO-B inhibitor therapy (selegiline) as adjunctive treatment. The short dosing intervals suggest emerging motor fluctuations with end-of-dose wearing off. | current levodopa/carbidopa regimen.<br>Rationale: Opicapone is a catechol-O-methyltransferase (COMT) inhibitor that prolongs levodopa effect by blocking its peripheral metabolism, thereby reducing "off" time in patients experiencing motor fluctuations. |

| | |
|---|---|
| | NEJM<br>+ 1<br>[1][7]<br><br>Plan:<br><br>Add opicapone 50 mg once daily at bedtime as adjunctive therapy to current levodopa/carbidopa regimen.<br>NEJM<br>+ 3<br>[1-4]<br><br>Rationale: Opicapone is a catechol-O-methyltransferase (COMT) inhibitor that prolongs levodopa effect by blocking its peripheral metabolism, thereby reducing "off" time in patients experiencing motor fluctuations.<br>NEJM<br>+ 2<br>[1-2][7]<br>It is FDA-approved specifically as adjunctive treatment to levodopa/carbidopa in patients with Parkinson's disease experiencing "off" episodes.<br>FDA<br>[4]<br>Recent evidence-based reviews classify opicapone as efficacious for treating motor fluctuations in patients already on levodopa therapy.<br>Movement Disorders<br>[3]<br>Compared to entacapone (another COMT inhibitor), opicapone offers the advantage of once-daily dosing, which may improve adherence.<br>Lancet<br>+ 1<br>[2][8]<br><br>Monitoring and follow-up:<br><br>Monitor for improvement in tremor control and duration of "on" time over 4- |

| | |
|---|---|
| | 6 weeks<br><br>Assess for potential adverse effects including dyskinesia, nausea, or diarrhea (common with COMT inhibitors)<br>JAMA<br>[7]<br><br>If dyskinesia emerges, consider reducing individual levodopa/carbidopa doses while maintaining frequency<br>NEJM<br>+ 1<br>[1][7]<br><br>Continue citalopram 20 mg daily for mood symptoms; reassess depression severity at follow-up as improved motor function may positively impact mood<br>Lancet<br>+ 1<br>[2][5]<br><br>Maintain current selegiline dose as combination therapy with MAO-B inhibitors and COMT inhibitors is appropriate<br>NEJM<br>+ 1<br>[1][9]<br><br>Alternative considerations if inadequate response: If motor fluctuations persist despite opicapone addition, future options include switching to extended-release levodopa/carbidopa formulations, adding a dopamine agonist (though side effect profile may be less favorable), or considering device-aided therapies for advanced disease.<br>NEJM<br>+ 2<br>[1][3][10]<br><br>Functional goals: Aim to reduce tremor amplitude and improve balance stability to allow resumption of meaningful |

| | | |
|---|---|---|
| | activities, which should secondarily benefit mood and quality of life. | |
| 30 | This assessment and plan will address nocturnal hypokinesia (difficulty turning in bed) in a patient with advanced Parkinson's disease. The evidence supports optimizing nighttime dopaminergic therapy as the primary intervention, with options including extended-release formulations, dopamine agonists, and COMT inhibitors. The treatment recommendation will be based on current literature demonstrating efficacy for nocturnal motor symptoms.<br>Current Neurology and Neuroscience Reports<br>+ 2<br>[1-3]<br><br>Assessment and Plan<br><br>Assessment:<br><br>74-year-old male with 10-year history of idiopathic Parkinson's disease presenting with severe nocturnal hypokinesia manifesting as inability to turn over in bed without fully waking or sitting upright. This symptom is consistent with inadequate nocturnal dopaminergic coverage despite current regimen of levodopa/carbidopa 100/25 mg two tablets four times daily and controlled-release levodopa/carbidopa 200/50 mg at bedtime.<br><br>Nocturnal hypokinesia affects up to 70% of patients with mid-to-advanced Parkinson's disease and significantly impairs sleep quality and quality of life. The patient's current symptoms suggest that the bedtime controlled-release formulation is providing insufficient dopaminergic stimulation during the night.<br>Parkinsonism & Related Disorders | A: Nocturnal hypokinesia affects up to 70% of patients with mid-to-advanced Parkinson's disease and significantly impairs sleep quality and quality of life. The patient's current symptoms suggest that the bedtime controlled-release formulation is providing insufficient dopaminergic stimulation during the night.<br>P: Primary Treatment Recommendation: Add rotigotine transdermal patch 4 mg/24 hours at bedtime, titrating up by 2 mg weekly to a target dose of 8-10 mg/24 hours as tolerated. Rotigotine provides continuous dopaminergic stimulation throughout the night and has demonstrated significant improvement in nocturnal hypokinesia in randomized controlled trials, with objective improvements in number of turns in bed and degree of axial rotation, as well as improvements in validated sleep scales. Extended-release dopamine agonists like rotigotine control nighttime motor symptoms and improve sleep quality based on multiple assessment modalities. This approach is recommended for moderate to advanced Parkinson's disease with nocturnal symptoms. |

| | |
|---|---|
| | [2]<br>His concurrent use of melatonin 10 mg and escitalopram 10 mg indicates pre-existing sleep concerns, though escitalopram may potentially worsen REM sleep behavior disorder if present.<br>Lancet<br>[4]<br><br>Plan:<br><br>Primary Treatment Recommendation: Add rotigotine transdermal patch 4 mg/24 hours at bedtime, titrating up by 2 mg weekly to a target dose of 8-10 mg/24 hours as tolerated.<br><br>Rotigotine provides continuous dopaminergic stimulation throughout the night and has demonstrated significant improvement in nocturnal hypokinesia in randomized controlled trials, with objective improvements in number of turns in bed and degree of axial rotation, as well as improvements in validated sleep scales. Extended-release dopamine agonists like rotigotine control nighttime motor symptoms and improve sleep quality based on multiple assessment modalities. This approach is recommended for moderate to advanced Parkinson's disease with nocturnal symptoms.<br>Current Neurology and Neuroscience Reports<br>+ 2<br>[1][3][5]<br><br>Rationale: The transdermal delivery system ensures stable overnight dopaminergic stimulation without requiring the patient to wake for additional dosing. Studies show that continuous dopaminergic stimulation during the night improves nocturnal mobility, reduces motor complications, and results in better control of sleep |

| | |
|---|---|
| | quality. Rotigotine has been specifically validated using both objective wearable sensors and clinical rating scales for this indication.<br>Lancet<br>+ 1<br>[3][5]<br><br>Monitoring and Follow-up:<br><br>Assess for improvement in nocturnal turning ability and sleep quality at 4-6 weeks<br><br>Monitor for potential adverse effects including excessive daytime sleepiness, orthostatic hypotension, and impulse control disorders<br>Lancet<br>[3]<br><br>Consider alternative options if inadequate response, including opicapone (COMT inhibitor) 50 mg once daily as adjunct to levodopa, or nighttime-only subcutaneous apomorphine infusion for refractory cases<br>Current Neurology and Neuroscience Reports<br>+ 2<br>[1][6-7]<br><br>Additional Considerations:<br><br>Continue current levodopa/carbidopa regimen<br><br>Maintain melatonin for sleep onset support<br><br>Review escitalopram necessity given potential to worsen REM sleep behavior disorder if present<br>Lancet<br>[4] |

| | | |
|---|---|---|
| | Implement sleep hygiene measures and bedroom safety adaptations<br>Parkinsonism & Related Disorders<br>[2] | |
| 31 | This assessment and plan addresses tremor-predominant Parkinson's disease with suboptimal tremor control on current levodopa therapy. The medical literature indicates that tremor can be less responsive to levodopa than other motor symptoms, and several adjunctive options exist including propranolol, which has demonstrated efficacy for parkinsonian tremor.<br>NEJM<br>+ 2<br>[1-3]<br><br>Assessment and Plan<br><br>Assessment:<br><br>59-year-old female with idiopathic Parkinson's disease (3-year duration) presenting with persistent resting tremor of the left hand despite current levodopa/carbidopa therapy. She is currently taking levodopa/carbidopa 100/25 mg, 1.5 tablets three times daily (total daily levodopa dose: 450 mg). The tremor partially improves with medication but never fully resolves, worsens with stress/anxiety, and causes significant social embarrassment.<br><br>This presentation is consistent with tremor-predominant Parkinson's disease. Tremor may be less responsive to levodopa than bradykinesia and rigidity in some patients, and there is evidence that distinct phenotypes of dopamine-responsive and dopamine-resistant resting tremor exist in Parkinson's disease.<br>NEJM<br>+ 1<br>[1][4] | A: This presentation is consistent with tremor-predominant Parkinson's disease. Tremor may be less responsive to levodopa than bradykinesia and rigidity in some patients, and there is evidence that distinct phenotypes of dopamine-responsive and dopamine-resistant resting tremor exist in Parkinson's disease. The patient's partial response suggests she may have an intermediate or dopamine-resistant tremor phenotype.<br>P: Add propranolol 10 mg three times daily as adjunctive therapy for tremor control. Propranolol has been shown to reduce resting tremor amplitude by 70% and postural tremor by 50% in parkinsonian tremor and can be used as adjunctive therapy. Propranolol should be initiated at a low dose and titrated based on tremor response and tolerability. The patient should be counseled about potential side effects including fatigue, bradycardia, and hypotension. Blood pressure and heart rate should be monitored. If tremor remains inadequately controlled after 4-6 weeks of propranolol therapy, alternative adjunctive options could include increasing the levodopa dose, adding a COMT inhibitor such as opicapone 50 mg once daily, or considering other tremor-specific agents. |

| | | |
|---|---|---|
| | The patient's partial response suggests she may have an intermediate or dopamine-resistant tremor phenotype.<br><br>Plan:<br><br>Add propranolol 10 mg three times daily as adjunctive therapy for tremor control. Propranolol has been shown to reduce resting tremor amplitude by 70% and postural tremor by 50% in parkinsonian tremor and can be used as adjunctive therapy.<br>JAMA<br>[3]<br>Beta-blockers are recognized as alternative treatment options for Parkinson's disease tremor.<br>Drugs<br>+ 1<br>[2][5]<br><br>Propranolol should be initiated at a low dose and titrated based on tremor response and tolerability. The patient should be counseled about potential side effects including fatigue, bradycardia, and hypotension. Blood pressure and heart rate should be monitored. If tremor remains inadequately controlled after 4-6 weeks of propranolol therapy, alternative adjunctive options could include increasing the levodopa dose, adding a COMT inhibitor such as opicapone 50 mg once daily, or considering other tremor-specific agents.<br>Lancet<br>[6]<br><br>The patient should continue her current levodopa/carbidopa regimen while propranolol is initiated. Follow-up in 4-6 weeks to assess tremor response and medication tolerability is recommended. | |
| 32 | This patient presents with classic early morning OFF symptoms and difficulty with transfers, which are common motor | A: The patient demonstrates classic features of motor fluctuations with end-of-dose wearing off, manifesting as |

| | | |
|---|---|---|
| | fluctuations in advanced Parkinson's disease. The medical literature supports adding a COMT inhibitor to optimize levodopa therapy in this setting, and opicapone is FDA-approved specifically for adjunctive treatment in patients experiencing OFF episodes.<br>NEJM<br>+ 3<br>[1-4]<br>The National Parkinson Foundation guidelines recommend adjusting dopaminergic medication for transfer difficulties, and recent evidence supports opicapone as an effective once-daily COMT inhibitor for motor fluctuations.<br>Lancet<br>+ 2<br>[2][5-6]<br><br>Assessment and Plan<br><br>Assessment:<br><br>65-year-old male with 6-year history of idiopathic Parkinson's disease experiencing motor fluctuations with early morning OFF symptoms and difficulty with transfers. Current regimen includes levodopa/carbidopa 100/25 mg 1.5 tablets four times daily (total daily levodopa dose 600 mg), safinamide 100 mg daily, and mirabegron 50 mg daily.<br><br>The patient demonstrates classic features of motor fluctuations with end-of-dose wearing off, manifesting as significant difficulty rising from low positions (bed, car seat, soft couch). These symptoms are consistent with the shortened duration of levodopa effect that typically emerges after 4 years of treatment.<br>NEJM<br>[1]<br>The need to use compensatory strategies (rolling to side, rocking for momentum, requiring assistance) indicates inadequate | significant difficulty rising from low positions (bed, car seat, soft couch). These symptoms are consistent with the shortened duration of levodopa effect that typically emerges after 4 years of treatment. His difficulty with transfers represents a specific motor complication that warrants optimization of dopaminergic therapy.<br>P: Add opicapone 50 mg once daily at bedtime to extend levodopa duration and reduce OFF time. |

| | |
|---|---|
| | dopaminergic coverage during OFF periods, particularly in the early morning.<br><br>His difficulty with transfers represents a specific motor complication that warrants optimization of dopaminergic therapy.<br>Parkinsonism & Related Disorders<br>[5]<br>The National Parkinson Foundation guidelines recommend adjusting dopaminergic medication for transfer difficulties in the absence of orthostatic hypotension.<br>Parkinsonism & Related Disorders<br>[5]<br><br>Plan:<br><br>Add opicapone 50 mg once daily at bedtime to extend levodopa duration and reduce OFF time.<br>Lancet<br>+ 2<br>[2-4]<br><br>Opicapone is a once-daily COMT inhibitor FDA-approved as adjunctive treatment to levodopa/carbidopa in patients experiencing OFF episodes.<br>FDA<br>[4]<br>Trial data support the use of COMT inhibitors in reducing OFF time in patients with motor fluctuations, and opicapone specifically has demonstrated efficacy for early morning OFF symptoms.<br>Lancet<br>+ 2<br>[2-3][6]<br>The once-daily dosing at bedtime makes it particularly suitable for addressing morning akinesia.<br><br>The patient is already on safinamide (MAO-B inhibitor), which provides some benefit for motor fluctuations.<br>NEJM |

| | |
|---|---|
| | [1]<br>Adding opicapone provides complementary dopamine-sparing effects through a different mechanism (blocking COMT-mediated levodopa degradation).<br>Lancet<br>+ 1<br>[2][7]<br><br>Additional recommendations:<br><br>Continue current levodopa/carbidopa dosing (150 mg levodopa four times daily)<br><br>Continue safinamide 100 mg daily<br><br>Continue mirabegron 50 mg daily<br><br>Assess for orthostatic hypotension at follow-up, as this could contribute to transfer difficulties and may require dose adjustment if present<br>Parkinsonism & Related Disorders<br>[5]<br><br>Refer to physical therapy for transfer training using cues and cognitive movement strategies<br>Lancet<br>+ 2<br>[2][5][7]<br><br>Encourage regular exercise including balance training, progressive resistance training, and gait training, which have demonstrated efficacy for motor symptoms and functional outcomes in Parkinson's disease<br>Lancet<br>+ 1<br>[2][7]<br><br>Monitor for dyskinesias with the addition of opicapone, as enhanced levodopa effect may unmask or worsen involuntary movements |

| | | |
|---|---|---|
| | NEJM<br>[1]<br><br>Follow up in 4-6 weeks to assess response to opicapone and adjust therapy as needed | |
| 33 | This assessment and plan addresses a patient with advanced Parkinson's disease experiencing significant gait impairment and fall risk despite maximal medical therapy. The National Parkinson Foundation guidelines recommend optimizing dopaminergic medication and initiating physical therapy with cueing strategies for shuffling gait, stooped posture, and postural instability. Given the evidence that exercise interventions reduce falls by approximately 26% in Parkinson's disease, and that this patient is already on substantial dopaminergic therapy, the primary recommendation focuses on structured physical therapy while maintaining current medications.<br>Parkinsonism & Related Disorders<br>+ 1<br>[1-2]<br><br>Assessment and Plan<br><br>Assessment:<br><br>72-year-old female with 10-year history of idiopathic Parkinson's disease presenting with progressive gait impairment and high fall risk. Current symptoms include shuffling gait with reduced step length, stooped posture, postural instability with near-falls during turns and when bumped, and requirement for walker assistance. She is currently on substantial dopaminergic therapy: levodopa/carbidopa 100/25 mg two tablets four times daily (total daily levodopa dose 800 mg), entacapone 200 mg four times daily, and rasagiline 1 mg daily. | A: The clinical presentation is consistent with advanced Parkinson's disease with multiple fall risk factors, including shuffling gait, stooped posture, postural instability, and impaired turning. These features represent disease progression beyond what dopaminergic medication alone can adequately address. The patient is already on a robust medication regimen including levodopa, a COMT inhibitor (entacapone), and an MAO-B inhibitor (rasagiline), suggesting limited room for further medication optimization without risking adverse effects.<br>P: Initiate structured physical therapy program focusing on gait training with cueing techniques, balance exercises, and muscle strengthening. Exercise interventions have been shown to reduce fall rates by approximately 26% and decrease the number of people experiencing falls in Parkinson's disease. |

| | |
|---|---|
| | The clinical presentation is consistent with advanced Parkinson's disease with multiple fall risk factors, including shuffling gait, stooped posture, postural instability, and impaired turning. These features represent disease progression beyond what dopaminergic medication alone can adequately address. The patient is already on a robust medication regimen including levodopa, a COMT inhibitor (entacapone), and an MAO-B inhibitor (rasagiline), suggesting limited room for further medication optimization without risking adverse effects.<br>Parkinsonism & Related Disorders<br>+ 1<br>[1][3]<br><br>Plan:<br><br>Initiate structured physical therapy program focusing on gait training with cueing techniques, balance exercises, and muscle strengthening. Exercise interventions have been shown to reduce fall rates by approximately 26% and decrease the number of people experiencing falls in Parkinson's disease. The physical therapy program should include:<br>Parkinsonism & Related Disorders<br>+ 1<br>[1-2]<br><br>Gait training using visual, auditory, or tactile cues to improve step length and reduce shuffling<br>Parkinsonism & Related Disorders<br>[1]<br><br>Balance training exercises (such as Tai Chi) to address postural instability<br>Parkinsonism & Related Disorders<br>[1]<br><br>Progressive resistance and strength training to improve trunk mobility and |

| | |
|---|---|
| | posture<br>Parkinsonism & Related Disorders<br>[1]<br><br>Training in dual-tasking and turning strategies to reduce fall risk during complex movements<br>Parkinsonism & Related Disorders<br>[1]<br><br>Home safety assessment and environmental modifications (improved lighting, creating larger spaces for turning)<br>Parkinsonism & Related Disorders<br>[1]<br><br>Maintain current medication regimen of levodopa/carbidopa 100/25 mg two tablets four times daily, entacapone 200 mg four times daily, and rasagiline 1 mg daily. The patient is already on appropriate doses of levodopa with adjunctive COMT and MAO-B inhibition. While the National Parkinson Foundation guidelines suggest considering increased dopaminergic medication for shuffling gait and stooped posture, further dose escalation at this stage carries risk of dyskinesias, orthostatic hypotension, and other adverse effects without clear benefit given the advanced nature of her gait impairment.<br>Parkinsonism & Related Disorders<br>+ 2<br>[1][3-4]<br><br>Additional considerations:<br><br>Assess for freezing of gait episodes, as this is a common cause of falls in Parkinson's disease and may require specific cueing strategies<br>Parkinsonism & Related Disorders<br>[1]<br><br>Screen for orthostatic hypotension during |

| | | |
|---|---|---|
| | transfers, as this can contribute to fall risk<br>Parkinsonism & Related Disorders<br>[1]<br><br>Evaluate for cognitive impairment, particularly frontal executive dysfunction, which is associated with falls and may benefit from cholinesterase inhibitor therapy if present<br>Parkinsonism & Related Disorders<br>+ 1<br>[1][5]<br><br>Ensure walker is properly fitted and patient is trained in safe use<br><br>Consider fall diary to track circumstances and frequency of near-falls and actual falls<br><br>Follow-up: Reassess in 4-6 weeks to evaluate response to physical therapy intervention and determine if medication adjustment or additional interventions are needed. | |
| 34 | This assessment and plan will address freezing of gait in a patient with advanced Parkinson's disease who is already on optimized dopaminergic therapy. The evidence supports a dual approach: optimizing medication timing and dosing for OFF-phase freezing, combined with structured physical therapy interventions, particularly cueing strategies and gait training. Given the patient's current high-dose levodopa regimen and lack of cognitive impairment, the recommendation will focus on medication adjustment with MAO-B inhibitor addition and referral for specialized physical therapy.[1]<br><br>Assessment and Plan<br><br>Assessment:<br><br>69-year-old male with 9-year history of | A: The FOG pattern described—occurring with gait initiation and when approaching doorways—is consistent with OFF-phase freezing, which typically responds to optimization of dopaminergic medication. The patient is already on a substantial levodopa dose with COMT inhibition, suggesting the current regimen may not be providing adequate coverage throughout the dosing interval.[1] The absence of cognitive impairment or hallucinations is favorable for medication intensification and makes him an excellent candidate for physical therapy interventions.<br><br>P: Add rasagiline 1 mg once daily (or selegiline 5 mg twice daily as alternative). MAO-B inhibitors have been associated with decreased likelihood of developing FOG and can |

| | | |
|---|---|---|
| | idiopathic Parkinson's disease presenting with freezing of gait (FOG) characterized by episodes when approaching doorways and initiating walking, lasting several seconds with associated fear of falling due to forward lurching while feet remain stuck. Currently treated with levodopa/carbidopa 100/25 mg, 2 tablets four times daily every 4 hours (total daily levodopa dose 800 mg), and entacapone 200 mg four times daily. No cognitive impairment or hallucinations reported.<br><br>The FOG pattern described—occurring with gait initiation and when approaching doorways—is consistent with OFF-phase freezing, which typically responds to optimization of dopaminergic medication. The patient is already on a substantial levodopa dose with COMT inhibition, suggesting the current regimen may not be providing adequate coverage throughout the dosing interval.[1] The absence of cognitive impairment or hallucinations is favorable for medication intensification and makes him an excellent candidate for physical therapy interventions.<br><br>Plan:<br><br>Pharmacological Management:<br><br>Add rasagiline 1 mg once daily (or selegiline 5 mg twice daily as alternative). MAO-B inhibitors have been associated with decreased likelihood of developing FOG and can improve OFF-phase freezing when added to levodopa therapy. This addition may extend levodopa duration of action and reduce OFF periods when freezing occurs.[1]<br><br>Continue current levodopa/carbidopa 100/25 mg, 2 tablets four times daily and entacapone 200 mg four times daily. | improve OFF-phase freezing when added to levodopa therapy. This addition may extend levodopa duration of action and reduce OFF periods when freezing occurs.[1] + Physical Therapy and Occupational Therapy Referral. |

| | |
|---|---|
| | Monitor for dyskinesia or other adverse effects with medication adjustment.<br><br>Physical Therapy Referral:<br><br>Refer to physical therapy specializing in Parkinson's disease for gait training with cueing strategies. Visual and auditory cueing techniques have Level A evidence for reducing FOG severity and improving gait. Specific interventions should include:[2-4]<br><br>Auditory cues (metronome, music)<br><br>Visual cues (laser pen, floor markers)<br><br>Cognitive movement strategies for gait initiation and doorway navigation<br><br>Consider treadmill training, which has demonstrated significant improvement in FOG severity.[2-3]<br><br>Obstacle training may provide additional benefit given the doorway-triggered nature of his freezing.[2]<br><br>Occupational Therapy:<br>Refer for home safety evaluation and environmental modifications, including optimizing lighting and creating larger spaces for turning near doorways.[1]<br>Fall Risk Management:<br><br>Educate patient on fall prevention strategies specific to FOG episodes.<br><br>Consider assistive devices if freezing episodes worsen despite interventions.<br><br>Follow-up:<br><br>Reassess in 4-6 weeks to evaluate response to MAO-B inhibitor and physical therapy interventions. |

| | | |
|---|---|---|
| | Monitor for medication-related adverse effects, particularly dyskinesia or orthostatic hypotension.<br><br>Assess FOG severity using validated questionnaire (FOG-Q) at follow-up visits.<br><br>If inadequate response, consider advanced therapies such as deep brain stimulation or levodopa infusion therapy in consultation with movement disorders specialist.[5] | |
| 35 | This patient presents with peak-dose dyskinesia, characterized by involuntary movements occurring 45–60 minutes after levodopa dosing—the timing when levodopa levels are highest. The medical literature supports amantadine as the only FDA-approved pharmacological treatment with proven efficacy for levodopa-induced dyskinesia, with extended-release formulations offering improved pharmacokinetics and once-daily dosing.<br>Drugs<br>+ 5<br>[1-6]<br><br>Assessment and Plan<br><br>Assessment:<br><br>67-year-old male with 6-year history of idiopathic Parkinson's disease presenting with peak-dose dyskinesia. The patient experiences irregular, involuntary movements of the limbs and trunk occurring 45–60 minutes after each levodopa/carbidopa dose, more prominent in the afternoon. These movements are more noticeable to his wife than to himself, which is typical of mild dyskinesia. The timing—occurring at the expected peak plasma concentration of levodopa—is consistent with peak-dose dyskinesia, the most common form of levodopa-induced dyskinesia. | A: 67-year-old male with 6-year history of idiopathic Parkinson's disease presenting with peak-dose dyskinesia. The patient experiences irregular, involuntary movements of the limbs and trunk occurring 45–60 minutes after each levodopa/carbidopa dose, more prominent in the afternoon. These movements are more noticeable to his wife than to himself, which is typical of mild dyskinesia. The timing—occurring at the expected peak plasma concentration of levodopa—is consistent with peak-dose dyskinesia, the most common form of levodopa-induced dyskinesia.<br>P: Add amantadine extended-release 274 mg orally once daily at bedtime to reduce dyskinesia. Amantadine is the only medication with proven efficacy for treating levodopa-induced dyskinesia and is FDA-approved for this indication. The extended-release formulation provides higher and more continuous plasma levels compared to immediate-release formulations, allowing once-daily evening dosing while improving both dyskinesia and off-time. In pivotal trials, amantadine ER demonstrated significant reduction in dyskinesia severity.<br><br>Alternative strategies—such as reducing individual levodopa doses or fractionating the dosing schedule— |

| | | |
|---|---|---|
| | Drugs<br>+ 1<br>[1-2]<br><br>Current medications include levodopa/carbidopa 100/25 mg, 2 tablets four times daily (total daily levodopa dose: 800 mg), and safinamide 100 mg once daily. The patient remains functionally independent, suggesting the dyskinesia is not yet severely disabling.<br><br>Plan:<br><br>Add amantadine extended-release 274 mg orally once daily at bedtime to reduce dyskinesia.<br>Drugs<br>+ 1<br>[5-6]<br><br>Amantadine is the only medication with proven efficacy for treating levodopa-induced dyskinesia and is FDA-approved for this indication. The extended-release formulation provides higher and more continuous plasma levels compared to immediate-release formulations, allowing once-daily evening dosing while improving both dyskinesia and off-time. In pivotal trials, amantadine ER demonstrated significant reduction in dyskinesia severity.<br>Lancet<br>+ 3<br>[3-6]<br><br>Alternative strategies—such as reducing individual levodopa doses or fractionating the dosing schedule—could theoretically reduce peak-dose dyskinesia, but would likely worsen motor control and increase off-time given the patient's current four-times-daily dosing regimen.<br>Drugs<br>[1]<br>Since the patient remains functionally | could theoretically reduce peak-dose dyskinesia, but would likely worsen motor control and increase off-time given the patient's current four-times-daily dosing regimen. Since the patient remains functionally independent and the dyskinesia is mild (more noticeable to family than patient), adding an antidyskinetic agent is preferable to reducing dopaminergic therapy.<br><br>The patient should be monitored for potential amantadine side effects including visual hallucinations, peripheral edema, and livedo reticularis. If dyskinesia persists or worsens despite amantadine therapy, consider referral to a movement disorders specialist for evaluation of advanced therapies including deep brain stimulation. |

| | | |
|---|---|---|
| | independent and the dyskinesia is mild (more noticeable to family than patient), adding an antidyskinetic agent is preferable to reducing dopaminergic therapy.<br>NEJM<br>[7]<br><br>The patient should be monitored for potential amantadine side effects including visual hallucinations, peripheral edema, and livedo reticularis. If dyskinesia persists or worsens despite amantadine therapy, consider referral to a movement disorders specialist for evaluation of advanced therapies including deep brain stimulation.<br>JAMA<br>[6] | |
| 36 | The following assessment and plan addresses orthostatic hypotension in a patient with Parkinson's disease, incorporating both non-pharmacological and pharmacological management strategies. The treatment recommendation prioritizes midodrine as first-line therapy based on evidence from multiple guidelines and clinical trials demonstrating its efficacy in neurogenic orthostatic hypotension.<br>ACC<br>+ 2<br><br>Assessment and Plan<br><br>Assessment:<br><br>72-year-old female with 9-year history of idiopathic Parkinson's disease presenting with symptomatic neurogenic orthostatic hypotension. Current antiparkinsonian regimen includes levodopa-carbidopa controlled release 200/50 mg at bedtime, levodopa-carbidopa 100/25 mg 1.5 tablets four times daily every 4 hours, and rasagiline 1 mg once daily. She reports frequent lightheadedness and | A: The orthostatic hypotension is likely neurogenic in etiology, secondary to autonomic dysfunction associated with Parkinson's disease. Morning symptoms may be exacerbated by overnight dosing of controlled-release levodopa-carbidopa and nocturnal supine hypertension, which is commonly associated with neurogenic orthostatic hypotension.<br>Lancet<br><br>P: Non-pharmacological measures: Educate patient to rise slowly from supine or seated positions, particularly in the morning, Elevate head of bed 30-45 degrees to reduce nocturnal supine hypertension and improve morning orthostatic tolerance, Increase fluid intake to 2-2.5 liters daily, Increase dietary salt intake (unless contraindicated by other medical conditions), Consider waist-high compression stockings (20-30 mmHg) if tolerated, Avoid large meals and minimize alcohol consumption, Review and discontinue any medications that may worsen orthostatic hypotension |

| | | |
|---|---|---|
| | near-fainting episodes upon standing in the morning and after prolonged sitting, without associated chest pain or palpitations. Orthostatic vital signs demonstrate a significant blood pressure drop from 130/80 mmHg supine to 95/60 mmHg standing, meeting diagnostic criteria for orthostatic hypotension (≥20 mmHg systolic or ≥10 mmHg diastolic drop within 3 minutes of standing).<br><br>The orthostatic hypotension is likely neurogenic in etiology, secondary to autonomic dysfunction associated with Parkinson's disease.<br>Lancet<br>Morning symptoms may be exacerbated by overnight dosing of controlled-release levodopa-carbidopa and nocturnal supine hypertension, which is commonly associated with neurogenic orthostatic hypotension.<br>Lancet<br><br>Plan:<br><br>Non-pharmacological measures:<br><br>Educate patient to rise slowly from supine or seated positions, particularly in the morning<br><br>Elevate head of bed 30-45 degrees to reduce nocturnal supine hypertension and improve morning orthostatic tolerance<br>Lancet<br><br>Increase fluid intake to 2-2.5 liters daily<br><br>Increase dietary salt intake (unless contraindicated by other medical conditions)<br><br>Consider waist-high compression stockings (20-30 mmHg) if tolerated<br>Lancet | <br>Pharmacological treatment:<br>Initiate midodrine 2.5 mg three times daily (morning, midday, and early afternoon), with the last dose at least 3-4 hours before bedtime to minimize supine hypertension.<br>Midodrine is an alpha-1 adrenergic agonist that increases peripheral vascular resistance through vasoconstriction and is FDA-approved for orthostatic hypotension.<br>It has demonstrated significant improvement in standing systolic blood pressure and symptom scores in randomized controlled trials.<br>Titrate midodrine by 2.5 mg increments every 3-7 days as needed for symptom control, up to a maximum of 10 mg three times daily<br>Monitor for adverse effects including piloerection, scalp itching, urinary retention, and supine hypertension<br>Measure supine and standing blood pressures at follow-up visits. Avoid dosing within 3-5 hours of bedtime to prevent nocturnal supine hypertension |

| | |
|---|---|
| | + 1<br><br>Avoid large meals and minimize alcohol consumption<br><br>Review and discontinue any medications that may worsen orthostatic hypotension<br><br>Pharmacological treatment:<br><br>Initiate midodrine 2.5 mg three times daily (morning, midday, and early afternoon), with the last dose at least 3-4 hours before bedtime to minimize supine hypertension.<br>ACC<br>+ 2<br>Midodrine is an alpha-1 adrenergic agonist that increases peripheral vascular resistance through vasoconstriction and is FDA-approved for orthostatic hypotension.<br>AAFP<br>It has demonstrated significant improvement in standing systolic blood pressure and symptom scores in randomized controlled trials.<br>AAFP<br><br>Titrate midodrine by 2.5 mg increments every 3-7 days as needed for symptom control, up to a maximum of 10 mg three times daily<br>AAFP<br><br>Monitor for adverse effects including piloerection, scalp itching, urinary retention, and supine hypertension<br>AAFP<br><br>Measure supine and standing blood pressures at follow-up visits<br><br>Avoid dosing within 3-5 hours of bedtime to prevent nocturnal supine hypertension<br>AAFP<br>+ 1 |

| | | |
|---|---|---|
| | Alternative considerations if midodrine is ineffective or not tolerated:<br><br>Droxidopa 100 mg three times daily, titrated up to 600 mg three times daily as needed (note: carbidopa may decrease effectiveness of droxidopa)<br>ACC<br>+ 1<br><br>Fludrocortisone 0.1-0.2 mg once daily (use with caution due to risk of supine hypertension, edema, and hypokalemia; doses >0.2 mg/day not recommended)<br>Lancet<br>+ 2<br><br>Follow-up:<br><br>Reassess symptoms and orthostatic vital signs in 2-4 weeks<br><br>Monitor for development or worsening of supine hypertension<br><br>If symptoms persist despite midodrine optimization, consider referral to autonomic disorders specialist or combination therapy<br>Lancet<br><br>Continue current Parkinson's disease medications without adjustment at this time, as optimizing orthostatic hypotension management takes priority | |
| 37 | The assessment and plan will address this patient's benign visual hallucinations in the context of Parkinson's disease psychosis. The evidence supports a stepwise approach, with acetylcholinesterase inhibitors as first-line pharmacological treatment for well-formed visual hallucinations in Parkinson's disease, particularly given that these hallucinations are currently non-distressing and the patient maintains | A: These visual hallucinations are consistent with Parkinson's disease-related psychosis, which commonly manifests as well-formed visual hallucinations and is associated with cholinergic, serotonergic, and dopaminergic dysfunction. The patient's high total daily levodopa dose (1200 mg) may be contributing to hallucination development. Importantly, the patient's preserved |

| | | |
|---|---|---|
| | insight.<br>Expert Review of Neurotherapeutics<br><br>Assessment and Plan<br><br>Assessment:<br><br>70-year-old male with idiopathic Parkinson's disease (8-year duration) presenting with benign visual hallucinations consistent with Parkinson's disease psychosis. The patient experiences well-formed visual hallucinations (small animals, children running) occurring in the evenings while watching television. Critically, he maintains full insight into the unreality of these perceptions and reports they are non-distressing.<br><br>Current medications include:<br><br>Levodopa/benserazide (Prolopa) 200 mg six times daily (total daily levodopa dose: 1200 mg)<br><br>Safinamide 100 mg once daily<br><br>Mirabegron 50 mg once daily<br><br>Citalopram 20 mg once daily<br><br>The hallucinations are characteristic of Parkinson's disease psychosis, which commonly manifests as well-formed visual hallucinations and involves cholinergic, serotonergic (especially 5-HT2A), and dopaminergic dysfunction.<br>Lancet<br>While these symptoms do not always require treatment when they are non-distressing and insight is preserved, the presence of visual hallucinations warrants intervention given their association with greater cognitive | insight and lack of distress indicate these hallucinations do not currently require aggressive intervention.<br>Plan: Given that the hallucinations are non-distressing and insight is preserved, active pharmacological intervention is not immediately indicated. However, the following approach is recommended:<br>Patient and caregiver education: Reassure that visual hallucinations are common in Parkinson's disease and do not necessarily indicate dementia or require treatment when non-distressing.<br>Neurology, Neurosurgery, and Psychiatry. Monitor for progression: Assess at follow-up visits whether hallucinations become more frequent, distressing, or associated with loss of insight, paranoid delusions, or cognitive decline.<br><br>Rule out medical triggers: Ensure no intercurrent infection, constipation, metabolic derangement, or medication changes that could acutely worsen psychosis.<br><br>Committed treatment recommendation if symptoms worsen: Should hallucinations become distressing or problematic, initiate rivastigmine 1.5 mg twice daily, titrating up to 3-6 mg twice daily as tolerated. Rivastigmine has demonstrated efficacy for visual hallucinations in Parkinson's disease dementia and may have preventive effects. This approach is preferred over antipsychotics given the patient's lack of dementia and the specific efficacy of cholinesterase inhibitors for visual hallucinations even in the absence of significant cognitive impairment. |
| 38 | The assessment and plan will address diarrhea as a known adverse effect of | A: Diarrhea is a well-recognized adverse effect of entacapone, the |

| | | |
|---|---|---|
| | entacapone (the COMT inhibitor component of Stalevo), which is well-documented in the literature.<br>FDA<br>+ 1<br>The treatment recommendation will be to discontinue Stalevo and transition to carbidopa-levodopa alone, as the diarrhea is likely medication-related and discontinuation of entacapone-containing products typically leads to resolution.<br>FDA<br><br>Assessment and Plan<br><br>Assessment:<br><br>68-year-old female with idiopathic Parkinson's disease (1 year duration) presenting with persistent loose stools and mild abdominal cramps for several weeks, temporally associated with initiation of Stalevo 100 mg three times daily. No alarm features (blood, mucus, fever, or weight loss).<br><br>Differential Diagnosis:<br><br>Medication-induced diarrhea (most likely) - Entacapone, the COMT inhibitor component of Stalevo, is a well-recognized cause of diarrhea<br>FDA<br>+ 1<br><br>Parkinson's disease-related gastrointestinal dysfunction<br><br>Infectious or inflammatory colitis (less likely given absence of alarm features)<br><br>Clinical Reasoning:<br><br>Diarrhea is one of the more prominent non-dopaminergic adverse events associated with levodopa-carbidopa-entacapone (Stalevo). | COMT inhibitor component of Stalevo, and can have delayed onset. This adverse effect is attributable to the entacapone component rather than levodopa or carbidopa. While diarrhea may occasionally be associated with colitis in patients taking entacapone, the absence of blood, mucus, or systemic symptoms makes this less likely.<br><br>Plan: Discontinue Stalevo and transition to carbidopa/levodopa 25/100 mg three times daily (matching the levodopa dose currently received). Diarrhea associated with entacapone is expected to resolve after discontinuation of the drug. |

| | | |
|---|---|---|
| | European Journal of Neurology<br>The temporal relationship between Stalevo initiation and symptom onset strongly suggests medication-induced etiology. Prolonged diarrhea associated with carbidopa-levodopa-entacapone is expected to resolve upon discontinuation of the drug.<br>FDA<br>While Parkinson's disease itself commonly causes gastrointestinal symptoms, constipation is far more typical than diarrhea.<br>Lancet<br>+ 2<br><br>The absence of blood, mucus, fever, or systemic symptoms makes infectious or inflammatory causes less likely, though colitis has been reported in rare cases with | |
| 39 | This assessment and plan addresses a tyramine-induced hypertensive reaction in a patient on rasagiline. The evidence indicates that while rasagiline is selective for MAO-B at recommended doses and dietary tyramine restriction is not ordinarily required, certain foods containing very high amounts of tyramine (≥150 mg) can cause hypertensive reactions in individual patients due to increased sensitivity. The recommended approach is to reduce the rasagiline dose when used as adjunct to levodopa and reinforce dietary education.<br>FDA<br>+ 1<br><br>Assessment and Plan<br><br>Assessment:<br><br>65-year-old male with idiopathic Parkinson's disease (5-year duration) presenting with a tyramine-induced hypertensive reaction following ingestion of aged cheese while on rasagiline 1 mg | A: The patient's clinical presentation is consistent with postmarketing reports of significantly elevated blood pressure after ingestion of tyramine-rich foods while taking recommended doses of rasagiline.<br><br>P: Reduce rasagiline to 0.5 mg once daily. When used as adjunct to levodopa, the recommended starting dose is 0.5 mg once daily, which may be increased to 1 mg as needed.<br><br>The lower dose maintains adequate MAO-B inhibition while reducing tyramine sensitivity and the risk of hypertensive reactions.<br><br>Counsel the patient to avoid foods containing very large amounts of tyramine (particularly aged cheeses) while taking rasagiline.<br>Advise the patient to contact their healthcare provider if they experience elevation of blood pressure or feel unwell after eating tyramine-rich foods. |

| | | |
|---|---|---|
| | daily as adjunct to levodopa/carbidopa therapy.<br><br>The patient experienced headache, flushing, and hypertension (190/100 mmHg) consistent with a tyramine reaction. While rasagiline is selective for MAO-B at recommended doses and dietary tyramine restriction is not ordinarily required, certain foods containing very high amounts of tyramine (≥150 mg, such as aged cheeses) can cause hypertensive reactions in individual patients due to increased sensitivity to tyramine. Postmarketing reports have documented rare cases of hypertensive crisis<br>FDA<br>+ 1 | Monitor blood pressure at follow-up visits, as rasagiline may be associated with elevations in blood pressure requiring medication adjustment. |
| 40 | The assessment and plan will address the likely selegiline-induced insomnia and increased daytime fatigue, with a specific treatment recommendation grounded in the available evidence. The medical literature supports that MAO-B inhibitors like selegiline can contribute to sleep disturbances, and several pharmacological options have demonstrated efficacy for insomnia in Parkinson's disease patients.<br>Lancet<br>+ 3<br><br>Assessment and Plan<br><br>Assessment:<br><br>71-year-old female with idiopathic Parkinson's disease (7-year duration) presenting with new-onset insomnia and increased daytime fatigue temporally associated with selegiline initiation.<br>Current regimen includes levodopa/carbidopa 100/25 mg, 1.5 tablets four times daily every 4 hours, and selegiline 5 mg twice daily. | A:<br>71-year-old female with idiopathic Parkinson's disease (7-year duration) presenting with new-onset insomnia and increased daytime fatigue temporally associated with selegiline initiation.<br>The insomnia is characterized by difficulty with sleep onset and maintenance without vivid dreams or anxiety, which is consistent with medication-induced sleep disruption rather than REM sleep behavior disorder. Selegiline, as an MAO-B inhibitor with amphetamine metabolites, is a recognized contributor to sleep disturbances in Parkinson's disease.<br>The concurrent increase in daytime fatigue likely reflects poor nocturnal sleep quality, as sleep fragmentation commonly leads to excessive daytime sleepiness in this population.<br>While MAO-B inhibitors may have some benefit for fatigue in Parkinson's disease, the temporal relationship between selegiline initiation and symptom onset strongly suggests the |

| | | |
|---|---|---|
| | The insomnia is characterized by difficulty with sleep onset and maintenance without vivid dreams or anxiety, which is consistent with medication-induced sleep disruption rather than REM sleep behavior disorder. Selegiline, as an MAO-B inhibitor with amphetamine metabolites, is a recognized contributor to sleep disturbances in Parkinson's disease.<br>Lancet<br>The concurrent increase in daytime fatigue likely reflects poor nocturnal sleep quality, as sleep fragmentation commonly leads to excessive daytime sleepiness in this population.<br>Lancet<br>+ 1<br><br>While MAO-B inhibitors may have some benefit for fatigue in Parkinson's disease, the temporal relationship between selegiline initiation and symptom onset strongly suggests the medication is causing more harm than benefit in this patient.<br>Clinical Medicine<br>Insomnia in Parkinson's disease is multifactorial and requires assessment of contributing medications as a first-line management strategy.<br>Lancet<br><br>Plan:<br><br>Discontinue selegiline 5 mg twice daily. The temporal association with insomnia onset and the known sleep-disrupting effects of this medication make it the primary culprit. Discontinuation should improve both nocturnal sleep and daytime fatigue.<br><br>For insomnia management if symptoms persist after selegiline discontinuation, initiate melatonin 3 mg orally at bedtime. Melatonin has demonstrated efficacy and | medication is causing more harm than benefit in this patient. Insomnia in Parkinson's disease is multifactorial and requires assessment of contributing medications as a first-line management strategy.<br>Plan:<br>Discontinue selegiline 5 mg twice daily. The temporal association with insomnia onset and the known sleep-disrupting effects of this medication make it the primary culprit. Discontinuation should improve both nocturnal sleep and daytime fatigue.<br>For insomnia management if symptoms persist after selegiline discontinuation, initiate melatonin 3 mg orally at bedtime. Melatonin has demonstrated efficacy and excellent tolerability for sleep quality improvement in Parkinson's disease patients, with no reported adverse events in controlled trials. |

| | |
|---|---|
| | excellent tolerability for sleep quality improvement in Parkinson's disease patients, with no reported adverse events in controlled trials.<br>Neurological Sciences<br>This represents a safer first-line option compared to benzodiazepines or other sedative-hypnotics, which should be avoided in older adults per the American Geriatrics Society Beers Criteria.<br>American Geriatrics Society<br><br>Alternative pharmacological options if melatonin proves insufficient include low-dose doxepin (up to 6 mg at bedtime) or dual orexin receptor antagonists (suvorexant, lemborexant, or daridorexant), all for short-term use.<br>American Geriatrics Society<br>+ 1<br>Trazodone 50 mg at bedtime is another option that has shown efficacy for sleep quality in Parkinson's disease, though it demonstrated higher rates of mild adverse events compared to melatonin.<br>Neurological Sciences<br><br>Counsel on sleep hygiene measures including maintaining a consistent sleep schedule, optimizing the sleep environment, and avoiding caffeine in the afternoon and evening.<br>Lancet<br>+ 1<br><br>Reassess in 2-4 weeks after selegiline discontinuation to evaluate improvement in sleep quality and daytime fatigue. If fatigue persists despite improved sleep, consider alternative etiologies and potential treatments such as rasagiline (which has evidence for reducing physical fatigue without the sleep-disrupting effects of selegiline) or optimization of levodopa dosing.<br>JAMA<br>+ 1 |

| | | |
|---|---|---|
| 41 | This assessment and plan will address Parkinson's disease psychosis with visual hallucinations in a patient with preserved insight. The evidence supports pimavanserin as first-line therapy for mild, non-distressing psychotic symptoms in Parkinson's disease, which is the only FDA-approved medication for this indication.<br>Expert Opinion on Drug Safety<br>+ 5<br><br>Assessment and Plan<br><br>Assessment:<br><br>63-year-old male with 7-year history of idiopathic Parkinson's disease presenting with new-onset visual hallucinations (seeing animals quickly passing in front of him). The patient demonstrates preserved insight (recognizes hallucinations are not real) and is not distressed by these symptoms. Current dopaminergic regimen includes levodopa/carbidopa 100/25 mg (2 tablets six times daily), Sinemet CR 200/50 mg at bedtime, and safinamide 100 mg daily.<br><br>This presentation is consistent with Parkinson's disease psychosis (PDP), which occurs in 18-50% of patients with Parkinson's disease.<br>Current Pharmaceutical Design<br>Visual hallucinations are the most common psychotic manifestation in PDP.<br>Lancet<br>+ 1<br>The patient's preserved insight and lack of distress indicate mild psychotic symptoms that are appropriate for pharmacologic intervention.<br>Expert Opinion on Drug Safety<br>+ 1<br><br>Plan: | A:This presentation is consistent with Parkinson's disease psychosis (PDP), which occurs in 18-50% of patients with Parkinson's disease.<br>P: Rule out precipitating factors: Evaluate for delirium triggers including intercurrent infection, constipation, metabolic disturbances, or new medications, as these are the most frequent causes of acute or worsening psychosis symptoms.<br>Medication review: The current dopaminergic regimen is relatively high-dose. However, given the patient's lack of distress and preserved insight, immediate reduction of anti-parkinsonian medications is not necessary at this time.<br>Initiate pimavanserin: Start pimavanserin 34 mg once daily. Pimavanserin is the only FDA-approved medication for hallucinations and delusions associated with Parkinson's disease psychosis and is recommended as first-line treatment for mild psychotic symptoms that do not require rapid response. As a selective 5-HT2A inverse agonist, pimavanserin does not worsen motor function and has a favorable tolerability profile compared to traditional antipsychotics.<br><br>Alternative options if pimavanserin fails or is not tolerated:<br><br>Quetiapine (starting at 12.5-25 mg at bedtime) for symptoms requiring more rapid improvement, though evidence for efficacy is limited<br>Expert Opinion on Drug Safety |

| | |
|---|---|
| | Rule out precipitating factors: Evaluate for delirium triggers including intercurrent infection, constipation, metabolic disturbances, or new medications, as these are the most frequent causes of acute or worsening psychosis symptoms.<br>Lancet<br>+ 1<br><br>Medication review: The current dopaminergic regimen is relatively high-dose. However, given the patient's lack of distress and preserved insight, immediate reduction of anti-parkinsonian medications is not necessary at this time.<br>JAMA<br>+ 1<br><br>Initiate pimavanserin: Start pimavanserin 34 mg once daily. Pimavanserin is the only FDA-approved medication for hallucinations and delusions associated with Parkinson's disease psychosis and is recommended as first-line treatment for mild psychotic symptoms that do not require rapid response. As a selective 5-HT2A inverse agonist, pimavanserin does not worsen motor function and has a favorable tolerability profile compared to traditional antipsychotics.<br>Expert Opinion on Drug Safety<br>+ 6<br><br>Monitor response: Reassess symptoms in 4-6 weeks. If hallucinations persist or worsen, consider adding a cholinesterase inhibitor (rivastigmine or donepezil), which can be effective for visual hallucinations even in the absence of dementia.<br>Lancet<br>+ 1<br><br>Alternative options if pimavanserin fails or is not tolerated: |

| | | |
|---|---|---|
| | Quetiapine (starting at 12.5-25 mg at bedtime) for symptoms requiring more rapid improvement, though evidence for efficacy is limited<br>Expert Opinion on Drug Safety<br>+ 2<br><br>Clozapine (12.5-50 mg daily) for refractory or severe symptoms, though this requires Risk Evaluation and Mitigation Strategy (REMS) program enrollment and regular monitoring for neutropenia<br>Expert Opinion on Pharmacotherapy<br>+ 2<br><br>Patient and caregiver education: Discuss the nature of PDP, expected treatment timeline, and importance of reporting any worsening symptoms or new delusions. | |
| 42 | The dark orange urine discoloration is a well-recognized, benign adverse effect of entacapone therapy that requires no intervention beyond patient reassurance. The assessment and plan will document this finding, confirm its benign nature with the normal urinalysis, and provide patient education to continue current therapy without modification.<br>FDA<br><br>Assessment and Plan<br><br>Assessment:<br><br>69-year-old female with idiopathic Parkinson's disease (9-year duration) on levodopa/carbidopa 100/25 mg 2.5 tablets four times daily and entacapone 200 mg four times daily, presenting with dark orange urine discoloration. Urinalysis is normal with no dysuria, frequency, or pain.<br><br>Primary Diagnosis: Benign urine discoloration secondary to entacapone therapy | A: Primary Diagnosis: Benign urine discoloration secondary to entacapone therapy<br>The patient's dark orange urine discoloration is a well-documented, harmless adverse effect of entacapone occurring in approximately 10% of patients treated with this medication. This discoloration represents a brownish-orange color change caused by entacapone metabolites and is not clinically relevant. The normal urinalysis confirms the absence of urinary tract pathology, and the lack of dysuria, frequency, or pain further supports that this is a benign medication effect rather than a urological condition.<br><br>Plan: Continue current Parkinson's disease regimen without modification:<br>Patient education: Reassure the patient that the dark orange urine discoloration is an expected, harmless side effect of entacapone therapy that does not require treatment or medication adjustment. Advise that |

| | | |
|---|---|---|
| | FDA<br><br>The patient's dark orange urine discoloration is a well-documented, harmless adverse effect of entacapone occurring in approximately 10% of patients treated with this medication. This discoloration represents a brownish-orange color change caused by entacapone metabolites and is not clinically relevant. The normal urinalysis confirms the absence of urinary tract pathology, and the lack of dysuria, frequency, or pain further supports that this is a benign medication effect rather than a urological condition.<br>FDA<br><br>Plan:<br><br>Continue current Parkinson's disease regimen without modification:<br><br>Levodopa/carbidopa 100/25 mg, 2.5 tablets four times daily every 4 hours<br><br>Entacapone 200 mg four times daily (one tablet with each levodopa/carbidopa dose)<br>FDA<br><br>Patient education: Reassure the patient that the dark orange urine discoloration is an expected, harmless side effect of entacapone therapy that does not require treatment or medication adjustment. Advise that this discoloration may also affect saliva or sweat.<br>FDA<br>+ 2<br><br>No further urological workup indicated given normal urinalysis and absence of urinary symptoms.<br><br>Routine follow-up for Parkinson's disease management as previously scheduled. | this discoloration may also affect saliva or sweat.<br>No further urological workup indicated given normal urinalysis and absence of urinary symptoms. |

| | | |
|---|---|---|
| | Rationale: Entacapone is appropriately indicated as adjunctive therapy for this patient's end-of-dose wearing-off symptoms in Parkinson's disease. The urine discoloration, while noticeable, is a benign cosmetic effect that does not warrant discontinuation of an otherwise effective medication regimen.<br>FDA<br>No dose adjustment or medication change is necessary. | |
| 43 | This assessment and plan addresses sleep attacks (sudden onset sleep without warning) in a patient on pramipexole, a well-documented adverse effect of dopamine agonists. The medical literature supports reducing or discontinuing the dopamine agonist as the primary intervention for this potentially dangerous complication.<br>Lancet<br>+ 2<br><br>Assessment and Plan<br><br>Assessment:<br><br>62-year-old male with idiopathic Parkinson's disease (3 years duration) presenting with sleep attacks (sudden onset sleep without warning during activities of daily living). Currently treated with levodopa/carbidopa 100/25 mg four times daily and pramipexole 0.75 mg three times daily (total daily dose 2.25 mg).<br><br>The patient's presentation is consistent with dopamine agonist-induced sleep attacks, a recognized adverse effect of pramipexole. Patients may fall asleep during activities without perceiving warning signs of drowsiness, and these events can occur even one year after treatment initiation. Pramipexole and other dopamine agonists have higher | A: The patient's presentation is consistent with dopamine agonist-induced sleep attacks, a recognized adverse effect of pramipexole. Patients may fall asleep during activities without perceiving warning signs of drowsiness, and these events can occur even one year after treatment initiation. Pramipexole and other dopamine agonists have higher affinity for D2 and D3 receptors compared to levodopa, making them more likely to induce excessive daytime sleepiness and sleep attacks. The patient's current pramipexole dose (2.25 mg/day) exceeds the threshold where somnolence becomes significantly more common (>1.5 mg/day).<br>This represents a safety concern as sleep attacks during activities pose risk of injury, including motor vehicle accidents.<br><br>Plan: Discontinue pramipexole with gradual taper to minimize withdrawal effects and monitor for worsening parkinsonism. Reduce by 0.25 mg every 3-5 days until discontinued. |

| | |
|---|---|
| | affinity for D2 and D3 receptors compared to levodopa, making them more likely to induce excessive daytime sleepiness and sleep attacks. The patient's current pramipexole dose (2.25 mg/day) exceeds the threshold where somnolence becomes significantly more common (>1.5 mg/day).<br>Lancet<br>+ 3<br><br>This represents a safety concern as sleep attacks during activities pose risk of injury, including motor vehicle accidents.<br>FDA<br>+ 1<br><br>Plan:<br><br>Discontinue pramipexole with gradual taper to minimize withdrawal effects and monitor for worsening parkinsonism. Reduce by 0.25 mg every 3-5 days until discontinued.<br><br>Rationale: When patients develop significant daytime sleepiness or sleep attacks during activities requiring active participation, pramipexole should ordinarily be discontinued. Case reports demonstrate that progressive reduction of dopamine agonist doses can resolve excessive daytime sleepiness without worsening parkinsonism. If motor symptoms worsen during the taper, the levodopa dose can be increased (in controlled studies, levodopa doses were reduced by an average of 27% when pramipexole was added, suggesting room for dose adjustment).<br>Lancet<br>+ 2<br><br>The current levodopa/carbidopa regimen (100/25 mg four times daily) will be maintained initially and adjusted as needed based on motor symptom control |

| | | |
|---|---|---|
| | during the pramipexole taper.<br><br>Counseling: Advise the patient to avoid driving and other potentially dangerous activities until sleep attacks resolve. Educate regarding the medication-related nature of symptoms and expected improvement with pramipexole discontinuation.<br>FDA<br><br>Follow-up: Reassess in 2-3 weeks to evaluate resolution of sleep attacks and adequacy of motor symptom control on levodopa monotherapy. | |
| 44 | This assessment and plan will address dopamine agonist-induced impulse control disorder (ICD), specifically compulsive buying, in this patient with Parkinson's disease. The treatment recommendation follows expert consensus guidelines that prioritize gradual reduction or discontinuation of dopamine agonists as first-line therapy, while monitoring for dopamine agonist withdrawal syndrome and motor symptom worsening.<br>Movement Disorders<br>+ 2<br><br>Assessment and Plan<br><br>Assessment:<br><br>70-year-old female with 6-year history of idiopathic Parkinson's disease presenting with impulse control disorder (ICD), specifically compulsive buying, manifested by impulsive purchase of a luxury vehicle using most of her savings without consulting her wife. This behavior is consistent with dopamine agonist-induced ICD, a well-recognized complication affecting approximately 10% of patients per year on dopaminergic therapy.<br>Current Opinion in Neurology | A: Dopamine agonists, particularly ropinirole, are the primary risk factor for ICDs in Parkinson's disease.<br>Plan: Primary Treatment Recommendation: Gradual tapering and discontinuation of ropinirole:<br>Reduce ropinirole by 1 mg every 1-2 weeks (e.g., decrease from 4 mg TID to 3 mg TID, then 2 mg TID, etc.) with the goal of complete discontinuation<br>Monitor closely for dopamine agonist withdrawal syndrome (DAWS), characterized by anxiety, panic attacks, dysphoria, fatigue, and drug craving<br>Monitor for motor symptom worsening during taper |

| | |
|---|---|
| | The patient is currently on:<br><br>Levodopa/carbidopa 100/25 mg, 2 tablets four times daily (total daily levodopa dose: 800 mg)<br><br>Ropinirole 4 mg three times daily (total daily dose: 12 mg)<br><br>Dopamine agonists, particularly ropinirole, are the primary risk factor for ICDs in Parkinson's disease.<br>CNS Drugs<br>+ 2<br>The patient's relatively high dose of ropinirole (12 mg daily) significantly increases this risk. ICDs can have severe psychosocial and financial consequences for patients and families, making prompt intervention essential.<br>Movement Disorders<br>+ 1<br><br>Additional risk factors present in this patient include relatively young age at onset (diagnosis at age 64) and adequate levodopa coverage, which may facilitate dopamine agonist reduction.<br>Current Opinion in Neurology<br><br>Plan:<br><br>Primary Treatment Recommendation: Gradual tapering and discontinuation of ropinirole<br><br>Reduce ropinirole by 1 mg every 1-2 weeks (e.g., decrease from 4 mg TID to 3 mg TID, then 2 mg TID, etc.) with the goal of complete discontinuation<br>Movement Disorders<br>+ 2<br><br>Monitor closely for dopamine agonist withdrawal syndrome (DAWS), characterized by anxiety, panic attacks, |

| | |
|---|---|
| | dysphoria, fatigue, and drug craving<br>Lancet<br>+ 1<br><br>Monitor for motor symptom worsening during taper<br><br>Supportive Measures:<br><br>Involve the patient's wife in monitoring and treatment planning, as caregiver involvement is essential for successful ICD management<br>Movement Disorders<br>+ 1<br><br>Screen for other ICDs (gambling, hypersexuality, binge eating) and related behaviors (punding, hobbyism)<br>Movement Disorders<br><br>Consider financial safeguards during treatment (e.g., joint account oversight, spending limits)<br><br>Schedule follow-up visits every 2-4 weeks during ropinirole taper to assess ICD symptoms, withdrawal symptoms, and motor function<br><br>If Motor Symptoms Worsen During Dopamine Agonist Taper:<br><br>May increase levodopa/carbidopa dose incrementally (e.g., add one-half to one tablet per dose) to maintain motor control<br>CNS Drugs<br>+ 1<br><br>Current total daily levodopa dose of 800 mg provides room for modest increases if needed<br><br>Second-Line Options if ICD Persists Despite Dopamine Agonist Discontinuation: |

| | |
|---|---|
| | Cognitive behavioral therapy has shown benefit in improving ICD severity<br>Lancet<br>+ 2<br><br>Consider psychiatric consultation for management of any emerging mood or anxiety symptoms<br><br>Naltrexone or other pharmacological agents have limited evidence but may be considered in refractory cases<br>Lancet<br>+ 2<br><br>Monitoring:<br><br>Regular assessment of ICD symptoms using validated screening tools at each visit<br>Movement Disorders<br>+ 1<br><br>Monitor for development of DAWS, particularly anxiety, depression, and drug craving<br><br>Assess motor symptoms (tremor, rigidity, bradykinesia) and functional status<br><br>Screen for other neuropsychiatric complications including depression, anxiety, and apathy<br><br>Patient and Caregiver Education:<br><br>Explain that ICDs are a medication side effect, not a character flaw<br><br>Discuss expected timeline for ICD improvement (typically within weeks to months of dopamine agonist reduction)<br>CNS Drugs<br>+ 1<br><br>Warn about potential for DAWS and |

| | | |
|---|---|---|
| | motor symptom worsening<br><br>Emphasize importance of adherence to tapering schedule and close follow-up | |
| 45 | The assessment and plan will address the localized application site reaction to rotigotine, which is a common adverse event occurring in 20-46% of patients in clinical trials. The evidence supports continuing dopaminergic therapy while managing the skin reaction through proper patch rotation and consideration of alternative dopamine agonist formulations if the reaction persists or worsens.<br>FDA<br>[1]<br><br>Assessment and Plan<br><br>Assessment:<br><br>61-year-old female with idiopathic Parkinson's disease (4-year duration), currently on levodopa/carbidopa 100/25 mg four times daily and rotigotine patch 4 mg/24 hours for advanced-stage disease. She presents with localized application site reaction (erythema, pruritus, mild desquamation) after two weeks of rotigotine use. No evidence of generalized skin reaction or systemic hypersensitivity.<br><br>Application site reactions are among the most common adverse events with rotigotine transdermal therapy, occurring in 36-46% of patients with advanced-stage Parkinson's disease. In clinical trials, these reactions were typically mild to moderate and transient in the majority of cases. The absence of systemic symptoms or spreading beyond the application site suggests a localized irritant reaction rather than true contact sensitization.<br>FDA<br>+ 1 | A: Application site reactions are among the most common adverse events with rotigotine transdermal therapy, occurring in 36-46% of patients with advanced-stage Parkinson's disease. In clinical trials, these reactions were typically mild to moderate and transient in the majority of cases. The absence of systemic symptoms or spreading beyond the application site suggests a localized irritant reaction rather than true contact sensitization.<br><br>Plan: Continue rotigotine patch 4 mg/24 hours with enhanced site rotation protocol. Counsel patient to rotate application sites daily and avoid repeating the same site for at least 14 days, as daily rotation has been shown to reduce the incidence of application site reactions compared to repetitive application to the same site. Apply patch to clean, dry, intact skin on the abdomen, thigh, hip, flank, shoulder, or upper arm, avoiding oily, irritated, or damaged areas.<br>Monitor the application site reaction over the next 2-4 weeks. If the reaction persists for more than a few days, increases in severity, or spreads beyond the application site, reassess the risk-benefit ratio for continued rotigotine use. The majority of patients with application site reactions recover or are recovering, and most reactions are non-serious.<br>If application site reactions become intolerable despite proper rotation, consider switching to an alternative dopamine agonist such as oral pramipexole (starting at 0.125 mg three times daily, titrating as needed) or ropinirole (starting at 0.25 mg three |

| | | |
|---|---|---|
| | [1-2]<br><br>Plan:<br><br>Continue rotigotine patch 4 mg/24 hours with enhanced site rotation protocol. Counsel patient to rotate application sites daily and avoid repeating the same site for at least 14 days, as daily rotation has been shown to reduce the incidence of application site reactions compared to repetitive application to the same site. Apply patch to clean, dry, intact skin on the abdomen, thigh, hip, flank, shoulder, or upper arm, avoiding oily, irritated, or damaged areas.<br>FDA<br>[1]<br><br>Monitor the application site reaction over the next 2-4 weeks. If the reaction persists for more than a few days, increases in severity, or spreads beyond the application site, reassess the risk-benefit ratio for continued rotigotine use. The majority of patients with application site reactions recover or are recovering, and most reactions are non-serious.<br>FDA<br>+ 1<br>[1][3]<br><br>If application site reactions become intolerable despite proper rotation, consider switching to an alternative dopamine agonist such as oral pramipexole (starting at 0.125 mg three times daily, titrating as needed) or ropinirole (starting at 0.25 mg three times daily, titrating as needed), both of which are efficacious for motor fluctuations in advanced Parkinson's disease. Alternatively, consider adding a COMT inhibitor such as opicapone 50 mg once daily at bedtime to optimize levodopa therapy.<br>Movement Disorders | times daily, titrating as needed), both of which are efficacious for motor fluctuations in advanced Parkinson's disease. Alternatively, consider adding a COMT inhibitor such as opicapone 50 mg once daily at bedtime to optimize levodopa therapy. |

| | | |
|---|---|---|
| | + 2<br>[4-6]<br><br>Continue current levodopa/carbidopa regimen at 100/25 mg four times daily. No drug-drug interaction exists between rotigotine and levodopa/carbidopa.<br>FDA<br>[1]<br><br>Follow-up in 2-4 weeks to reassess application site reaction and motor symptom control. | |
| 46 | The assessment and plan will address rivastigmine-induced gastrointestinal adverse effects, which are dose-related and common at the current 6 mg twice daily dose.<br>FDA<br>The treatment recommendation will focus on dose reduction of rivastigmine, as this is the most evidence-based approach to managing these symptoms while maintaining cognitive benefit.<br>FDA<br><br>Assessment and Plan<br><br>Assessment:<br><br>78-year-old female with idiopathic Parkinson's disease (10-year duration) presenting with daily nausea, reduced appetite, and 2 kg weight loss following recent titration to rivastigmine 6 mg twice daily. Currently managed with levodopa/carbidopa 100/25 mg four times daily (every 4 hours) and rivastigmine 6 mg twice daily for Parkinson's disease dementia.<br><br>The clinical presentation is most consistent with rivastigmine-induced gastrointestinal adverse effects. Rivastigmine commonly causes dose-related nausea, vomiting, anorexia, and weight loss, with the highest incidence | A: The clinical presentation is most consistent with rivastigmine-induced gastrointestinal adverse effects. Rivastigmine commonly causes dose-related nausea, vomiting, anorexia, and weight loss, with the highest incidence during dose titration.<br>While delayed gastric emptying is a primary feature of Parkinson's disease and can contribute to nausea, the temporal relationship between rivastigmine dose escalation and symptom onset strongly suggests medication-related etiology.<br>Levodopa absorption may be affected by gastroparesis, but the patient's current motor control on levodopa appears stable based on the dosing regimen.<br><br>P: Reduce rivastigmine dose to 4.5 mg twice daily with meals |

| | |
|---|---|
| | during dose titration.<br>FDA<br>In controlled trials, 29% of patients on rivastigmine 6-12 mg/day developed nausea (versus 11% on placebo), 17% developed anorexia (versus 3% on placebo), and 26% of women experienced ≥7% weight loss.<br>FDA<br>These gastrointestinal effects are the most common reason for treatment discontinuation.<br>FDA<br><br>While delayed gastric emptying is a primary feature of Parkinson's disease and can contribute to nausea, the temporal relationship between rivastigmine dose escalation and symptom onset strongly suggests medication-related etiology.<br>Lancet<br>+ 1<br>Levodopa absorption may be affected by gastroparesis, but the patient's current motor control on levodopa appears stable based on the dosing regimen.<br><br>Plan:<br><br>Reduce rivastigmine dose to 4.5 mg twice daily with meals<br>FDA<br><br>The rivastigmine dose should be decreased to manage gastrointestinal toxicity while maintaining cognitive benefit. Per FDA labeling, if adverse effects cause intolerance during treatment, dosing should be reduced to the same or next lower dose level.<br>FDA<br>The 4.5 mg twice daily dose represents the next lower maintenance dose and may provide adequate cognitive benefit with improved tolerability.<br>FDA |

| | | |
|---|---|---|
| | The medication should continue to be administered with meals in divided morning and evening doses.<br>FDA<br><br>If symptoms do not improve within 1-2 weeks at the reduced dose, further dose reduction to 3 mg twice daily should be considered.<br>FDA<br>If gastrointestinal symptoms resolve but cognitive decline becomes apparent, cautious re-titration may be attempted after at least 4 weeks at the lower dose.<br>FDA<br><br>Monitor weight, appetite, and nausea symptoms at 2-week follow-up<br><br>Ensure rivastigmine is taken with food to minimize gastrointestinal effects<br>FDA<br><br>Consider domperidone 10-30 mg/day if nausea persists despite dose reduction, provided QTc interval is not prolonged. Domperidone is a peripherally acting dopamine antagonist that does not worsen parkinsonian symptoms, though it requires careful cardiac monitoring.<br>Lancet<br>+ 1<br><br>Optimize levodopa timing relative to meals (administer on relatively empty stomach) to maximize absorption<br>Expert Review of Neurotherapeutics<br><br>Continue current levodopa/carbidopa regimen unless motor fluctuations develop | |
| 47 | The assessment and plan will address this patient's vivid, disturbing dreams and fragmented sleep, which are likely manifestations of REM sleep behavior disorder (RBD) in the context of advanced Parkinson's disease. The evidence | A: The clinical presentation is most consistent with REM sleep behavior disorder (RBD), a common parasomnia in Parkinson's disease characterized by dream enactment behaviors and vivid dreams. The patient's donepezil is a |

| | | |
|---|---|---|
| | supports discontinuing donepezil as the primary intervention, as acetylcholinesterase inhibitors can trigger or worsen RBD, and donepezil increases REM sleep percentage and may fragment sleep.<br>Lancet<br>+ 2<br>This recommendation is grounded in current literature on managing sleep disturbances in Parkinson's disease.<br><br>Assessment and Plan<br><br>Assessment:<br><br>79-year-old male with 11-year history of idiopathic Parkinson's disease presenting with vivid, disturbing dreams occurring almost nightly, fragmented sleep, and morning anxiety. Current medications include levodopa/carbidopa 100/25 mg, 2.5 tablets four times daily every 4 hours, and donepezil 10 mg once daily.<br><br>The clinical presentation is most consistent with REM sleep behavior disorder (RBD), a common parasomnia in Parkinson's disease characterized by dream enactment behaviors and vivid dreams. The patient's donepezil is a likely contributing factor, as acetylcholinesterase inhibitors including donepezil can trigger or worsen RBD symptoms. Additionally, donepezil significantly increases REM sleep percentage and may contribute to sleep fragmentation.<br>Lancet<br>+ 2<br><br>Levodopa at the current dosing regimen may also contribute to sleep disturbances, including nocturnal hallucinations and fragmented sleep, particularly at higher doses. However, the temporal relationship with donepezil | likely contributing factor, as acetylcholinesterase inhibitors including donepezil can trigger or worsen RBD symptoms. Additionally, donepezil significantly increases REM sleep percentage and may contribute to sleep fragmentation.<br><br>Levodopa at the current dosing regimen may also contribute to sleep disturbances, including nocturnal hallucinations and fragmented sleep, particularly at higher doses. However, the temporal relationship with donepezil initiation and the known association between cholinesterase inhibitors and RBD make donepezil the more likely primary contributor.<br><br>P: Discontinue donepezil 10 mg once daily. The risks of continuing donepezil—specifically worsening RBD symptoms and sleep fragmentation—outweigh the cognitive benefits in this clinical context. SSRIs and tricyclic antidepressants should also be avoided or reconsidered if introduced in the future, as these can similarly trigger or worsen RBD.<br><br>Reassess sleep symptoms 2-4 weeks after donepezil discontinuation. If vivid dreams and sleep fragmentation persist despite stopping donepezil, consider initiating pharmacologic treatment for RBD with either:<br><br>Melatonin 3 mg at bedtime (preferred initial option given favorable safety profile), or<br><br>Clonazepam 0.5-1 mg at bedtime (alternative if melatonin ineffective) |

| | |
|---|---|
| | initiation and the known association between cholinesterase inhibitors and RBD make donepezil the more likely primary contributor.<br>Lancet<br>+ 1<br><br>Plan:<br><br>Discontinue donepezil 10 mg once daily. The risks of continuing donepezil—specifically worsening RBD symptoms and sleep fragmentation—outweigh the cognitive benefits in this clinical context. SSRIs and tricyclic antidepressants should also be avoided or reconsidered if introduced in the future, as these can similarly trigger or worsen RBD.<br>Lancet<br>+ 1<br><br>Reassess sleep symptoms 2-4 weeks after donepezil discontinuation. If vivid dreams and sleep fragmentation persist despite stopping donepezil, consider initiating pharmacologic treatment for RBD with either:<br><br>Melatonin 3 mg at bedtime (preferred initial option given favorable safety profile), or<br>Lancet<br>+ 3<br><br>Clonazepam 0.5-1 mg at bedtime (alternative if melatonin ineffective)<br>Lancet<br>+ 2<br><br>Both melatonin and clonazepam have demonstrated efficacy for RBD in Parkinson's disease, with melatonin showing particular benefit for reducing RBD symptoms and improving sleep quality in short-term use.<br>Expert Opinion on Pharmacotherapy<br>+ 2 |

| | | |
|---|---|---|
| | Counsel patient and wife on bed safety measures to minimize risk of injury from dream enactment behaviors. Optimize sleep hygiene practices.<br>Lancet<br>+ 1<br><br>Monitor for any decline in cognitive function after donepezil discontinuation, though the immediate priority is addressing the disruptive sleep disturbance and associated RBD symptoms. | |
| 48 | The assessment and plan will address the temporal relationship between memantine dose escalation and new neuropsychiatric symptoms. Confusion is a recognized adverse effect of memantine, occurring in 6% of patients in clinical trials, and the patient's symptoms appeared after dose escalation, suggesting a likely drug-related etiology.<br>FDA<br><br>Assessment and Plan<br><br>Assessment:<br><br>80-year-old female with 12-year history of idiopathic Parkinson's disease presenting with new-onset confusion and irritability temporally associated with memantine dose escalation from 10 mg to 20 mg daily. Infectious and metabolic etiologies have been appropriately excluded.<br><br>The clinical presentation is most consistent with memantine-induced neuropsychiatric adverse effects. Confusion occurs in 6% of patients treated with memantine in controlled trials, compared to 5% with placebo.<br>FDA<br>Irritability has also been reported as an adverse reaction to memantine. | Assessment:<br>The clinical presentation is most consistent with memantine-induced neuropsychiatric adverse effects. Confusion occurs in 6% of patients treated with memantine in controlled trials, compared to 5% with placebo.<br><br>Plan: Discontinue memantine immediately. Given the lack of evidence supporting memantine use in Parkinson's disease, the temporal association with neuropsychiatric symptoms, and the known adverse effect profile, memantine should be stopped rather than dose-reduced. |

| | |
|---|---|
| | FDA<br>+ 1<br>The temporal relationship between dose escalation and symptom onset strongly supports a causal relationship.<br><br>Importantly, memantine is FDA-approved for moderate to severe Alzheimer's disease dementia, not for Parkinson's disease.<br>FDA<br>While memantine has been studied in Parkinson's disease dementia, the evidence is inconsistent and insufficient to support routine use.<br>Lancet<br>The Movement Disorders Society taskforce concluded that evidence for memantine in Parkinson's disease dementia is insufficient.<br>Lancet<br><br>Current levodopa/carbidopa dosing (250 mg levodopa four times daily, total 1000 mg/day) is appropriate for advanced Parkinson's disease.<br><br>Plan:<br><br>Discontinue memantine immediately. Given the lack of evidence supporting memantine use in Parkinson's disease, the temporal association with neuropsychiatric symptoms, and the known adverse effect profile, memantine should be stopped rather than dose-reduced.<br><br>Monitor for resolution of confusion and irritability over the next 1-2 weeks<br><br>Continue levodopa/carbidopa 100/25 mg, 2.5 tablets four times daily (current regimen)<br><br>If cognitive symptoms persist after memantine discontinuation, consider re- |

| | | |
|---|---|---|
| | evaluation for Parkinson's disease dementia and potential trial of rivastigmine, which has demonstrated efficacy in this population<br>Lancet<br><br>Follow-up in 2-4 weeks to assess symptom resolution and reassess cognitive status | |
| 49 | The bilateral leg swelling is likely related to amantadine, as peripheral edema is a known side effect that requires dose adjustment.<br>FDA<br>+ 1<br>The assessment and plan will address discontinuing amantadine and optimizing levodopa/carbidopa therapy, which remains the cornerstone of treatment for this patient with 8-year disease duration.<br>NEJM<br>+ 1<br><br>Assessment and Plan<br><br>Assessment:<br><br>69-year-old male with idiopathic Parkinson's disease for 8 years presenting with bilateral lower extremity edema. Currently treated with amantadine 100 mg twice daily and levodopa/carbidopa 100/25 mg, 2 tablets four times daily (total daily levodopa dose 800 mg). No cardiac symptoms or history to suggest heart failure. The bilateral leg swelling is most likely medication-related, specifically from amantadine, which is known to cause peripheral edema and requires dose adjustment in affected patients.<br>FDA<br>+ 1<br><br>The patient's current regimen includes levodopa/carbidopa at 4-hour intervals, which is appropriate for his disease | A: The bilateral leg swelling is most likely medication-related, specifically from amantadine, which is known to cause peripheral edema and requires dose adjustment in affected patients.<br><br>P: Discontinue amantadine 100 mg twice daily given the peripheral edema. The dose of amantadine requires careful adjustment in patients with peripheral edema. |

| | |
|---|---|
| | duration of 8 years. At this stage, motor fluctuations are common, typically emerging after approximately 4 years of levodopa therapy.<br>NEJM<br>Amantadine can be useful for managing dyskinesia and "off" time in advanced disease but should be used with caution in older populations.<br>American Geriatrics Society<br><br>Plan:<br><br>Discontinue amantadine 100 mg twice daily given the peripheral edema. The dose of amantadine requires careful adjustment in patients with peripheral edema.<br>FDA<br>+ 1<br><br>Continue levodopa/carbidopa 100/25 mg, 2 tablets four times daily (every 4 hours while awake). This provides 800 mg of levodopa daily, which is within the typical therapeutic range of 400-1600 mg per day. Levodopa remains the primary treatment for motor symptoms in Parkinson's disease.<br>NEJM<br>+ 2<br><br>Monitor for motor symptom changes after amantadine discontinuation. If the patient experiences worsening motor fluctuations or dyskinesia after stopping amantadine, consider adding a COMT inhibitor (such as entacapone or opicapone) or MAO-B inhibitor (such as rasagiline) as adjunctive therapy to levodopa.<br>NEJM<br>+ 1<br><br>Reassess lower extremity edema in 2-4 weeks. The edema should improve with amantadine discontinuation. If it persists, |

| | | |
|---|---|---|
| | further evaluation for other causes would be warranted.<br><br>Counsel patient to report any worsening of motor symptoms, development of dyskinesias, or new symptoms. Advise gradual increase in physical activity as tolerated and to avoid sudden position changes to minimize orthostatic symptoms.<br>FDA<br><br>Continue periodic monitoring for melanoma with skin examinations, as patients with Parkinson's disease have 2- to 6-fold higher risk.<br>FDA<br>+ 2 | |
| 50 | This assessment and plan will address predictable wearing-off motor fluctuations in a patient already on levodopa/carbidopa and entacapone. The evidence supports adding opicapone as the most effective next step, as it has been shown to reduce off time more effectively than increasing levodopa dose in patients with early wearing-off, and provides approximately 1 hour of additional on time when added to existing levodopa/carbidopa therapy.<br>Neurology<br>+ 3<br><br>Assessment and Plan<br><br>Assessment:<br><br>62-year-old female with idiopathic Parkinson's disease (7-year duration) experiencing predictable end-of-dose wearing-off on current regimen of levodopa/carbidopa 100/25 mg 1.5 tablets four times daily (total daily levodopa 600 mg) plus entacapone 200 mg four times daily. Wearing-off symptoms manifest 30-45 minutes before scheduled doses with both motor | A: The patient demonstrates classic motor fluctuations that typically emerge after several years of levodopa therapy due to the short half-life of levodopa, inconsistent gastrointestinal absorption, and progressive dopaminergic neuronal degeneration.<br><br>Plan: Add opicapone 50 mg once daily at bedtime (at least 1 hour before or after levodopa/carbidopa doses). |

| | |
|---|---|
| | symptoms (hand tremor, leg stiffness) and non-motor symptoms (anxiety, fatigue) that resolve with subsequent dosing. These fluctuations significantly impact quality of life and functional independence.<br><br>The patient demonstrates classic motor fluctuations that typically emerge after several years of levodopa therapy due to the short half-life of levodopa, inconsistent gastrointestinal absorption, and progressive dopaminergic neuronal degeneration.<br>NEJM<br>Current therapy with entacapone, a catechol-O-methyltransferase (COMT) inhibitor, extends levodopa half-life but is providing insufficient coverage between doses.<br><br>Plan:<br><br>Add opicapone 50 mg once daily at bedtime (at least 1 hour before or after levodopa/carbidopa doses).<br><br>Opicapone is a once-daily COMT inhibitor that is FDA-approved as adjunctive treatment to levodopa/carbidopa in patients with Parkinson's disease experiencing off episodes.<br>FDA<br>In patients with early wearing-off already on levodopa and entacapone, opicapone has been shown to reduce absolute off time by approximately 29 minutes more than adding an additional 100 mg levodopa dose.<br>Neurology<br>More broadly, COMT inhibitors including opicapone typically provide approximately 1 hour of additional on time with corresponding reduction in off time.<br>NEJM<br>+ 2 |

| | |
|---|---|
| | When initiating opicapone:<br><br>Discontinue entacapone when starting opicapone, as both are COMT inhibitors and should not be used together<br><br>Opicapone is dosed once daily (unlike entacapone which requires dosing with each levodopa dose), which may improve medication adherence<br><br>Continue current levodopa/carbidopa dosing initially (100/25 mg 1.5 tablets four times daily)<br><br>Monitor for dyskinesias as the primary potential adverse effect, as enhanced dopaminergic activity may unmask or worsen involuntary movements<br>Neurology<br>+ 1<br><br>If dyskinesias emerge, reduce individual levodopa/carbidopa doses by 12.5-25 mg levodopa (approximately 10-20% reduction) while maintaining dosing frequency<br>JAMA<br><br>Additional management considerations:<br><br>Counsel patient to separate levodopa doses from protein-rich meals by at least 1 hour, as concurrent protein intake can block levodopa absorption and contribute to dose failures<br>Lancet<br><br>Address any constipation or gastrointestinal dysmotility, as these can impair levodopa absorption and contribute to unpredictable responses<br>Lancet<br><br>Reassess in 4 weeks to evaluate reduction in off time and emergence of any adverse |

| | |
|---|---|
| | effects<br><br>Alternative approaches if opicapone is not tolerated or provides insufficient benefit include adding an MAO-B inhibitor (rasagiline or safinamide), increasing levodopa dosing frequency, or considering extended-release levodopa formulations.<br>NEJM<br>+ 2<br>However, opicapone represents the most evidence-based first-line adjunctive therapy for this clinical scenario. |

*Supp Table 3: Openevidence response for each case*

### Supp. Table 6- Gemini Qualitative error analysis:

| case number | case | Gemini response | Assessment arbitrated Likert score | Plan arbitrated Likert score | What is the mistake? |
|---|---|---|---|---|---|
| 4 | 75 f, ipd 10 years, tx levocarb 100/25 2 tab qid q4h, Levocrab CR 200/50 QHS, rasagiline 1mg qd, entacapone 200 md QID. Myrbetriq 50 qd. Laxaday 17 gr QD. Describes that at least once a day and sometimes more often there are events in which she does not go ON after taking LEVOCARB. States that she takes LEVOCARB half an hour before meals. And has regular bowel movements every day. | A. The most likely clinical impression is refractory delayed or failed motor response (delayed or absent ON) to levodopa in a patient with fluctuating Parkinson's disease.<br><br>P. Add opicapone to the current regimen to enhance the beneficial effects of levodopa and reduce OFF time. Opicapone 50 mg PO once daily. | 4 | 1 | The diagnosis is correct, but the plan adds a medication from the same class that the patient is already receiving, just a different version. The addition is made without discontinuing the prior medication. This is highly unacceptable and, in practice, an error. |

| | | | | | |
|---|---|---|---|---|---|
| 33 | 72 f, ipd 10 years, tx levocarb 100/25 2 tab qid q4h, entacapone 200 mg qid, rasagiline 1 mg qd. Her walking has become very impaired: she takes short, shuffling steps and has a stooped posture. She frequently has near-falls if she turns too quickly or if someone bumps into her, so she has to concentrate hard on every step to avoid stumbling. She now uses a walker for safety. | A. The most likely clinical impression is advanced Parkinson's disease with refractory gait failure and postural instability despite optimized medical management.<br><br>P. Given the inadequate response to maximal medical therapy and the patient's gait impairment, a definitive treatment plan is to evaluate the patient for Deep Brain Stimulation (DBS) therapy to improve motor fluctuations and gait symptoms. | 3 | 1 | The diagnosis is within the correct domain but is too extreme—the model determined that the condition is refractory and advanced, although this was not suggested by the case. The plan proposed is therefore also too extreme; moreover, the plan itself is incorrect, as it suggests an advanced therapy that may indeed help a range of problems, but not the problem that was presented. |
| 47 | 79 m, ipd 11 years , tx levocarb 100/25 2.5 tab qid q4h + donepezil 10 mg qd. His wife reports that he now experiences vivid and disturbing dreams almost every night. Sleep is fragmented, and he wakes up anxious. | A. The most likely clinical impression is REM Sleep Behavior Disorder (RBD) in the setting of long-standing Parkinson's disease, potentially exacerbated by existing dopaminergic therapy.<br>P. The treatment plan is to initiate clonazepam 0.5 mg PO at bedtime to manage the dream-related sleep disturbance and anxiety. The dose can be titrated based on clinical response, while monitoring for daytime sedation and balance. | 1 | 2 | The diagnosis is correct. The treatment line proposed by the model corresponds to second-line therapy; however, there is a simpler solution with fewer side effects at the first-line level, and therefore it should be attempted first. |

| | | | | | |
|---|---|---|---|---|---|
| 27 | 72 f, ipd 10 years, tx levocarb 100/25 2 tab qid q3h, entacapone 200 mg qid. Safinamide 100 mg qd, laxaday 17 g qd. She finds it hard to shower; she's afraid of losing balance in the slippery tub and her slow movements make it exhausting to wash and dry herself. She has started relying on her daughter to help with bathing and other hygiene tasks several times a week. | I am not sure, considering either Deep Brain Stimulation (DBS) or a Levodopa-Carbidopa Intestinal Gel (LCIG) infusion. | 1 | 2 | There is no diagnosis at all, and the proposed treatment is an invasive therapy for advanced Parkinson's disease, for which no appropriate details were presented in the case. |
| 3 | 75 m, ipd 10 years, tx levocarb 100/25 2 tab qid q4h, Levocrab CR 200/50 QHS, rasagiline 1mg qd, entacapone 200 md QID. Myrbetriq 50 qd. Random events are reported during the day where after taking LEVOCARB it takes a long time - sometimes more than an hour - for the drug to start working. takes the LEVOCARB 30 min before meals. | A: The patient is experiencing delayed onset of action of immediate-release levodopa, which is most likely due to food interference with absorption.<br><br>P: The most appropriate first-line intervention is to optimize medication timing by instructing the patient to take the immediate-release levocarb on an empty stomach: Levocap 100/25 two tablets by mouth four times daily, 60 minutes before meals. | 2 | 2 | The diagnosis is correct, but the therapeutic recommendation is very general and conservative and will most likely not be beneficial (though it will also not cause harm). |

| | | | | | |
|---|---|---|---|---|---|
| 10 | 80 f, ipd 10 years, tx levocarb 100/25 2 tab qid q3h (8-11-14-17-20), safinamide 100 qd, amantadine 100 mg bid. She reports seeing children in her living room who aren’t actually there, even in the middle of the day. Sometimes she also sees small animals darting around, or thinks she sees her deceased relatives momentarily. She is often able to recognize these visions are not real once she checks, but they are frightening and have caused her anxiety. | A: The most likely diagnosis is Parkinson's Disease Psychosis (PDP), presenting as visual hallucinations with preserved insight, a common non-motor complication of Parkinson's disease.<br><br>P: Initiate treatment with the first FDA-approved medication for this condition to manage psychotic symptoms without worsening motor control: Pimavanserin 34 mg PO once daily. Continue current Parkinson's disease medications but closely monitor for any potential drug interactions or adverse effects, especially increased anxiety. | 4 | 2 | The diagnosis is correct; however, the plan recommends a medication for which the indication may be appropriate, but in practice it is not customary to use it as first- or second-line therapy. In addition, the initial step in this patient is to reduce the dosage to the point of discontinuation of two medications that may exacerbate hallucinations, and this was not done. |

| | | | | | |
|---|---|---|---|---|---|
| 15 | 63 f, ipd 8 years, tx levocarb 100/25 1.5 tab qid q4h, Rasagiline 1 mg qd, citalopram 10 mg qd, laxaday 17 g qd prn. She has difficulty sleeping through the night, waking up every few hours and struggling to get back to sleep. She feels unrefreshed in the morning. There is no difficulty with initial sleep onset. The patient is unable to clearly identify the reason for awakening during the night; at times it is to use the bathroom, though not consistently. She does not experience any difficulty moving to the bathroom upon awakening. | A: The most likely diagnosis is Insomnia/Fragmented Sleep, a common non-motor symptom of Parkinson's disease, with potential contributions from levodopa-related sleep disturbances or nocturnal motor symptoms.<br><br>P: Initiate a short course of a non-benzodiazepine sedative hypnotic to improve sleep maintenance. Rx: Zolpidem 5 mg PO QHS PRN, ensuring no significant nocturnal motor symptoms (like tremor or difficulty turning) are the primary cause of awakening before escalating therapy. | 5 | 2 | The diagnosis is correct, but the plan proposes treatment with a medication that indeed is commonly used for this indication, but typically in an acute setting, and it is not recommended to administer this medication long term. Given that we are dealing with a chronic disease, it is not appropriate to provide a treatment that is intended to be temporary as a permanent solution. Moreover, in Parkinson's disease there are several other accepted options to treat this condition, and this was not the proposed approach. |

| | | | | | |
|---|---|---|---|---|---|
| 23 | 66-year-old, F IPD 10-year duration, Tx Levocarb 100/25 2 tabs 5 times a day, Sinemet CR 200/50 1 tab qhs, safinamide 100 md qd, Citalopram 20 mg qd, melatonin 10 mg qhs. complaining about bothersome drooling. no conection to wearing off times. No Treatment was tried. | A: This patient presents with sialorrhea (drooling) secondary to Idiopathic Parkinson's Disease, which is unrelated to fluctuating motor symptoms or medication wearing off.<br><br>P: I recommend initiating an oral anticholinergic agent to reduce saliva production. The treatment plan is to start glycopyrrolate 1 mg PO TID, titrating carefully based on response and tolerability, particularly for potential side effects such as dry mouth or cognitive impairment. | 5 | 2 | The diagnosis is correct, but the recommendation to initiate an anticholinergic medication is highly unfavorable in Parkinson’s disease, as it may worsen cognitive status and can also cause hallucinations. There are multiple alternative solutions that the model did not propose. |

| | | | | | |
|---|---|---|---|---|---|
| 32 | 65 m, ipd 6 years, tx levocarb 100/25 1.5 tab qid q4h, safinamide 100 mg qd, myrbetriq 50 mg qd. He has great difficulty rising from low positions — getting out of bed in the morning or lifting himself out of his car seat or a soft couch has become a slow, taxing process. In the mornings, he often needs to roll to the side and push up with his arms to sit at the edge of the bed, then gather strength to stand. When exiting his car, he has to rock back and forth a few times to gain momentum to stand up, and often requires someone to hold the door or give him a hand to pull him out of a deep chair. | A. This patient is likely experiencing motor "off" periods, manifesting as severe levodopa-responsive bradykinesia and difficulty with transitions/axial movements, despite being on a complex dopaminergic regimen.<br>P. Increase the frequency of levocarb to maximize "on" time and reduce specific "off" period motor symptoms: levocarb 100/25 tab q4h (total 5 doses) PO. | 3 | 2 | The diagnosis is incorrect—the model determined that this represents OFF, although it is not clear from the case that this is indeed the situation. There was a need to ask many additional questions in order to understand the source of the problem. Because the diagnosis is incorrect, the recommendation was also not relevant. |

| | | | | | |
|---|---|---|---|---|---|
| 40 | 71 f, ipd 7 years, tx levocarb 100/25 1.5 tab qid q4h + selegiline 5 mg bid. Since starting selegiline, she reports difficulty falling asleep and frequent nighttime awakenings. No vivid dreams or anxiety. Daytime fatigue increased. | **A** The patient is experiencing selegiline-induced insomnia, manifesting as difficulty falling asleep and frequent nighttime awakenings, leading to increased daytime fatigue.<br><br>**P** To address the sleep disturbance likely caused by selegiline, discontinue selegiline 5 mg bid and initiate a short-acting non-benzodiazepine for sleep-onset insomnia: zolpidem 5 mg PO qhs PRN. | 4 | 2 | The diagnosis is also not precise—it is possible, but it is also possible that additional contributing factors exist, and the model did not inquire further. In addition, the therapeutic recommendation is incorrect—it suggests a medication that is indeed indicated for this condition, but only for short-term use; however, since Parkinson’s disease is chronic and long-term, it is not customary to use such treatments as first- or second-line therapy. |
| 44 | 70 f, ipd 6 years, tx levocarb 100/25 2 tab qid q4h + ropinirole 4 mg tid. The patient recently replaced his car with a luxury vehicle beyond his financial means, using most of his savings to do so. He did not consult his wife beforehand and describes the decision as an impulsive act, stating that he “just felt like doing it.” | A: The most likely diagnosis is impulse control disorder, specifically pathological spending, induced by dopamine agonist therapy (ropinirole) in a patient with Parkinson's disease.<br><br>P: Discontinue ropinirole 4 mg tid immediately and manage any withdrawal symptoms; levocarb should be continued at the current dose. | 5 | 2 | The diagnosis is correct, and the concept of the therapeutic recommendation is correct—discontinuation of the medication. However, this discontinuation should be performed gradually; otherwise, there is a risk of another adverse condition. The fact that the model recommended immediate discontinuation without tapering is the issue in its response. |

| | | | | | |
|---|---|---|---|---|---|
| 45 | 61 f, ipd 4 years, tx levocarb 100/25 1 tab qid q4h + rotigotine patch 4 mg qd. Reports redness, itching, and mild peeling of skin at the patch site after two weeks of use. No systemic rash or allergy history. | A. The most likely clinical impression is a common application site reaction related to the rotigotine transdermal system in a patient with Parkinson's disease.<br>P. Discontinue the current rotigotine transdermal patch due to the skin irritation and switch the patient to an equivalent dose of an oral nonergot dopamine agonist. The new therapeutic strategy is to initiate ropinirole 0.5 mg PO TID and titrate weekly as tolerated to maintain dopamine agonist effect. | 5 | 2 | The diagnosis is correct, and the therapeutic concept is correct—but as a second-line approach. As a first-line approach, there are additional simple measures that can be attempted in order to preserve the current first-line treatment. |

*Supp Table 4: Gemini Qualitative error analysis*

### Supp. Table 7- OpenEvidence Qualitative error analysis:

| case number | case | openevidance response | Assessment arbitrated Likert score | Plan arbitrated Likert score | What is the mistake? |
|---|---|---|---|---|---|

| | | | | | |
|---|---|---|---|---|---|
| 50 | 62 f, ipd 7 years, tx levocarb 100/25 1.5 tab qid, entacapone 200 mg QID. She experiences clear wearing-off symptoms about 30–45 minutes before her next LEVOCARB dose is due. Her hands begin to tremble, her legs stiffen, and she feels anxious and fatigued. These motor and non-motor symptoms resolve shortly after taking the next dose, but the fluctuation disrupts her ability to plan activities and leaves her feeling vulnerable and dependent on the clock. | A: The patient demonstrates classic motor fluctuations that typically emerge after several years of levodopa therapy due to the short half-life of levodopa, inconsistent gastrointestinal absorption, and progressive dopaminergic neuronal degeneration.<br><br>Plan: Add opicapone 50 mg once daily at bedtime (at least 1 hour before or after levodopa/carbidopa doses). | 4 | 1 | Correct diagnosis, but recommendation to add a medication from the same class that the patient is already taking, without discontinuing the prior medication—this is not appropriate. |

| | | | | | |
|---|---|---|---|---|---|
| 1 | 65 m, ipd 5 years, tx levocarb 100/25 2 tab qid q4h, safinamide 100 mg qd. Reports involuntary movements of the limbs during the day. He usually does not notice this himself, but his wife asks him why he cannot sit comfortably in a chair. There is fluctuation during the day. It seems to him that it usually happens about an hour after taking LEVOCARB, but not necessarily. It does not interfere with his function, but it bothers his wife to see him like this. | A: The patient exhibits involuntary limb movements occurring approximately one hour after levodopa dosing, consistent with peak-dose dyskinesia—the most common pattern of levodopa-induced dyskinesia.[6] The movements are non-disabling and do not interfere with function, though they cause caregiver distress. The temporal relationship to levodopa administration and the choreiform nature of the movements support this diagnosis.<br>P: Add amantadine extended-release 274 mg once daily at bedtime.<br>Amantadine is the only FDA-approved medication with established efficacy for levodopa-induced dyskinesia.[2-3][5] A recent network meta-analysis of 85 randomized controlled trials identified immediate-release amantadine as the highest-ranking oral medication for dyskinesia reduction (SUCRA = 86.5%).[1] Extended-release | 5 | 2 | The diagnosis is correct, but the recommendation to add a medication that is indeed indicated represents a second-line approach; the appropriate first-line step would be to attempt adjustment of the dosages and timing of the medications the patient is already taking. |

| | | |
|---|---|---|
| | | formulations provide superior pharmacokinetics with once-daily evening dosing, offering higher and more continuous plasma levels compared to immediate-release formulations that require up to three times daily administration.[4-5]<br>The extended-release formulation has demonstrated efficacy in reducing dyskinesia and "off" time in pivotal trials.[4] Two extended-release preparations are FDA-approved for this indication.[5] The recommended starting dose is 274 mg once daily at bedtime, which can be titrated based on response and tolerability. |

| | | | | | |
|---|---|---|---|---|---|
| 2 | 66 f, ipd 7 years, tx levocarb 100/25 1.5 tab qid q4h, Levocrab CR 200/50 QHS, rasagiline 1mg qd, entacapone 200 md QID. Myrbetriq 50 qd, laxaday 17 g qd. Describes a recurrence of the tremor in her right hand, a general bad feeling including anxiety, and FREEZING OF GAIT events quite regularly about 40 minutes before she is supposed to take her next dose of LEVOCARB. | Assessment: The patient is experiencing classic motor fluctuations with predictable wearing-off despite being on combination therapy with levodopa, a MAO-B inhibitor (rasagiline), and a COMT inhibitor (entacapone). The symptoms occurring 40 minutes before the next dose suggest inadequate duration of levodopa effect despite the use of entacapone to prolong levodopa availability.<br><br>Plan: Add amantadine immediate-release 100 mg three times daily (morning, midday, and early evening). Rationale: Amantadine provides additional benefit for motor fluctuations and wearing-off symptoms through NMDA receptor antagonism. It can reduce off-time and may also help with dyskinesias if they develop. The three-times-daily dosing will provide coverage throughout the waking hours when wearing-off is problematic. | 5 | 2 | The diagnosis is correct, but the recommendation to add a medication that is indeed indicated represents a second-line approach; the appropriate first-line step would be to attempt adjustment of the dosages and timing of the medications the patient is already taking. |

| | | |
|---|---|---|
| | | Titration: Start with 100 mg once daily for 1 week, then increase to 100 mg twice daily for 1 week, then advance to the target dose of 100 mg three times daily. The last dose should be given no later than early evening to minimize insomnia risk. Monitoring: Reassess in 4-6 weeks for improvement in wearing-off symptoms, freezing of gait, and anxiety associated with off periods. Monitor for potential side effects including livedo reticularis, peripheral edema, hallucinations, and confusion. |

| | | | | | |
|---|---|---|---|---|---|
| 17 | 64 f, ipd 4 years, tx levocarb 100/25 1.5 tab tid, gabapentin 300 mg qhs. She experiences aching muscle pain in her shoulders and arms that tends to worsen when her Parkinson's symptoms are more pronounced. The chronic pain significantly affect her quality of life, often requiring use of pain relievers, with various outcomes. | A: 64-year-old female with idiopathic Parkinson's disease (4-year duration) presenting with chronic musculoskeletal pain in shoulders and arms that worsens with motor symptom fluctuations, consistent with Parkinson's disease-related fluctuation-associated nociceptive pain.<br>P: Optimize dopaminergic therapy as the first-line intervention for Parkinson's disease-related pain.[1-2] The current levodopa dosing schedule may be inadequate to maintain stable dopaminergic levels throughout the day, contributing to fluctuation-related pain.<br><br>Treatment Recommendation:<br><br>Increase levodopa/carbidopa to 25/100 mg, 2 tablets four times daily (total daily levodopa dose: 800 mg divided into more frequent dosing). This adjustment provides both increased total daily levodopa and more frequent dosing intervals to reduce motor | 5 | 2 | The plan has inappropriately converted a second-line approach into a first-line approach. |

| | | |
|---|---|---|
| | | fluctuations and associated pain.[1]<br><br>Rationale: Optimization of dopaminergic treatment should always be the initial step in managing Parkinson's disease-related chronic pain, as dopaminergic deficiency is often a key contributor to pain syndromes.[1] More frequent dosing intervals help maintain stable dopaminergic levels and reduce off-period symptoms, including fluctuation-related musculoskeletal pain.[1] This approach is supported by current treatment algorithms that prioritize dopaminergic optimization before considering non-dopaminergic analgesics.[1]<br><br>Additional Considerations:<br><br>Discontinue gabapentin 300 mg at bedtime, as this dose is subtherapeutic and unlikely to provide benefit for her pain type<br><br>Monitor for hyperdopaminergic effects including dyskinesia, orthostatic |

| | | |
|---|---|---|
| | | hypotension, and neuropsychiatric symptoms with increased levodopa dosing[1]<br><br>Reassess pain severity in 2-4 weeks using a standardized pain scale<br><br>If pain persists despite optimized dopaminergic therapy, consider adding a COMT inhibitor (e.g., entacapone) or MAO-B inhibitor to further stabilize dopaminergic levels[1]<br><br>Non-pharmacological interventions including physical therapy, stretching exercises, and massage should be incorporated as adjunctive measures[2][4]<br><br>If pain remains refractory after dopaminergic optimization, consider trial of duloxetine or prolonged-release oxycodone/naloxone, though evidence for the latter is mixed[1-3][5]<br><br>Follow-up:<br><br>Schedule return visit in 2-4 weeks to assess treatment response, monitor for adverse effects, and adjust |

| | | |
|---|---|---|
| | | therapy as needed. |

| | | | | | |
|---|---|---|---|---|---|
| 21 | 62 f, ipd 6 years, tx levocarb 100/25 1 tab qid q4h, Rasagiline 1 mg qd, amantadine 100 mg tid. She experiences overwhelming sense of exhaustion that is not relieved by rest. Even after a full night's sleep, she often feels she has no energy to do her usual activities or exercises. | Assessment: Fatigue is a common and debilitating non-motor symptom affecting 40-50% of patients with Parkinson's disease.[3-4] This symptom significantly impacts quality of life and represents a leading cause of disability in PD.[3] The patient's current medication regimen includes rasagiline, which has demonstrated efficacy in reducing physical fatigue in PD, though evidence suggests potential dose-dependent effects.[1-3]<br>Plan: Primary recommendation: Increase rasagiline from 1 mg daily to 2 mg daily (maximum approved dose).<br><br>Rationale: High-quality evidence demonstrates that rasagiline is associated with reduced progression of physical fatigue in PD.[1-2] A large randomized controlled trial (N=1176) showed that rasagiline significantly slowed fatigue progression compared to placebo (SMD -0.27, 95% CI -0.39 to -0.16).[1-2] A pilot study specifically examining | 5 | 2 | The plan proposed by the model is highly unconventional in practice—use of a medication known in Parkinson's disease at a dose double the accepted dose for the given indication. The model supports its recommendation based on a single study, which is an error, as clinical practice is generally not based on a single study but rather on multiple studies demonstrating efficacy. |

| | | |
|---|---|---|
| | | rasagiline for fatigue treatment showed significant improvement in fatigue scores at 1 mg daily, and the pivotal trials included both 1 mg and 2 mg daily dosing with combined benefit.[2-3] Rasagiline represents the intervention with the strongest evidence base for fatigue management in PD, and dose optimization may provide additional benefit.[2][5-6] |

| | | | | | |
|---|---|---|---|---|---|
| 23 | 66-year-old, F IPD 10-year duration, Tx Levocarb 100/25 2 tabs 5 times a day, Sinemet CR 200/50 1 tab qhs, safinamide 100 md qd, Citalopram 20 mg qd, melatonin 10 mg qhs. complaining about bothersome drooling. no conection to wearing off times. No Treatment was tried. | Assessment: Sialorrhea affects up to 74% of patients with Parkinson's disease and results from inefficient elimination of oral saliva rather than increased production. This symptom can cause significant embarrassment, social stigma, perioral skin excoriation, and aspiration risk.<br>Plan:<br>Botulinum toxin injection into the parotid and submandibular glands is recommended as first-line therapy for this patient's sialorrhea. Specifically, incobotulinumtoxinA 100 units total dose (divided between bilateral parotid and submandibular glands) is the committed treatment recommendation.<br><br>IncobotulinumtoxinA 100 units has demonstrated significant reduction in unstimulated salivary flow rate and improvement in patient-reported outcomes at 4 weeks post-injection, with therapeutic effects lasting approximately 16 weeks. This represents FDA-approved therapy for chronic | 5 | 2 | Recommendation of a second-line treatment as first-line—i.e., this is too aggressive. |

| | | |
|---|---|---|
| | | sialorrhea in adults. The treatment is well-tolerated with the most common adverse events being mild to moderate dry mouth (2.7%) and dysphagia (0% at this dose).<br><br>Alternative botulinum toxin formulations with evidence for sialorrhea include rimabotulinumtoxinB 2500-3500 units, which showed significant efficacy with onset as early as 1 week post-injection. However, incobotulinumtoxinA is preferred given its FDA approval specifically for this indication and favorable safety profile at the 100-unit dose.<br><br>Oral anticholinergic agents such as glycopyrrolate (1 mg three times daily) represent an alternative approach but carry risks of systemic anticholinergic effects including dry mouth, constipation, urinary retention, and potential cognitive impairment—particularly problematic in patients with Parkinson's disease. Given these concerns |

| | | |
|---|---|---|
| | | and the patient's advanced disease duration, botulinum toxin injection is the superior choice. |

| | | | | | |
|---|---|---|---|---|---|
| 27 | 72 f, ipd 10 years, tx levocarb 100/25 2 tab qid q3h, entacapone 200 mg qid. Safinamide 100 mg qd, laxaday 17 g qd. She finds it hard to shower; she's afraid of losing balance in the slippery tub and her slow movements make it exhausting to wash and dry herself. She has started relying on her daughter to help with bathing and other hygiene tasks several times a week. | A: Despite this comprehensive regimen including levodopa, a COMT inhibitor (entacapone), and an MAO-B inhibitor (safinamide), the patient demonstrates persistent motor fluctuations with bradykinesia significantly impairing function. Her fear of falling and need for assistance with bathing indicate both motor symptom inadequacy and safety concerns that warrant intervention.<br><br>Plan: Medication Adjustment:<br>Add opicapone 50 mg once daily at bedtime, and discontinue entacapone. Opicapone is a once-daily COMT inhibitor that has demonstrated efficacy in reducing off-time in patients with Parkinson's disease experiencing motor fluctuations.[1-3] In the International Parkinson and Movement Disorder Society evidence-based review, opicapone was assessed as efficacious for treating motor fluctuations.[3] Opicapone offers the advantage of once-daily dosing | 2 | 2 | A very non-specific case was presented, and instead of asking further questions, the model decided on a specific problem, although it is not at all certain from the case that this is the issue (this may also reflect a prompting issue). From a therapeutic standpoint: addition of a medication from the same class that the patient is already receiving, without discontinuation of the prior medication, which may lead to significant adverse effects. |

| | | |
|---|---|---|
| | | compared to entacapone's four-times-daily regimen, which may improve medication adherence and provide more consistent COMT inhibition throughout the day.[2][6]<br>The starting dose of opicapone is 50 mg once daily, taken at bedtime at least one hour before or after levodopa/carbidopa.[6-7] This can be titrated to 25 mg daily if tolerability issues arise, though 50 mg is the standard therapeutic dose.<br><br>Physical Therapy Referral:<br>Refer for comprehensive physical therapy evaluation with focus on gait and balance training. Exercise interventions including gait and balance training, progressive resistance training, and strength training have demonstrated benefit for motor symptoms, balance, gait, and function in Parkinson's disease.[4] The American Geriatrics Society recommends optimizing exercise, strengthening, balance, and physical therapy |

| | | |
|---|---|---|
| | | at time of diagnosis and throughout the course of care.[5] Physical therapy can provide strategies to improve safety during bathing and other activities of daily living, potentially reducing fall risk and improving independence.[4]<br><br>Occupational Therapy Consideration: Consider occupational therapy consultation to assess home safety modifications, particularly bathroom adaptations (grab bars, non-slip mats, shower chair) and to develop compensatory strategies for hygiene tasks.[4] |

| | | | | | |
|---|---|---|---|---|---|
| 30 | 74 m, ipd 10 years, tx levocarb 100/25 2 tab qid q4h, levocarb CR 200/50 QHS, melatonin 10 mg qhs, Escitalopram 10 mg qd. He finds it nearly impossible to turn over in bed at night. If he wants to shift positions, he has to wake up fully and sometimes even sit upright to reposition himself. | A: Nocturnal hypokinesia affects up to 70% of patients with mid-to-advanced Parkinson's disease and significantly impairs sleep quality and quality of life. The patient's current symptoms suggest that the bedtime controlled-release formulation is providing insufficient dopaminergic stimulation during the night.<br>P: Primary Treatment Recommendation: Add rotigotine transdermal patch 4 mg/24 hours at bedtime, titrating up by 2 mg weekly to a target dose of 8-10 mg/24 hours as tolerated. Rotigotine provides continuous dopaminergic stimulation throughout the night and has demonstrated significant improvement in nocturnal hypokinesia in randomized controlled trials, with objective improvements in number of turns in bed and degree of axial rotation, as well as improvements in validated sleep scales. Extended-release dopamine agonists like rotigotine control | 5 | 2 | Correct diagnosis, but the plan is overly aggressive in two ways—use of a third-line medication as first-line, and the use of excessively high doses, which may lead to significant adverse effects. |

| | | |
|---|---|---|
| | | nighttime motor symptoms and improve sleep quality based on multiple assessment modalities. This approach is recommended for moderate to advanced Parkinson's disease with nocturnal symptoms. |

| | | | | | |
|---|---|---|---|---|---|
| 32 | 65 m, ipd 6 years, tx levocarb 100/25 1.5 tab qid q4h, safinamide 100 mg qd, myrbetriq 50 mg qd. He has great difficulty rising from low positions — getting out of bed in the morning or lifting himself out of his car seat or a soft couch has become a slow, taxing process. In the mornings, he often needs to roll to the side and push up with his arms to sit at the edge of the bed, then gather strength to stand. When exiting his car, he has to rock back and forth a few times to gain momentum to stand up, and often requires someone to hold the door or give him a hand to pull him out of a deep chair. | A: The patient demonstrates classic features of motor fluctuations with end-of-dose wearing off, manifesting as significant difficulty rising from low positions (bed, car seat, soft couch). These symptoms are consistent with the shortened duration of levodopa effect that typically emerges after 4 years of treatment. His difficulty with transfers represents a specific motor complication that warrants optimization of dopaminergic therapy. P: Add opicapone 50 mg once daily at bedtime to extend levodopa duration and reduce OFF time. | 2 | 2 | The diagnosis is not necessarily correct—the presented case includes multiple problems, each of which may have a different cause. The model independently determined the problem, although it is not at all clear that this is correct based on the case. |

| | | | | | |
|---|---|---|---|---|---|
| 35 | 67 m, ipd 6 years, tx levocarb 100/25 2 tab qid q4h, safinamide 100 mg qd. Reports irregular, involuntary movements of the limbs and trunk occurring about 45–60 minutes after each levocarb dose. Movements are more noticeable in the afternoon. He remains functionally independent. In general, the patient's wife notices these movements more than he does himself. | A: 67-year-old male with 6-year history of idiopathic Parkinson's disease presenting with peak-dose dyskinesia. The patient experiences irregular, involuntary movements of the limbs and trunk occurring 45–60 minutes after each levodopa/carbidopa dose, more prominent in the afternoon. These movements are more noticeable to his wife than to himself, which is typical of mild dyskinesia. The timing—occurring at the expected peak plasma concentration of levodopa—is consistent with peak-dose dyskinesia, the most common form of levodopa-induced dyskinesia.<br>P: Add amantadine extended-release 274 mg orally once daily at bedtime to reduce dyskinesia. Amantadine is the only medication with proven efficacy for treating levodopa-induced dyskinesia and is FDA-approved for this indication. The extended-release formulation provides higher | 4 | 2 | Recommendation to use a third-line medication as first-line. |

| | | |
|---|---|---|
| | | and more continuous plasma levels compared to immediate-release formulations, allowing once-daily evening dosing while improving both dyskinesia and off-time. In pivotal trials, amantadine ER demonstrated significant reduction in dyskinesia severity.<br><br>Alternative strategies—such as reducing individual levodopa doses or fractionating the dosing schedule—could theoretically reduce peak-dose dyskinesia, but would likely worsen motor control and increase off-time given the patient's current four-times-daily dosing regimen. Since the patient remains functionally independent and the dyskinesia is mild (more noticeable to family than patient), adding an antidyskinetic agent is preferable to reducing dopaminergic therapy.<br><br>The patient should be monitored for potential amantadine side |

| | | |
|---|---|---|
| | | effects including visual hallucinations, peripheral edema, and livedo reticularis. If dyskinesia persists or worsens despite amantadine therapy, consider referral to a movement disorders specialist for evaluation of advanced therapies including deep brain stimulation. |

| | | | | | |
|---|---|---|---|---|---|
| 41 | 63 m, ipd 7 years, tx levocarb 100/25 2 tab at 7-10-13-16-19-22, Sinemet CR 200/50 1 tab QHS, safinamide 100 mg qd. New episodes have emerged in which the patient reports seeing animals quickly passing in front of him. He recognizes that these are not real, and they do not cause him fear or distress. | A:This presentation is consistent with Parkinson's disease psychosis (PDP), which occurs in 18-50% of patients with Parkinson's disease.<br>P: Rule out precipitating factors: Evaluate for delirium triggers including intercurrent infection, constipation, metabolic disturbances, or new medications, as these are the most frequent causes of acute or worsening psychosis symptoms.<br>Medication review: The current dopaminergic regimen is relatively high-dose. However, given the patient's lack of distress and preserved insight, immediate reduction of anti-parkinsonian medications is not necessary at this time.<br>Initiate pimavanserin: Start pimavanserin 34 mg once daily. Pimavanserin is the only FDA-approved medication for hallucinations and delusions associated with Parkinson's disease psychosis and is recommended as first-line treatment | 4 | 2 | Correct diagnosis, but recommendation of a third-line medication as a first-line approach. |

| | | |
|---|---|---|
| | | for mild psychotic symptoms that do not require rapid response. As a selective 5-HT2A inverse agonist, pimavanserin does not worsen motor function and has a favorable tolerability profile compared to traditional antipsychotics.<br><br>Alternative options if pimavanserin fails or is not tolerated:<br><br>Quetiapine (starting at 12.5-25 mg at bedtime) for symptoms requiring more rapid improvement, though evidence for efficacy is limited<br>Expert Opinion on Drug Safety |

| | | | | | |
|---|---|---|---|---|---|
| 47 | 79 m, ipd 11 years , tx levocarb 100/25 2.5 tab qid q4h + donepezil 10 mg qd. His wife reports that he now experiences vivid and disturbing dreams almost every night. Sleep is fragmented, and he wakes up anxious. | A: The clinical presentation is most consistent with REM sleep behavior disorder (RBD), a common parasomnia in Parkinson's disease characterized by dream enactment behaviors and vivid dreams. The patient's donepezil is a likely contributing factor, as acetylcholinesterase inhibitors including donepezil can trigger or worsen RBD symptoms. Additionally, donepezil significantly increases REM sleep percentage and may contribute to sleep fragmentation.<br><br>Levodopa at the current dosing regimen may also contribute to sleep disturbances, including nocturnal hallucinations and fragmented sleep, particularly at higher doses. However, the temporal relationship with donepezil initiation and the known association between cholinesterase inhibitors and RBD make donepezil the more likely primary contributor. | 2 | 2 | Correct diagnosis. A three-step treatment plan was presented. The first line is incorrect because it explicitly prioritizes one aspect of the disease over another, whereas the other aspect should clearly be preserved to a greater extent; however, this was not appropriately considered, and this is an error. The subsequent lines are correct. |

| | | |
|---|---|---|
| | | P: Discontinue donepezil 10 mg once daily. The risks of continuing donepezil—specifically worsening RBD symptoms and sleep fragmentation—outweigh the cognitive benefits in this clinical context. SSRIs and tricyclic antidepressants should also be avoided or reconsidered if introduced in the future, as these can similarly trigger or worsen RBD.<br><br>Reassess sleep symptoms 2-4 weeks after donepezil discontinuation. If vivid dreams and sleep fragmentation persist despite stopping donepezil, consider initiating pharmacologic treatment for RBD with either:<br><br>Melatonin 3 mg at bedtime (preferred initial option given favorable safety profile), or<br><br>Clonazepam 0.5-1 mg at bedtime (alternative if melatonin ineffective) |

| | | | | | |
|---|---|---|---|---|---|
| 15 | 63 f, ipd 8 years, tx levocarb 100/25 1.5 tab qid q4h, Rasagiline 1 mg qd, citalopram 10 mg qd, laxaday 17 g qd prn. She has difficulty sleeping through the night, waking up every few hours and struggling to get back to sleep. She feels unrefreshed in the morning. There is no difficulty with initial sleep onset. The patient is unable to clearly identify the reason for awakening during the night; at times it is to use the bathroom, though not consistently. She does not experience any difficulty moving to the bathroom upon awakening. | A: The patient meets criteria for chronic insomnia disorder with predominant sleep maintenance difficulty. While Parkinson's disease commonly causes sleep fragmentation, her preserved mobility upon awakening and inconsistent relationship to nocturia suggest the insomnia may have a significant primary component amenable to targeted treatment.<br><br>Plan: Cognitive behavioral therapy for insomnia (CBT-I) is recommended as first-line treatment for chronic insomnia disorder.[1-2] CBT-I includes sleep restriction therapy, stimulus control, relaxation techniques, and cognitive therapy targeting maladaptive beliefs about sleep.[5-6] This multicomponent approach produces substantial and sustained improvements in sleep maintenance, with evidence showing superiority over pharmacotherapy in long-term outcomes.[2] CBT-I can be delivered via | 2 | 4 | Correct diagnosis. The plan is too conservative and will most likely not be beneficial (though it will not cause harm). |

| | | |
|---|---|---|
| | | individual therapy, group sessions, or telehealth platforms.[2][5] Refer to behavioral sleep medicine specialist or trained clinician for CBT-I |

*Supp Table 5: OpenEvidence Qualitative error analysis*

### Supplementary figures:

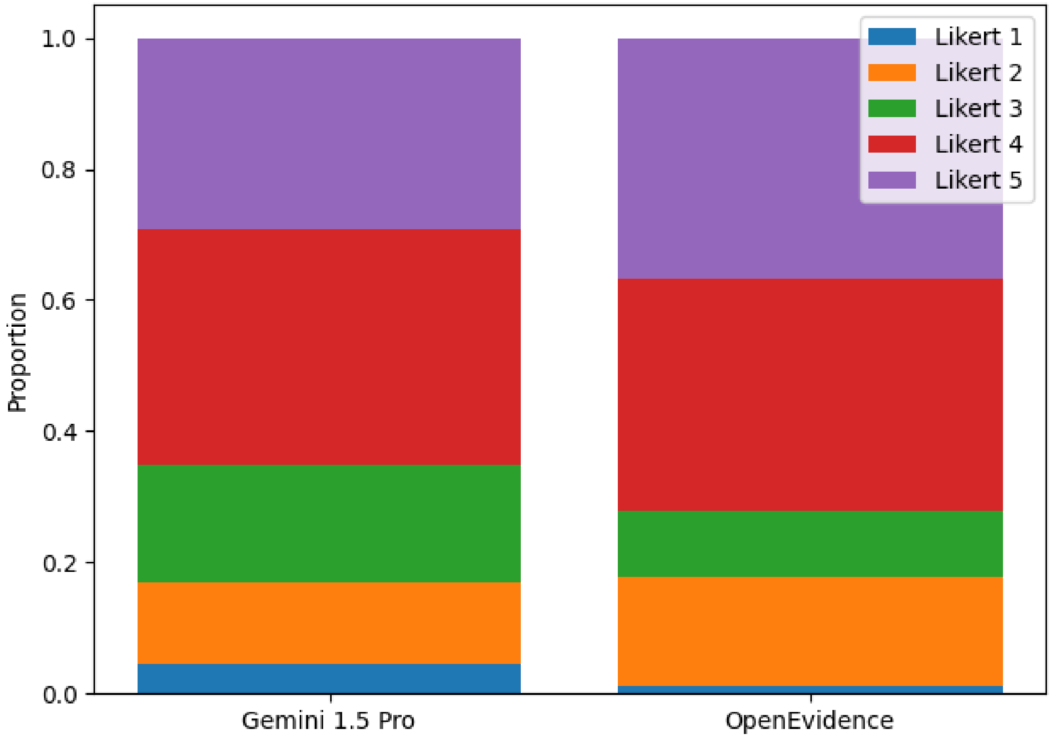

*Supp Figure 1: Likert score distribution by model*

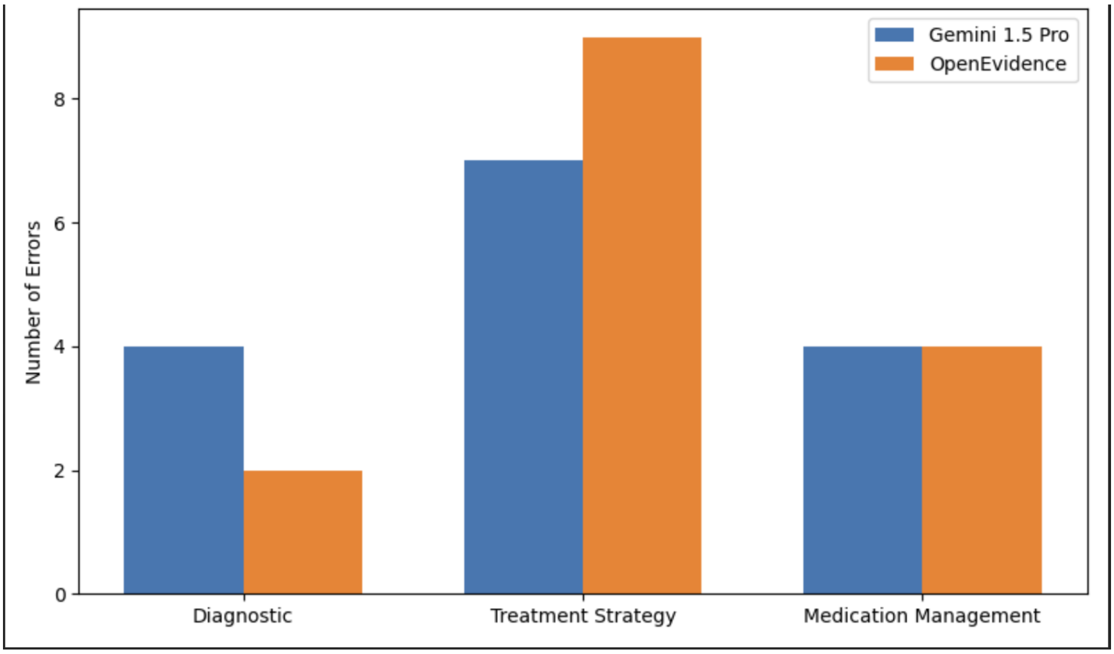

*Supp Figure 2: Distribution of Error Categories by Model*
